# Automated diagnosis of autism: State of the art

**DOI:** 10.1101/2021.06.29.21254249

**Authors:** Amir Valizadeh, Mana Moassefi, Amin Nakhostin-Ansari, Soheil Heidari Some’eh, Hossein Hosseini-Asl, Mehrnush Saghab Torbati, Reyhaneh Aghajani, Zahra Maleki Ghorbani, Iman Menbari-Oskouie, Faezeh Aghajani, Alireza Mirzamohamadi, Mohammad Ghafouri, Shahriar Faghani, Amir Hossein Memari

## Abstract

**Background:** Autism spectrum disorder (ASD) represents a panel of conditions that begin during the developmental period and result in impairments of personal, social, academic, or occupational functioning. Early diagnosis is directly related to a better prognosis. Unfortunately, the diagnosis of ASD requires a long and exhausting subjective process.

**Objective:** To review the state of the art for the automated autism diagnosis.

**Methods:** In February 2022, we searched multiple databases and several sources of grey literature for eligible studies. We used an adapted version of the QUADAS-2 tool to assess the risk of bias in the studies. A brief report of the methods and results of each study is presented. Data were synthesized for each modality separately using the Split Component Synthesis (SCS) method. We assessed heterogeneity using the I^2^ statistics and evaluated publication bias using trim and fill tests combined with ln DOR. Confidence in cumulative evidence was evaluated using the GRADE approach for diagnostic studies.

**Results:** We included 344 studies from 186020 participants (51129 are estimated to be unique) for nine different modalities in this review, from which 232 reported sufficient data for meta-analysis. The area under the curve was in the range of 0.71-0.90 for all the modalities. The studies on EEG data provided the best accuracy, with the area under the curve ranging between 0.85 to 0.93.

**Conclusions:** The literature is rife with bias and methodological/reporting flaws. Recommendations are provided for future research to provide better studies and fill in the current knowledge gaps.

## Background

### Rationale

Autism spectrum disorder (ASD) represents a panel of conditions that begin during the developmental period and result in impairments of personal, social, academic, or occupational functioning and is currently estimated to affect about 1 in 44 children in the U.S. [1] and 1 in 100 children worldwide [2]. Furthermore, estimates show that about 2.2% of all adults in the U.S. have ASD [3]. It is associated with a high economic burden. The cost of caring for Americans with autism reached $268 billion in 2015 and is estimated to rise to $460 billion by 2025 as individuals with ASD require special healthcare, special education, and government assistance [4]. Individuals with ASD usually have a low quality of life as they have difficulties living independently and having social relationships. They also have problems getting employed as fewer than half of young adults with ASD maintain a job, even lower than the employment rate of ex-convicts who achieve a 75% employment rate [5]. Furthermore, ASD individuals experience substantially higher rates of mood disorders than the general population, contributing to less quality of life and higher mortality through suicide [6].

ASD is associated with verbal and nonverbal communication, social interactions, and behavioral complications [7]. Clinical manifestations typically occur in the 2nd–3rd year of life and persist through adulthood [8]. Evidence suggests that early diagnosis is directly related to a better prognosis [9]. Different health systems follow different clinical pathways; In some countries, children get screened using pre-diagnosis screening methods, and potential cases will be referred to specialized services by general practitioners [7]. In such cases, a lack of training and technical knowledge may lead to a late diagnosis. In some other countries, professionals directly approach the patient and their parents, often without former screening [10], which usually happens when the patient develops significant signs and symptoms.

The diagnosis of ASD requires a long and exhausting subjective process. Clinicians must conduct a clinical assessment of the patient’s developmental age based on a variety of factors (e.g., behavior excesses, communication, self-care, social skills) [7] or a combination of parent/caregiver information, ideally assessed by standardized instrument, with results from standardized direct observation and other additional, independent information from school teachers, partner or public authorities [11]. ASD cannot yet be diagnosed with objectively specific and sensitive biomarkers.

Machine learning provides novel opportunities for human behavior research and clinical translation. With the new advancements in this area, complex, sophisticated predictive models could be quickly developed based on labeled data. It can be applied to the data from individuals with ASD to offer new diagnostic biomarkers. ASD diagnosis could be seen as a classification problem in machine learning where the clinician tries to build a classifier based on data of individuals to classify participants as ASD or typically developed (TD).

Previously, there have been other reviews [12]–[16] with a similar aim, but we believe all those reviews had severe limitations and flaws. First, almost none of the previous studies investigated all the modalities used for this subject. Most of these reviews focused on brain imaging methods, with hardly any mention of other existing methods such as facial recognition models, electroencephalograms, eye tracking systems, etc. Also, their search strategies were seriously flawed. Two reviews [12], [16] used methodological search filters, which are not recommended for diagnostic test accuracy reviews due to the lack of availability of appropriate subject headings, and inconsistent use of those which are available by database indexers [17]. Four other studies [13]–[15] did not report their search strategies or reported them in insufficient detail.

Nevertheless, when comparing the number of studies we found with those reviews, it is evident that the flaws in their search resulted in missing a substantial amount of the available evidence. Additionally, those reviews either did not assess the risk of bias in the included studies or did it ambiguously. Another problem with some of the previous reviews were mixing the diagnostic accuracy results of different assessment modalities into one meta-analysis. This is a classic example of mixing apples and oranges in a meta-analysis, as was stated by Sharpe in 1997 [18]. Other limitations included the short presentation of data from the included studies, ignoring heterogeneity and not investigating its possible sources, not evaluating for the risk of publication biases, and not assessing the overall confidence in the cumulative evidence. Taking all these factors into consideration, this study aims to present a comprehensive, in-depth systematic review (with meta-analyses when feasible) of the specifications and diagnostic accuracy of the current state-of-the-art automated ASD diagnostic or classification models to help future researchers in their efforts and reveal the current gaps in knowledge, pitfalls, and promises.

### Clinical role of the index test

In April 2018, the Food and Drug Administration (FDA) approved the first artificial intelligence (AI)-based system for clinical use. The system, named IDx-DR, was an AI model with a retinal camera device that was developed to detect diabetic retinopathy in adults who have diabetes [19]. Since then, the FDA has evaluated dozens of AI-based health-related models, with at least nine achieving a de novo pathway clearance and three getting pre-market approval [20], [21]. Nevertheless, most algorithms still need to be refined enough to substitute for a clinician’s judgment. It would be preferable for patients and clinicians alike if they had a simple explanation of how a classification algorithm determines a particular label for a case [22].

Thus, when these models are used in a clinical setting, the problem is not all about providing the correct decision, but one should also be able to describe how the model managed to reach that decision. Unfortunately, the decision-making process of most of these algorithms happens in a “black box” manner. There are few justifiable reasons behind those decisions, at least by the current state of clinical knowledge. Other challenges for delivering clinical impact with AI systems include legal issues, logistical difficulties in implementation, and sociocultural considerations [23]. Thus, at the moment, we propose these models should only be considered a low-cost, accessible, non-invasive method for screening children to identify those at risk of developing ASD. So, if enough sensitivity is provided, they may be used as a screening test. Generally, screening tests should provide reasonable sensitivities since test-negatives will not be tested by more specific tests, but they may have lower specificities. Although, if enough specificity is provided, they might be used as add-on tests to ascertain the diagnosis and differentiate it from other conditions with similar symptoms.

### Summary of artificial intelligence model pipelines

Although studies on the diagnosis or classification of clinical outcomes using AI might have developmental details different from one study to another, they usually follow a similar pipeline. This pipeline generally includes (a) data preprocessing and wrangling, (b) feature engineering, (c) feature scaling and selection, and (d) model training and evaluation. This pipeline is presented in **Figure 1**.

**Figure 1.**
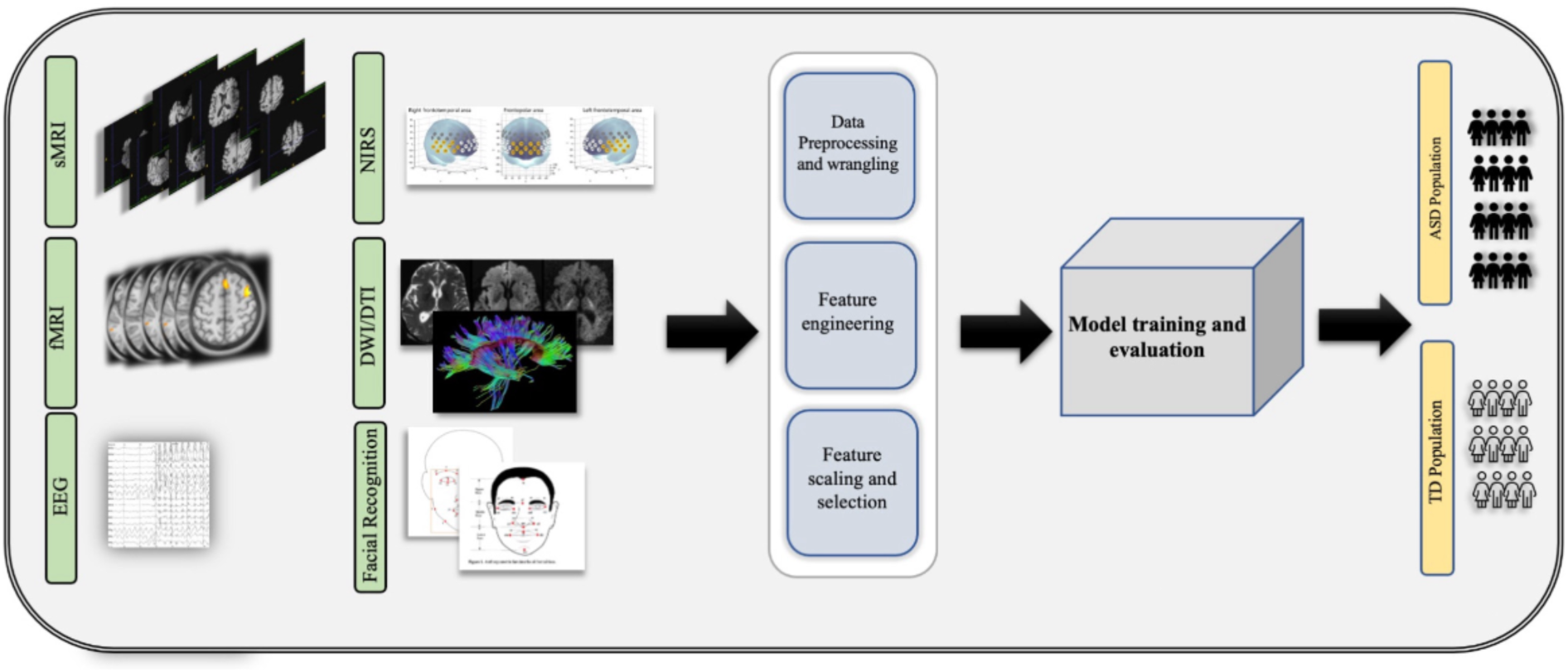
Summary of artificial intelligence model pipelines. DWI/DTI: Diffusion-Weighted Imaging/Diffusion Tensor Imaging, EEG: Electroencephalogram, fMRI: Functional Magnetic Resonance Imaging, NIRS: Near-Infrared Spectroscopy, S-MRI: Structural Magnetic Resonance Imaging.

#### Data preprocessing and wrangling

In health-related tasks, datasets can consist of a variety of modality types. These data are also usually filled with different nuisance sources, which can interfere with optimal model training. Thus, data cleaning is typically necessary to ensure or enhance performance. Although modern deep learning models do not require ideal datasets, preprocessing and wrangling techniques have dramatically increased the model’s performance. Preprocessing techniques usually take advantage of statistical methods to improve data quality in different ways. For instance, in the case of neuroimaging data, slice-timing correction, head-motion correction, and susceptibility distortion correction address particular artifacts, while co-registration and spatial normalization are concerned with signal localization [24].

#### Feature engineering

While data processing includes cleaning and preparing the raw data, feature engineering creates actual model training features. In the case of classifying problems, a feature refers to any measurable property with information about the class ownership extracted from raw data. For instance, measures such as gray matter volume, white matter volume, or mean diffusivity may be used as features in neuroimaging data. The feature engineering process may include various techniques, such as numerical transformations, category encoding, clustering, and principal component analysis [25].

#### Feature scaling and selection

Feature scaling is the process of normalizing the range of independent variables or features. In contrast, feature selection is the selection of a subset of relevant features for use in model development. They shorten training time, improve data compatibility with a learning model class, and benefit the model by avoiding the curse of dimensionality [26]. Feature selection methods are divided into three categories: filtering techniques such as t-test, wrapper methods such as recursive cluster elimination, and embedding methods such as least absolute shrinkage and selection operator (Lasso).

#### Model training and evaluation

The training process is the optimization of model parameters based on the training dataset to find better representations of the corresponding class. This process sometimes leads to overfitting. Overfitting is “the production of an analysis that corresponds too closely or exactly to a particular set of data and may fail to fit additional data or predict future observations reliably” [27]. To test for possible overfitting and evaluate the model’s generalizability, it should be further validated on a dataset different than the one used for the training. Different validation methods exist, such as k-fold cross-validation (CV), leave-one-out cross-validation (LOOCV), and external validation methods [28].

### Objectives

To review the current state-of-the-art automated ASD diagnostic or classification models and present the gaps in knowledge, pitfalls, and promises

## Methods

The design and methods used for this review complied with the Centre for Reviews and Dissemination (CRD’s) Guidance for Undertaking Reviews in Healthcare [29] and are reported in line with Preferred Reporting Items for Systematic Reviews and Meta-Analyses of Diagnostic Test Accuracy (PRISMA-DTA) [30].

### Eligibility criteria

**Population:** individuals with autism spectrum disorder (ASD) regardless of age, sex, and ethnicity

**Index test:** AI models and systems

**Target condition:**

- Autism spectrum disorder (ASD) as defined by the Diagnostic and Statistical Manual of Mental Disorders-IV (DSM-IV), DSM-V, and International Statistical Classification of Diseases and Related Health Problems 11 (ICD-11)
- Childhood autism, atypical autism, and Asperger syndrome as defined by ICD-10
- Autism as defined by Autism Diagnostic Observation Schedule (ADOS), Autism Diagnostic Interview-Revised (ADI-R), The Childhood Autism Rating Scale (CARS), or Gilliam Autism Rating Scale (GARS)
- We did not consider participants at risk for autism as eligible for this review.

**Comparator:** typically developed (TD) individuals

**Reference standard:** diagnosis made by a trained physician, psychiatrist, or other eligible experts based on diagnostic criteria mentioned above

**Study design:** cross-sectional design, including both single-gate (cohort type) and two-gates (case-control type) designs

We assessed the eligibility of studies reports irrespective of their language, publication status, and publication date. We did not include studies that used AI models for the questionnaire-based diagnosis of ASD because those studies are mostly about using machine and deep learning methods to tailor available diagnostic questionnaires, which is different from the scope of this review.

### Information sources

In February 2022, we searched Embase, MEDLINE (Ovid), APA PsycINFO (Ovid), IEEE Xplore, Scopus, and Web of Science Core Collection for eligible studies. Additionally, we searched gray literature using OpenGrey, Center for Research Libraries Online Catalogue (CRL), and Open Access Theses and Dissertations (OATD) to find any unpublished potentially relevant studies. We also carried out a ‘snowball’ search through citation searching (forward and backward citation tracking) using Scopus on all included studies in this review to identify further eligible studies or study reports. As a final step, we checked the references of the reviews with a similar subject identified through our search to see if other potentially eligible studies existed.

Our search strategy is reported in line with the PRISMA search extension [31]. No restriction or search filter was used. Free-text terms and keywords were identified using the MeSH Browser [32] website and PubMed PubReMiner [33] word frequency analysis tool. The search strategy was reviewed using the Peer Review of Electronic Search Strategies (PRESS) [34] guideline. The **Multimedia Appendix 1**: Search strategy file presents a detailed report of the search strategy.

### Study selection

Citations identified from the literature searches and reference list checking were imported to EndNote [35], a citation management software. Duplicates were identified and removed using EndNote’s de-duplication tool. Next, the remaining records were imported into Rayyan QCRI [36], a web-based application that employs natural language processing, artificial intelligence, and machine learning technologies to speed the screening of titles and abstracts of records. Another step of de-duplication was conducted in Rayyan. In this process, duplicates were identified, manually re-checked, and removed using Rayyan’s automatic de-duplication feature, with the similarity threshold set to 0.85. Then four reviewers in two teams independently reviewed the titles and abstracts of the first 50 records. Inter-rater reliability was calculated using Cohen’s kappa to be 0.87, interpreted as almost perfect agreement. Then the same two teams independently screened titles and abstracts of the retrieved records. Two other reviewers were consulted to make the final decision in disagreements. Afterward, the full text of all potentially eligible records was retrieved. Records from the same study were linked to avoiding including data from the same study more than once. Next, the same two teams independently screened full-text studies for inclusion. A study was included when both team reviewers assessed it as satisfying the inclusion criteria. In cases of disagreements, two other reviewers were consulted.

### Data collection process

A data extraction form was developed. The form was pilot-tested by four reviewers using five randomly selected studies. Inter-rater reliability was calculated again using Cohen’s kappa to be 0.93 (almost perfect agreement). After holding discussions to resolve discrepancies, the same reviewers used the form independently to extract data from eligible studies. Extracted data were compared, with any differences being resolved through further discussion. We tried to contact the study authors in cases of missing or unclear data. Extracted data included:

● Study identifiers and design: study name, location, dates, design (single-gate; two-gates), and funding sources.
● Characteristics of dataset: dataset publicity, inclusion, and exclusion criteria, sample size, gender, and age.
● Performance and validation methods: a qualitative description of pre-processing methods, feature extraction, and selection methods, modality parameters, qualitative description of the AI algorithms, validation methods, and reference standard used.
● Model evaluation metrics.

In cases where one study evaluated several algorithms and presented different results, we only extracted the data for the model with the best accuracy.

### Risk of bias

Quality Assessment of Diagnostic Accuracy Studies (QUADAS-2) [37] tool addresses four domains: 1) Patient selection; 2) Index test(s); 3) Reference standard; 4) Flow and timing. Each domain is assessed regarding the risk of bias and concerns regarding applicability to the review question.

Since this review aims to evaluate the diagnostic accuracy of artificial intelligence algorithms, some revisions to the QUADAS-2 were deemed necessary. Thus, some alterations and modifications were done by the discussion and consensus among authors. The patient selection domain in such studies is usually more about data sets for training and testing the model. This domain’s signaling questions and assessments have been revised to cover those issues. The index test was divided into two sections, one evaluating the potential biases in the modality used and the other assessing the AI algorithms in the study. For question eight (validation of the algorithm), judgments were based on the results of a study [38]. In their study, which evaluated the validation methods of studies with the aim of automated autism diagnosis, they found that in small sample sizes (<60), the leave-one-out cross-validation (LOOCV) is the least biased internal validation method.

In contrast, in the studies with a sample size of >60, k-fold cross-validation is the optimal choice. Also, external validation and data-split methods are considered at low risk of validation method bias regardless of the sample size. For the reference standard, we assessed if participants were diagnosed based on valid criteria or by an expert. Finally, the flow and timing domain was judged not to apply to machine learning methodologies. No overall summary score was calculated. A detailed summary of the adopted version of the tool is presented in **Table 1**.

**TABLE 1.** Modified version of the QUADAS-2 tool

|  |
| --- |
| **Domain 1: Patient selection** |
| 1. Are the data sources clear? |
| 2. Were the controls matched with the cases for basic characteristics (age, sex, and IQ)? |
| 3. Did the dataset avoid inappropriate exclusions? |
| **Domain 2: Index test – Modality** |
| 4. Were data acquisition methods judged to be valid? |
| 5. Was the data preprocessing pipeline described in appropriate detail? |
| 6. Was the data preprocessing pipeline judged to be valid? |
| **Domain 3: Index test - AI algorithm** |
| 7. Is there enough clarity about the specification of the algorithms used? |
| 8. Was an appropriate validation and evaluation method used? |
| **Domain 3: Reference standard** |
| 9. Was a valid reference standard test used? |

The tool was piloted before use. Four reviewers independently evaluated 10% of the included studies to calculate inter-rater reliability using Cohen’s kappa, which was 0.78. After re-training and holding discussions to resolve discrepancies, the same four reviewers evaluated all the included studies independently. They recorded supporting information and justifications for judgments of risk of bias for each domain (low, unclear, high, or not applicable). We discussed disagreements and resolved discrepancies through discussion.

### Diagnostic accuracy measures

We used the data from the two-by-two tables to calculate true-positives (TP), true-negatives (TN), false-positives (FP), false-negatives (FN), sensitivity (Se), specificity (Sp), and accuracy for the best-performing algorithm in each study. When any TP/FP/TN/FN values were 0, 0.5 was added to prevent zero cell count. Our primary accuracy measure for the meta-analyses was the diagnostic odds ratio (DOR):

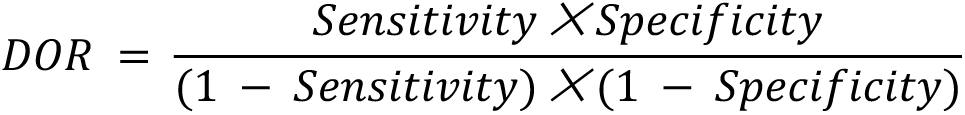

### Synthesis of results

We summarized diagnostic test accuracy by creating a two-by-two table for the best-performing algorithm in each study. When two-by-two tables were unavailable, we tried to calculate the metrics by putting the available data in equations.

Meta-analyses were conducted using R version 4 [39], function ‘SCSmeta’ [40]. Hierarchical bivariate random-effects models are commonly used for the meta-analysis of diagnostic test accuracy studies. These models can consider both within- and between-subject variability and threshold effect [41]. An issue with bivariate models is that the inputs into the model are the study-specific pairs of Se and Sp. The latter can demonstrate heterogeneity across studies either due to systematic differences or implicit dissimilarity in test thresholds, or both. Another issue is that some of the between-study variability could be due to some degree of threshold variability, and while the bivariate approach takes the negative correlation between Se and Sp into account when modeling Se/Sp pairs, such a correlation may also be artifactual because of systematic error (study biases), spectrum effects, or implicit variations in thresholds when tests are interpreted differently. We used the more robust Split Component Synthesis (SCS) method [40]. The SCS estimator for the DOR, Se, and Sp is less biased and has a more minor mean squared error than the bivariate model estimator. For this purpose, first, we estimated the logit Se, logit Sp, and ln DOR for each study. We estimated the summary ln DOR and variance using the IVhet model [42]. Then we generated a centered ln DOR for each study. We fitted two ordinary least squares regression models: one for the logit Se (dependent variable) on the centered DOR (independent variable) and the other for the logit Sp (dependent variable) on the centered DOR (independent variable). The intercept in each linear regression model corresponds to the summary logit Se and summary logit Sp respectively.

Summary Receiver Operating Characteristics (SROC) curves were generated for each modality based on parameter estimates extracted from the SCS model. The SROC curves were specified by the summary Se and Sp intersection point, its 95% CIs, and the confidence limits of the Se and 1-Sp. Individual study Se/Sp pairs were indicated on the plot as circles with size proportional to the inverse of the study’s variance in DOR. The area under the curve (AUC) was calculated using the following formula:

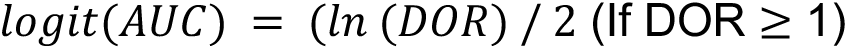

We quantified heterogeneity with I^2^ and τ^2^ statistics. I^2^ quantifies inconsistency across studies to assess the impact of heterogeneity on the meta-analysis [43]. I^2^ is interpreted as follows: 0% to 40%: might not be important; 30% to 60%: may represent moderate heterogeneity; 50% to 90%: may represent substantial heterogeneity; 75% to 100%: represents considerable heterogeneity.

To investigate the possible role of overfitting on the accuracy results of the included studies, we performed a meta-regression analysis. In this analysis, the accuracy of each study was plotted against its sample size. We also conducted sensitivity analyses on modalities with at least five studies at low risk of bias in all domains to evaluate the robustness of our results.

### Reporting bias assessment

The impact of publication bias has been understudied in the context of DTA systematic reviews. One possible reason is that the statistical significance of treatment efficacy is well-defined in clinical trials; however, statistical significance needs to be more intuitively defined for measures of diagnostic test accuracy. Another reason is that the odds ratio is expected to be large in diagnostic studies. Applying tests for funnel plot asymmetry in such circumstances will likely result in publication bias being incorrectly indicated by the test far too often [44]. Nevertheless, empirical evidence has demonstrated that smaller studies report greater diagnostic test accuracy; thus, ignoring the impact of publication bias when conducting a meta-analysis can lead to overestimating the diagnostic test accuracy [45].

A contour-enhanced funnel plot was designed to assess publication bias in our review with ln DOR on the x-axis and the inverse of the standard error on the y-axis. To check for potential unpublished studies on the plot, we used the trim and fill method [46] combined with ln DOR, as shown in a simulation study, to be superior to other funnel plot asymmetry tests for DTA reviews [47]. Using the Deeks method, we also performed a statistical test for funnel plot asymmetry [48].

### Confidence in cumulative evidence

The confidence in cumulative evidence for each synthesis was assessed using the Grading of Recommendations, Assessment, Development, and Evaluation (GRADE) approach for diagnostic tests and strategies [49], which takes into account five considerations: study design, risk of bias, inconsistency, indirectness, imprecision, and publication bias. Factors designed for upgrading the evidence (dose effect, plausible bias, large effect, and confounding) still need to be well developed, and thus we did not consider them in our review. The only exception is the ‘large effect’ domain, which was considered in this review. We decided that a substantial likelihood of disease associated with test results (sensitivity and specificity more than 0.85) will increase the quality of evidence. Four reviewers rated the certainty of the evidence for the outcome as high, moderate, low, or very low. We resolved any discrepancies by consensus.

## Results

### Overview of results

This research aimed to review the current state-of-the-art automated ASD diagnostic or classification models and present the gaps in knowledge, pitfalls, and promises. First, we have introduced a summary of the study selection process, followed by the characteristics of the included studies, including sample sizes, participants’ age, the modalities used, the sources of data (datasets), the classifiers implemented, and the evaluation metrics utilized by the included studies. Next, we have presented the results for assessing the risk of bias in the studies. It is immediately followed by the results of the syntheses, which include synthesis based on modality, synthesis based on feature set, meta-regression analysis, and sensitivity analysis. Finally, we have evaluated the reporting biases in the included studies and confidence in the cumulative evidence. A summary overview of the results, alongside a chart of the flow of the study, is presented in **Figure 2**.

**Figure 2.**
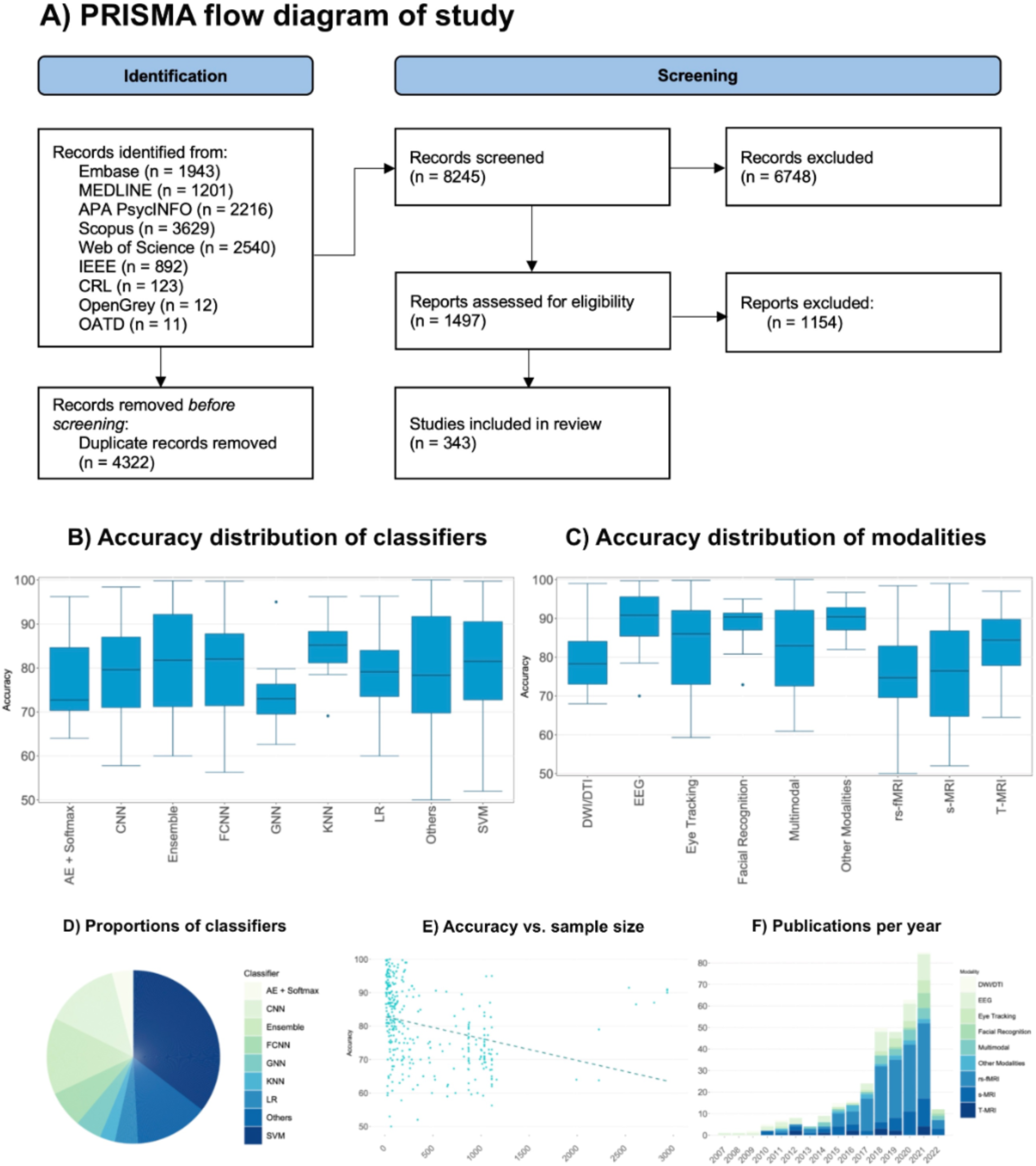
A) PRISMA flowchart of the review; B) Accuracy distribution of classifiers in this review; C) Accuracy distribution of modalities in this review; D) Proportions of classifiers in this review; E) Meta-regression of accuracy vs. sample size of the included studies; F) Publications per year. AE: Autoencoders, CNN: Convolutional neural networks, FCNN: Fully-Connected Neural Networks, GNN: Graph Convolutional Networks, KNN: K-Nearest Neighbors, LR: Logistic Regression, SVM: Support Vector Machines. DWI/DTI: Diffusion-Weighted Imaging/Diffusion-Tensor Imaging, EEG: Electroencephalography, rs-fMRI: Resting-state functional Magnetic Resonance Imaging, S-MRI: Structural Magnetic Resonance Imaging, T-fMRI: Task-based functional Magnetic Resonance Imaging.

### Study selection

For a detailed summary of the flow of the studies, see the PRISMA flow diagram presented in **Figure 2**. For this review, we identified 12567 records in our primary search. After removing duplicates, we screened the titles and abstracts of 8245 records. Another 6748 records were excluded at that stage, and 1497 records remained for full-text assessment. We excluded 1154 studies following the evaluation of the full text of the records. In the end, we included 344 studies [50]–[393] with 186020 participants (following the removal of overlapping participants in public databases, 51129 are estimated to be unique) in this review. Three studies reported results for two different modalities separately, resulting in 347 results being included in this review.

### Study characteristics

A summary of the characteristics of the contributing studies is provided in **Multimedia Appendix 2: Characteristics of included studies** which includes the parcellation atlas used by each study (if applicable), the feature selection methods used, the sample sizes, the validation method for the algorithm, the classifier and the algorithm specifications, the evaluation metrics of the algorithm for diagnosing or classifying ASD individuals, and the potential sources of bias in each study. Readers who are interested in designing new AI algorithms for diagnosing or classifying ASD individuals are highly recommended to check this data as it would give them an overview of the methods used so far and their effectiveness, guiding them through the next steps required to achieve better results, while also avoiding duplicate efforts.

#### Sample sizes

The median sample size was 152.5 participants (interquartile range 58–871). The smallest sample size was 12, and the largest was 17614.

#### Age of participants

The age group of participants overlapped in most of the included studies. Thirteen studies included toddlers (<2 years old), 143 included children and adolescents (2-18 years old), 30 included adults (>18 years old), and 139 included participants of all ages. Also, 23 studies did not report the age of the participants.

#### Modality

Our investigations revealed that 155 studies used the data from resting-state functional magnetic resonance imaging (rs-fMRI), 53 used structural MRI (S-MRI), 48 used electroencephalography (EEG), 21 used eye-tracking, 14 used task-based fMRI (T-fMRI), 11 used diffusive weighted imaging/diffusive tensor imaging (DWI/DTI), and 10 used facial recognition. Also, 24 studies used data from more than one modality. In contrast, 14 used other miscellaneous modalities (functional near-infrared spectroscopy, kinematic features, response to name, video analysis, photo shooting, vocal analysis, and positron emission tomography).

#### Dataset

We observed that 187 studies used the data stored in Autism Brain Imaging Data Exchange (ABIDE) initiative [394], [395] (a dataset that has aggregated functional and structural brain imaging data of ASD participants collected from laboratories around the world), 15 used the data from the National Database for Autism Research (NDAR) [396] (a dataset with data at all levels of biological and behavioral organization (molecules, genes, neural tissue, behavioral, social and environmental interactions) of ASD participants), and 141 studies used other sources.

#### Classifier

Our investigations revealed that 124 studies used a support vector machine (SVM) classifier, 48 used convolutional neural networks (CNN), 24 used fully-connected neural networks (FCNN), 17 used graph neural networks (GNN), 15 used logistic regression (LR), 14 used a combination of an autoencoder and softmax layer (AE + Softmax), and ten used k-nearest neighbors model (KNN). Also, 50 studies used ensemble algorithms, while 48 used other miscellaneous models. The choice of classifiers is usually highly dependent on the data structure. As a result, we sought to identify the type of classifiers used for each modality. Check **Figure 3** for the results.

**Figure 3.**
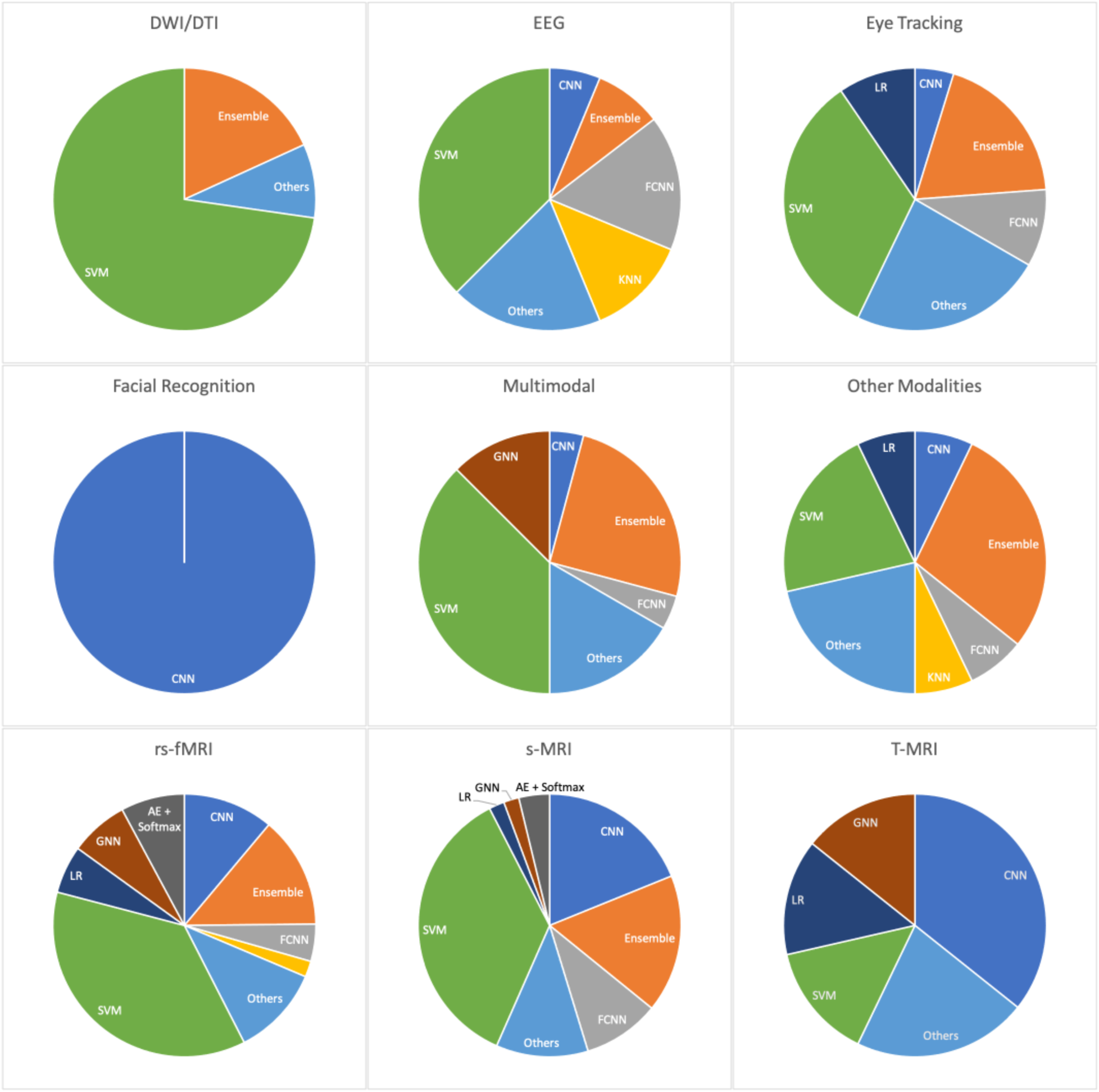
The type of classifiers used for each modality. AE: Autoencoders, CNN: Convolutional neural networks, FCNN: Fully-Connected Neural Networks, GNN: Graph Convolutional Networks, KNN: K-Nearest Neighbors, LR: Logistic Regression, SVM: Support Vector Machines. DWI/DTI: Diffusion-Weighted Imaging/Diffusion-Tensor Imaging, EEG: Electroencephalography, rs-fMRI: Resting-state functional Magnetic Resonance Imaging, S-MRI: Structural Magnetic Resonance Imaging, T-fMRI: Task-based functional Magnetic Resonance Imaging.

Most of the included modalities were classified using SVMs as they are easy to use and powerful conventional classifiers. The only exceptions were the facial recognition modality (which almost entirely used CNNs) and the T-MRI modality (which mainly used CNNs).

#### Evaluation metrics

The median Sensitivity was 80.8% (interquartile range 71.2%-89.7%). The lowest sensitivity was 24.8%, and the highest was 100%. Unfortunately, 116 studies did not report the sensitivity.

The median Specificity was 80.9% (interquartile range 71.0%-90.6%). The lowest specificity was 33.0%, and the highest was 100%. Unfortunately, 117 studies did not report the specificity.

The median accuracy was 80.0% (interquartile range 70.9%-90.0%). The lowest accuracy was 50.0%, and the highest accuracy was 100%.

### Risk of bias

**Figure 4** shows the judgments for risk of bias and concerns regarding applicability for each domain across the included studies.

**Figure 4.**
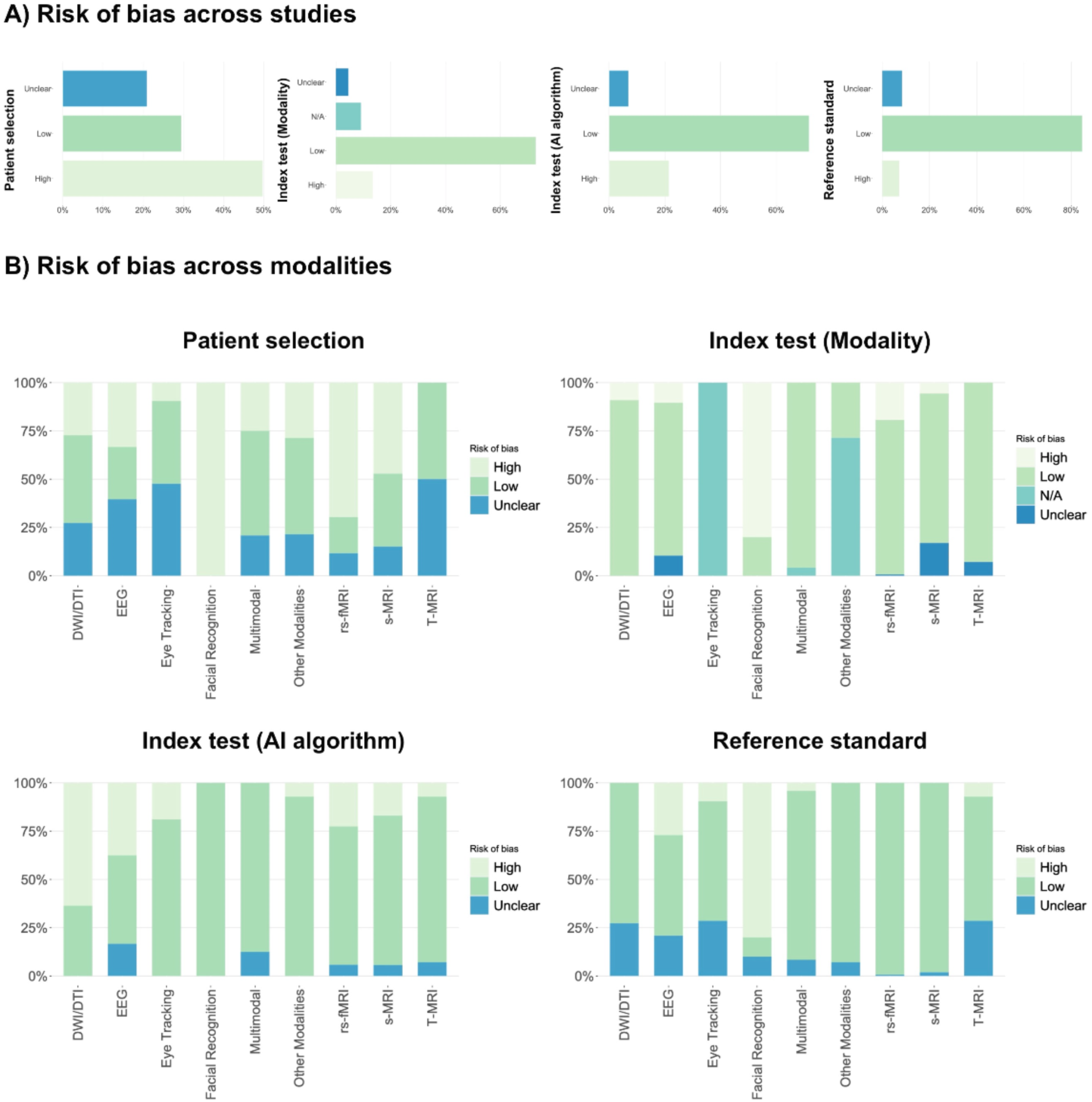
Risk of bias graph: review authors’ judgments about each risk of bias and applicability item presented as percentages across all included studies. DWI/DTI: Diffusion-Weighted Imaging/Diffusion-Tensor Imaging, EEG: Electroencephalography, rs-fMRI: Resting-state functional Magnetic Resonance Imaging, S-MRI: Structural Magnetic Resonance Imaging, T-fMRI: Task-based functional Magnetic Resonance Imaging.

#### Patient Selection

For this domain, 174 studies (51%) were judged to be at high risk of bias, while 73 (21%) were at unclear risk. Only 96 studies (28%) were considered to be at low risk of bias in patient selection. The three main issues with studies at high/unclear risk were:

1. Unclear data source and/or participants’ characteristics (age, sex, etc.)
2. ASD and TD groups not being matched for age or sex
3. Unclear inclusion/exclusion criteria

#### Index Test - Modality

For this domain, 47 studies (14%) were judged to be at high risk of bias, while 16 studies (5%) were considered to be at unclear risk. Also, this domain was not applicable (N/A) for 32 studies (9%). Most facial recognition studies were at high risk for this domain due to not performing data preprocessing/cleaning.

#### Index Test - AI Algorithm

For this domain, 75 studies (22%) were judged to be at high risk of bias, primarily due to inappropriate validation methods. In contrast, 24 (7%) were considered to be at unclear risk due to insufficient reports of the characteristics of the classifying algorithm used.

#### Reference Standard

For this domain, 25 studies (7%) were judged to be at high risk of bias because they did not specify if participants were diagnosed using valid criteria or with the help of an expert. Also, 29 studies (8%) mentioned that an expert diagnosed participants but did not specify which criteria were used for the diagnosis.

### Results of syntheses

#### Synthesis based on modality

Two hundred thirty-two studies provided sufficient data (sensitivity and specificity values or other diagnostic measures which allowed us to calculate them ourselves) for SCS analysis. Results of the meta-analyses are presented in SROC curves in **Figure 5** and **Table 2**.

**Figure 5.**
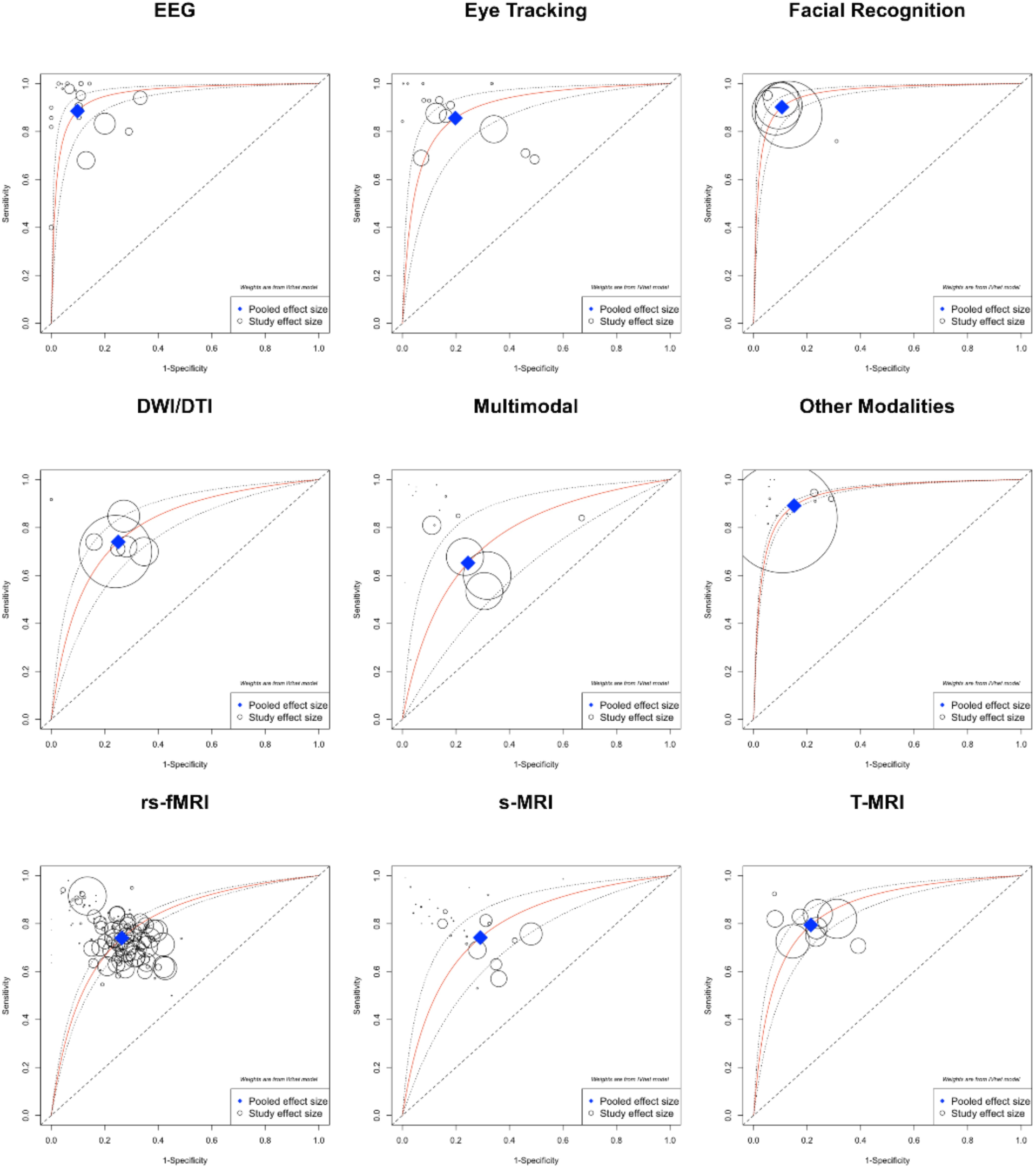
Summary Receiver Operating Characteristics (SROC) curves for SCS analysis of 232 included studies. The confidence limits of the DOR are also shown in the plot. Individual study Se/Sp pairs are indicated on the plot as open circles with size proportional to the inverse of the variance of the study’s logarithm of DOR. For better visualization purposes, the size proportions of open circles are different between graphs.

**TABLE 2.** Results of SCS meta-analyses for each modality.

| Modality | # Studies | N (ASD/TD) | Sen (95% CI) | Spe (95% CI) | AUC (95% CI) | +LR (95% CI) | -LR (95% CI) | I<sup>2</sup> | Tau<sup>2</sup> |
| --- | --- | --- | --- | --- | --- | --- | --- | --- | --- |
| rs-fMRI | 115 | 34283/32462 | 0.74 (0.71-0.77) | 0.74 (0.71-0.76) | 0.74 (0.72-0.76) | 2.80 (0.38-20.50) | 0.35 (0.05-2.61) | 94 | 0.74 |
| S-MRI | 30 | 4114/4667 | 0.74 (0.64-0.83) | 0.71 (0.60-0.80) | 0.73 (0.65-0.79) | 2.55 (0.38-16.95) | 0.36 (0.05-2.61) | 92 | 1.21 |
| EEG | 21 | 688/840 | 0.89 (0.82-0.93) | 0.90 (0.84-0.94) | 0.89 (0.85-0.93) | 9.06 (0.49-166.29) | 0.13 (0.01-2.12) | 41 | 1.08 |
| DWI/DTI | 7 | 379/387 | 0.74 (0.66-0.81) | 0.75 (0.67-0.82) | 0.74 (0.69-0.80) | 2.96 (0.38-22.76) | 0.35 (0.05-2.61) | 50 | 0.24 |
| Eye Tracking | 15 | 433/414 | 0.86 (0.76-0.92) | 0.80 (0.69-0.88) | 0.83 (0.76-0.88) | 4.33 (0.40-46.41) | 0.18 (0.01-2.34) | 66 | 1.34 |
| Facial Recognition | 6 | 6378/6368 | 0.90 (0.85-0.94) | 0.89 (0.84-0.93) | 0.90 (0.87-0.92) | 8.53 (0.48-150.37) | 0.11 (0.01-2.02) | 95 | 0.43 |
| Multimodal | 16 | 2354/2531 | 0.65 (0.48-0.79) | 0.76 (0.59-0.87) | 0.71 (0.59-0.80) | 2.66 (0.38-18.54) | 0.46 (0.08-2.59) | 93 | 1.27 |
| Other Modalities | 13 | 1758/1679 | 0.89 (0.88-0.91) | 0.85 (0.83-0.87) | 0.87 (0.86-0.88) | 5.89 (0.43-80.2) | 0.13 (0.01-2.13) | 0 | -0.06 |
| T-fMRI | 9 | 375/311 | 0.79 (0.73-0.85) | 0.79 (0.72-0.84) | 0.79 (0.75-0.83) | 3.72 (0.39-35.09) | 0.26 (0.03-2.53) | 32 | 0.17 |

#### Synthesis based on the feature set

In this synthesis, 232 studies that provided sensitivity and specificity were included. We performed statistical synthesis for the feature set used in the studies if there were at least five studies for the feature set. Data were presented as accuracy range in cases with fewer than five studies. See **Multimedia Appendix 3: Results of SCS meta-analyses** for the results of the syntheses.

#### Results of meta-regression analysis

All studies were included in the meta-regression analysis to investigate the effect of sample size on the reported accuracy measures. The results of this synthesis are presented in **Figure 1**. β_1_ was -0.002, indicating a negative correlation between the sample size of the studies and their accuracy result. This relationship was tested using a t-test, revealing that the correlation was statistically significant (p<.001). β_0_ was 81.0, while R^2^ was 0.003.

#### Results of sensitivity analyses

Thirty-six studies (5 modalities) provided sufficient data to be included in the sensitivity analyses. The results of the sensitivity analyses are presented in **Table 3**.

**TABLE 3.** Results of sensitivity analyses.

| Modality | # Studies | N (ASD/TD) | Sen (95% CI) | Spe (95% CI) | AUC (95% CI) | +LR (95% CI) | -LR (95% CI) | I<sup>2</sup> | Tau<sup>2</sup> |
| --- | --- | --- | --- | --- | --- | --- | --- | --- | --- |
| rs-fMRI | 8 | 602/566 | 0.74 (0.54-0.88) | 0.70 (0.50-0.85) | 0.72 (0.58-0.83) | 2.50 (0.38-16.34) | 0.37 (0.05-2.61) | 89 | 1.50 |
| S-MRI | 10 | 982/1420 | 0.86 (0.74-0.93) | 0.84 (0.71-0.92) | 0.85 (0.77-0.91) | 5.34 (0.42-67.55) | 0.16 (0.01-2.29) | 83 | 1.08 |
| Eye Tracking | 5 | 104/106 | 0.84 (0.57-0.95) | 0.82 (0.54-0.95) | 0.83 (0.65-0.93) | 4.68 (0.41-53.44) | 0.20 (0.02-2.4) | 73 | 2.75 |
| Multimodal | 8 | 405/373 | 0.82 (0.71-0.90) | 0.90 (0.81-0.95) | 0.87 (0.80-0.92) | 8.42 (0.48-147.26) | 0.19 (0.02-2.39) | 53 | 0.79 |
| Other Modalities | 5 | 124/132 | 0.91 (0.84-0.95) | 0.89 (0.81-0.94) | 0.90 (0.85-0.93) | 8.33 (0.48-144.44) | 0.10 (0.01-1.97) | 0 | -0.41 |

### Reporting biases

The contour-enhanced funnel plot for the studies that reported both sensitivity and specificity is presented in **Figure 6**.

**Figure 6.**
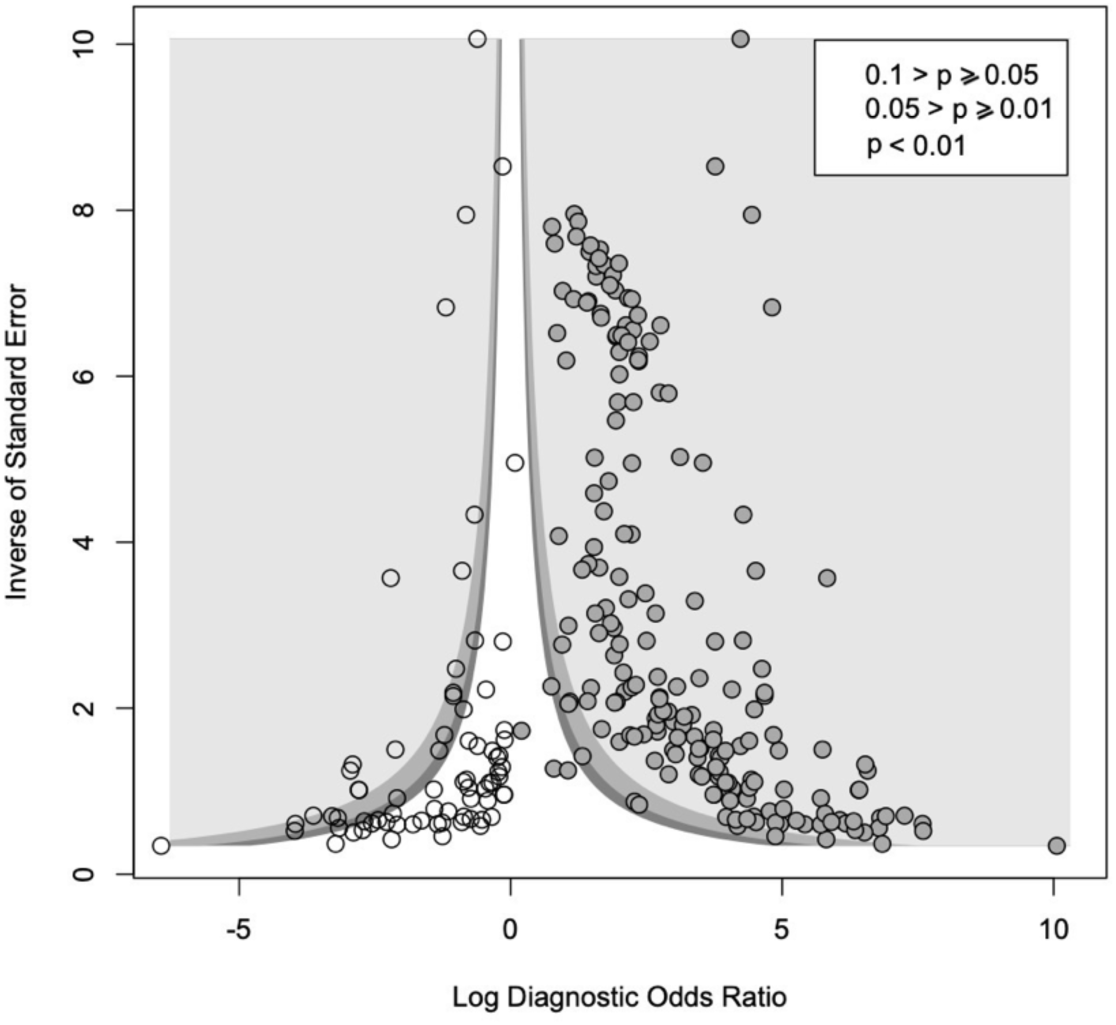
Funnel plot of the included studies with the trim-and-fill method applied. Solid points indicate published studies, while white points indicate unpublished studies.

Visual inspection of the plot revealed an asymmetrical plot, with missing studies in the lower-left portion of the plot. Though the statistical test (p=.115) was not significant, it cannot rule out the presence of publication bias.

### Confidence in cumulative evidence

Confidence in cumulative evidence was judged to be low for rs-fMRI, very low for S-MRI, high for EEG, moderate for eye tracking, moderate for T-fMRI, low for DWI/DTI, low for facial recognition, high for other modalities, and very low for multimodal results.

## Discussion

### Summary of main findings

This research offers the most extensive review of the automated diagnosis or classification of ASD up to this date. In this systematic review, we screened 12567 records and included 344 studies. For a summary report of the results, see **Table 4**.

**TABLE 4.** Summary of main findings.

| Modality | Number of studies | Sample size | Pooled measures* (95% CI) |  | AUC (95% CI) | Heterogeneity (I<sup>2</sup>) | Confidence in cumulative evidence (GRADE) |
| --- | --- | --- | --- | --- | --- | --- | --- |
|  |  |  | Sensitivity | Specificity |  |  |  |
| rs-fMRI | 115 | 66745 | 0.74 (0.71-0.77) | 0.74 (0.71-0.76) | 0.74 (0.72-0.76) | 94% | ⊗⊗⊕⊕ |
| S-MRI | 30 | 8781 | 0.74 (0.64-0.83) | 0.71 (0.60-0.80) | 0.73 (0.65-0.79) | 92% | ⊗⊗⊗⊕ |
| EEG | 21 | 1528 | 0.89 (0.82-0.93) | 0.90 (0.84-0.94) | 0.89 (0.85-0.93) | 41% | ⊕⊕⊕⊕ |
| Eye tracking | 15 | 847 | 0.86 (0.76-0.92) | 0.80 (0.69-0.88) | 0.83 (0.76-0.88) | 66% | ⊗⊕⊕⊕ |
| T-fMRI | 9 | 686 | 0.79 (0.73-0.85) | 0.79 (0.72-0.84) | 0.79 (0.75-0.83) | 32% | ⊗⊕⊕⊕ |
| DWI/DTI | 7 | 766 | 0.74 (0.66-0.81) | 0.75 (0.67-0.82) | 0.74 (0.69-0.80) | 50% | ⊗⊗⊕⊕ |
| Facial recognition | 6 | 12746 | 0.90 (0.85-0.94) | 0.89 (0.84-0.93) | 0.90 (0.87-0.92) | 95% | ⊗⊗⊕⊕ |
| Other modalities | 13 | 3437 | 0.89 (0.88-0.91) | 0.85 (0.83-0.87) | 0.87 (0.86-0.88) | 0% | ⊕⊕⊕⊕ |
| Multimodal | 16 | 4885 | 0.65 (0.48-0.79) | 0.76 (0.59-0.87) | 0.71 (0.59-0.80) | 93% | ⊗⊗⊗⊕ |
| * Estimated based on split component synthesis (SCS) meta-analysis. |  |  |  |  |  |  |  |
| ** ⊗⊗⊗⊕: Very low certainty, ⊗⊗⊕⊕: Low certainty, ⊗⊕⊕⊕: Moderate certainty, ⊕⊕⊕⊕: High certainty |  |  |  |  |  |  |  |

As our results suggest, EEG data may be the most reliable source for establishing models for automated diagnosis or classification of ASD. However, it should be noted that most of the included studies on EEG used in-house datasets, which are usually more homogenous in their core compared to the studies on other modalities such as rs-fMRI and S-MRI, where most studies used the more heterogenous publicly available datasets like ABIDE. Nevertheless, we encourage future research to work more on EEG data, considering the limited number of studies currently available for this modality. Unfortunately, at the moment, no large publicly available dataset exists for EEG of ASD patients, which in turn makes future research on this modality less feasible.

Another finding observed in our results was the high accuracy of the studies that utilized facial recognition models. Although the results of these studies look promising, one should be careful in interpreting these results because most of the studies on this modality used the Kaggle dataset, where most of the images are downloaded from websites and Facebook pages related to autism and thus, might not be considered a very reliable source of data.

We also found other interesting results across the included studies. These results are presented in **Highlight box 1**.

#### HIGHLIGHT BOX 1 |

Results of highlighted research. DNN: Deep Neural Network, DWI/DTI: Diffusion-Weighted Imaging/Diffusion Tensor Imaging, EEG: Electroencephalography, FC: Functional Connectivity, FS: Feature Selection, ROI: Region of Interest, rs-fMRI: Resting-state functional Magnetic Resonance Imaging, S-MRI: Structural Magnetic Resonance Imaging.

The study of A Ronicko 2020 [85] found that the classification of ASD using rs-fMRI data improves with full correlation FC compared to partial correlation.

The study of Abdolzadegan 2020 [219] on EEG data found that the use of the DBSCAN algorithm for artifact removal significantly improves classification results.

The study of Ahmed 2020 [199] proposed a single-volume image generator that can produce 2D three-channel images from a 4D fMRI image. They proposed that the generated 2D images represent activated brain regions during the performed task by the patients promptly.

The study of Aslam 2021 [252] proposed an EEG-based ASD classification processor that targets a patch-form factor sensor that may be used for long time monitoring in a wearable environment.

The study of Cancino 2021 [103] evaluated the impact on the performance of a given model when using different preprocessing strategies on rs-fMRI, namely global signal regression (GSR), bandpass filtering (BPF), BPF + GSR, and none. They concluded that the best preprocessing strategy depends on the classification model used.

The study of Demirhan 2018 [311] concluded that when using S-MRI data, the minimal-Redundancy Maximal-Relevance (mRMR) algorithm was more successful in FS than the ReliefF algorithm when the selected number of the features was the same.

The study of Eill 2019 [381] found that rs-fMRI data was significantly more informative than S-MRI and DWI/DTI data for the classification of ASD and non-ASD participants.

The study of Fan 2021 [302] proposed a novel federated deep learning framework for multi-site 3D brain MRI images that aggregates learned features from different sites without transferring raw data, ensuring the security of subject information, while also holding to high accuracy performance.

The study of Ferrari 2020 [329] introduced a new workflow to deal with confounders and outliers in medical data which allows for finding generalizable patterns even if the dataset is limited.

The study of Georges 2020 [341] investigated a large pool of existing FS techniques for boosting feature reproducibility within a dataset. They introduced FS-Select, a method capable of identifying the best FS method to discover the most reproducible and reliable subset of features.

The study of Graa 2019 [307] introduced a multi-view learning-based data proliferator that enables the classification of imbalanced multi-view representations by generating synthetic data for each view to handle imbalanced data and mapping all original and proliferated views into a shared subspace where their distributions are aligned for the target classification task.

The study of Gupta 2020 [129] proposed a new measure called ambivert degree that considers the node’s degree as well as connection weights in rs-fMRI data to identify nodes with both high degree and high connection weights as hubs, leading to significantly higher classification accuracy with significantly fewer trainable weights compared to using FC features.

In the study of Hu 2020 [102], an interpreting method was proposed which could explain a trained FCNN model with a precise linear formula for each input instance and identify decision features of the model which contributed most to the classification task.

In the study of Huang 2019 [65], a novel sparse low-rank constrained multi-templates data-based method for ASD diagnosis or classification using rs-fMRI data was proposed, which simultaneously performs FS and adaptive local structure learning.

In the study of Huang 2019 [71], novel multiple network-based frameworks for rs-fMRI modality were introduced to enhance the representation of FC networks by fusing the common and complementary information conveyed in multiple networks.

The study of Huang 2021 [174] found that using causal connectivities for FC data improves diagnostic accuracy significantly compared with using correlations and partial correlations.

The study of Jiao 2020 [87] utilized capsule networks to build classifiers for classifying ASD participants and stratifying them into groups with distinct FC patterns.

In the study of Jun 2019 [162], a novel method was proposed that directly models the regional temporal BOLD fluctuations in a stochastic manner and estimates the dynamic characteristics in the form of likelihoods. They also transformed the learned weight coefficients of the model into activation patterns, from which it was possible to identify the ROIs that are highly associated with ASD and TD groups.

In the study of Karampasi 2020 [193], they compared classification performance using different FS methods, namely Local Learning-based Clustering FS (LLCFS), Infinite FS (InfFS), and minimal-Redundancy Maximal-Relevance (mRMR) and Laplacian Score. They concluded that the best FS method depends highly on the classification model. They also introduced two novel features, namely the Haralick texture features and the Kullback-Leibler divergence.

In the study of Kazeminejad 2020 [150], graph theory was used to extract the positive correlation matrix (only the positive values of the original correlation matrix), the absolute value of the correlation matrix, and the anticorrelation matrix (only the negative correlation values). Following implementing a model on those data, they concluded that graph features extracted from the anti-correlation matrix led to the highest accuracy, suggesting that anti-correlation should not be discarded as they may include useful information that would aid the classification task.

The study of Kernbach 2018 [108] proposed a transdiagnostic hierarchical Bayesian modeling framework for rs-fMRI data which combined Indian Buffet Processes and Latent Dirichlet Allocation to find shared endo-phenotypes of default mode dysfunction in attention deficit hyperactivity disorder (ADHD) and ASD.

The study of Leming 2021 [301] introduces a novel technique of deriving symmetric similarity matrices from regional histograms of grey matter volumes estimated from S-MRI as features.

In Li 2017 (363) study, a new approach for estimating FCs by remodeling Pearson’s correlations as an optimization problem was suggested, which provided a way to incorporate biological/physical priors into the FCs.

The study of Li 2020 [62] proposed a federated learning approach, where a decentralized iterative optimization algorithm is implemented and shared local model weights are altered by a randomization mechanism, ensuring that private information cannot be recovered from the model gradients or weights.

The study of Li 2021 [346] designed novel ROI-aware graph convolutional layers for fMRI data, which contained ROI-selection pooling layers that highlight salient ROIs making it possible to infer which ROIs were important for prediction.

The study of Lu 2021 [391] for the first time integrated genomic data with rs-fMRI data for the classification of ASD.

The study of Ma 2021 [159] found that phase synchrony-based classification models of fMRI data outperformed static FC-based models.

The study of Manoharan 2021 [222] on EEG data found that cosine metric and FAN (fixed amount of nearest neighbors) outperformed Euclidean metric and threshold respectively as the distance metric and the neighborhood selection strategy.

The study of Naghashzadeh 2021 [156] exploited the transfer function perturbation (TFP) method to estimate the instantaneous phase and envelope features from rs-fMRI data for the classification task. They concluded that phase features had a significantly lower correlation than envelope features.

The study of Okamoto 2021 [111] implemented a model based on invariant information clustering (IIC) which improved the performance of the leave-one-site-out cross-validation technique.

In the study of Payabvash 2019 [387], tract-based spatial statistics (TBSS) were used for voxel-wise comparison and co-registration of edge density (ED) maps in addition to conventional DTI metrics, resulting in an improvement of model predictive power.

The study of Rabany 2019 [173] utilized dynamic FC measures from rs-fMRI data in a model to classify and study the overlap in neuropathology between schizophrenia and ASD.

The study of Reiter 2021 [120] used different ASD sample compositions stratified by gender and severity score for the classification task. Their findings suggested that model performance varies significantly with the sample composition.

The study of Sadiq 2022 [196] used the non-oscillatory brain connectivity (NOC) method instead of Pearson’s correlation coefficients (PCC) to distinguish subtypes of ASD (namely autistic disorder, Asperger’s disorder, and Pervasive developmental disorder-not other specified) from healthy controls. They found that the use of NOC measures significantly improved the performance of the model compared to using PCC.

In the study of Sartipi 2018 [186], ROIs from rs-fMRI data were decomposed using the double-density dual-tree discrete wavelet transform into time-frequency sub-bands and the generalized autoregressive conditional heteroscedasticity (GARCH) model was used for feature extraction from those sub-bands. Extracted features were used in a classification model and achieved satisfactory results.

The study of Shahamat 2020 [315] proposed a novel genetic algorithm-based brain masking (GABM) method for visualization of the knowledgeable regions used by a 3D-CNN trained on the fMRI FC data.

The study of Shi 2020 [128] proposed a novel FS method based on the minimum spanning tree (MST) to seek neuromarkers from rs-fMRI data, which significantly improved the performance of the classification task.

The study of Soussia 2018 [340] used high- and low-order relationships between morphological regions in the S-MRI data as features instead of the typical brain connectomes and found that they improve classification performance.

In the study of Xing 2018 [112], a new convolutional neural network with element-wise filters (CNN-EW) for FC data was proposed that gives a unique weight to each edge of the brain network which may reflect the topological structure information more realistically.

The study of Yang 2019 [86] implemented several classical machine learning classifiers (such as support vector machines, logistic regression, and ridge) on seven different brain atlases (CC400, CC200, AAL, HOA, EZ, TT, and Dosenbach) for rs-fMRI data and found that the most promising brain atlas was Craddock 400 (CC400).

The study of Yang 2020 [81] implemented DNNs with four different hidden layer configurations using the four different pipeline datasets from the ABIDE repository (CPAC, CCS, DPARSF, and NIAK). Their results indicated that the dataset preprocessed by using the CPAC (Configurable Pipeline for the Analysis of Connectomes) pipeline achieved the highest accuracy, recall, and precision.

In the study of Yang 2021 [337], three new models (2D CAM, 3D CAM, and 3D Grad-CAM) were proposed which help with the interpretability of the classification task on S-MRI data.

The study of Yao 2019 [144] proposed a multi-scale triplet graph convolutional network (MTGCN) for rs-fMRI data that utilizes multiple atlases to partition the brain into ROIs and also takes into account the underlying high-order (e.g., triplet) association among subjects.

In the study of Zhang 2020 [202] fixel-based analysis (FBA) method was implemented on DTI data for the first time to classify ASD participants.

The study of Zhang 2020 [334] introduced path signature (PS) features for rs-fMRI data, which can capture the dynamic longitudinal information of the brain development for ASD Identification.

Study of Zhao 2018 [182] proposed a multi-level, high-order FC network representation as an alternative to the pairwise correlation between ROIs in fMRI data that can capture complex interactions among brain regions.

In the study of Zhao 2020 [158], a central-moment method was proposed to extract temporal-invariance properties contained in either low- or high-order dynamic FC networks.

In the study of Zhuang 2019 [89], invertible networks were used to help interpret the classification model’s decisions on fMRI data, presenting new insights into model interpretation techniques.

In general, we believe state-of-the-art AI algorithms might be used to ascertain diagnosis by differentiating ASD from other conditions with similar symptoms. These tools might also be used as a screening test before the actual diagnosis by the clinician. Yet again, we do not suggest using these tools as the replacement for a clinician’s judgment, as stakeholders would prefer a simple explanation for how an algorithm arrives at its classification of a particular case. Unfortunately, the decision-making process of these algorithms happens in a “black box” manner. We found little effort in the included studies to explain the features their algorithms specifically used to get to their decisions and the justifiable reasons behind them. Thus, we suggest future studies try to present saliency maps or class activation maps (CAM) to visualize the features used by their models to classify ASD individuals and to what extent those decisions are in line with the available clinical knowledge. Some other methods to address this issue are provided elsewhere [397]–[402]. The inappropriate use of feature reduction methods in the literature was also an issue. It must be noted that any feature reduction steps should not be applied to test data as they may induce overfitting [403]. Very few studies seem to consider this issue. Some other critical challenges of using automated tools to achieve clinical impact are highlighted in the paper of Kelly et al. in 2019 [23]. Regrettably, very few attempts to address these challenges were observed in the included studies. Most notably, a considerable proportion of the studies reported their results only in one measure (accuracy) while not providing other essential measures such as sensitivity and specificity. Another primary concern with these studies is the cost and convenience of the modalities used. A simple brain MRI could cost between $1600 to $8400 in the U.S. [404], while an fMRI scan could cost the patient around $3500 to $5200 [405]. An EEG usually costs lower than MRI, and fMRI scans, with an estimated range of $200 to $700 for a standard EEG, but if extended monitoring is required, the cost could go as high as $3000 [406]. However, ASD patient cooperation in performing an EEG is often poor, requiring sedation, which decreases the convenience of the procedure [407].

Our results also indicate that overfitting is a serious issue in the included studies, as the meta-regression revealed that sample size was significantly correlated with accuracy results across the studies. A recent paper by Ying 2019 [408] offers a variety of solutions to address the overfitting problem. These include the early-stopping strategy, network-reduction strategy, data expansion, and use of regularization terms and dropout technique. Also, linear classifiers are less prone to overfitting than non-linear classifiers due to their lower complexity [116]. Such strategies should be strongly considered in future studies, especially with small sample sizes.

A significant flaw in the literature was that most studies relied on only two class labels (ASD and Non-ASD) for data labeling, assuming that both cases and controls are well-defined entities. This approach ignores that ASD can be broken down into subgroups such as typical autism, low/high function autism, idiopathic autism, and monogenic autism. This approach also ignores that the diagnostic criteria for ASD are composed of behavioral symptoms that may overlap with other mental disorders, such as attention deficit hyperactivity disorder (ADHD).

### Overall completeness and applicability

There were no significant concerns regarding completeness and applicability in this review. Assessments of indirectness using the GRADE tool confirmed this.

### Quality of the evidence

Unfortunately, the currently available evidence is rife with bias, especially in the patient selection domain. Future studies should be more accurate in their methods to gain robust results. Also, we could not include a considerable proportion of the studies in meta-analyses, as these studies only reported one single evaluation metric (mainly accuracy). We tried contacting the authors for additional data but were unsuccessful in most cases. Including those studies could have had a significant effect on our results.

### Potential biases in the review process

There was a considerable risk of publication bias, which downgraded our results’ confidence.

### Implications for research

Our recommendations for future research are summarized in **Highlight box 2**. The most prominent weakness in the literature was the need for better methodological and reporting quality. For example, in most studies, we couldn’t assess the risk of bias for at least one domain simply due to inadequate reporting. We strongly suggest that future researchers consider using standard reporting guidelines such as MINIMAR [409], CLAIM [410], and STARD-AI [411], which are specifically developed for reporting AI studies in healthcare.

We also detected a similar pattern across the literature: most studies implemented an AI algorithm on the data of ASD individuals and reported the results of the classification task, but very few studies tried to explain what features their algorithms use or how justifiable the reasons behind those features used for the classification task. As discussed above, providing a reasonable justification behind the algorithm’s decisions is as important as its results for such a technology to enter the clinical setting. Future studies should consider addressing this issue. Also, future studies should consider deriving more fair decisions from their AI models by (1) pre-processing: transforming the original dataset so that the underlying discrimination towards some groups is removed; (2) in-processing: adding a penalization term in the objective function or imposing a fairness-relevant constraint; and (3) post-processing: further recomputing the results from predictors to improve fairness. More on this subject could be found elsewhere [412].

Future studies must also consider the overfitting problem by addressing data-level and model-level issues. This includes using bigger training data, dropout technique, L1/L2 regularization, and batch normalization [413]. The results of the studies must also be reported in more than just one evaluation metric (sensitivity, specificity, recall, precision, etc.).

Another prominent issue across the literature was the use of datasets that do not reflect the biological diversity of patients optimally, with gender unbalance being the most frequent example of it. Current publicly available datasets contain primarily male patients, which may result in male-biased models. A similar pattern was observed for participants’ functioning, where most data came only from the high-functioning participants. Thus, the development of more balanced datasets seems to be a necessity. An ideal dataset should be Findable, Accessible, Interoperable, and Reusable (FAIR) [414]. More on the requirements for an ideal medical imaging dataset in the era of AI, including the essential metadata required, is discussed elsewhere [415].

We also encourage future researchers to evaluate and compare the biomarkers used to differentiate cases from controls by different models within the same modality, as such data could be of enormous value in finding markers specific to ASD patients and also help illuminate the “black-box” decision-making nature of these models.

Studies should also try to break ASD and non-ASD groups into smaller subgroups like “mild autism” and “severe autism” or “low function autism” and “high function autism.”

Finally, a gap in the knowledge is human-machine collaboration. As AI algorithms can complement, but not replace, physicians in most aspects of medicine, future models should be studied and implemented as an integral part of a complete healthcare system. Indeed, scientists, physicians, patients, regulatory agencies, and health insurance providers need to create a healthcare system that can learn and adapt as it develops [416]. At the same time, AI researchers should recognize the limits of their models to prevent their overuse and misuse, which could otherwise sow distrust and cause patient harm [417].

#### HIGHLIGHT BOX 2 |

Recommendations for future research.

- Use standard reporting guidelines (such as MINIMAR CLAIM, and STARD-AI).
- Use saliency maps, class activation maps (CAM), invertible networks, or other methods of model interpretation to avoid black-box dilemmas in AI modeling.
- Use the early-stopping strategy, network-reduction strategy, data expansion, regularization terms, dropout technique, or other techniques to avoid model overfitting on data.
- Report evaluation performance in more than just one metric (e.g., sensitivity, specificity, positive predictive value, etc.).
- Use datasets that are balanced for the most important potential confounders (e.g., gender, age, IQ, ASD subtype, etc.).
- Develop biologically diverse datasets to address gender unbalance and function unbalance problems of current publicly available datasets (e.g., ABIDE).
- Develop publicly available datasets (like ABIDE for fMRI) for other modalities, especially EEG data as its results are promising.
- Avoid aggregating data from different subtypes of ASD into one general “ASD class”.
- Evaluate the model performance on a separate dataset unseen to the model (i.e., test dataset).
- Avoid applying feature reduction methods to test data as it may induce overfitting.

## Supporting information

Multimedia Appendix 1: Search strategy

Multimedia Appendix 2: Characteristics of included studies

Multimedia Appendix 3: Results of SCS meta-analyses

## Declarations

**Availability of data and materials:** To access the data of the studies, contact their respective authors. Review data are available as appendix files.

**Competing interests:** The authors declare that they have no competing interests.

**Funding:** Not funded

## Authors’ contributions

- Coordination of the review: AV, MM, ANA, AHM
- Designing the study: AV, MM, ANA, SHS, IMO, FA, AHM
- Developing the protocol: AV, MM, ANA, AHM
- Performing the search: AV, ZMG, AM, MG
- Study selection: AV, MM, RA, SHH, MST, ZMG
- Data extraction: AV, RA, SHH, MST, ZMG
- Assessing the risk of bias in included studies: AV, MM, SHH, MST, RA, ZMG, SF
- Analysis of data: AV
- Interpretation of the results: AV, MM, AHM
- Assessing the confidence in cumulative evidence: AV, SHH, MST, RA, ZMG
- Writing the review: AV, MM, ANA, SHS, IMO, FA, AHM
- Correspondence: AHM

### Acknowledgments

**None**

**Registration:** PROSPERO registration ID: CRD42021262575, CRD42021262825, CRD42021262831.

**Amendments:** Initially, we planned to research three modalities (EEG, rs-fMRI, and S-MRI) and publish the results in three papers. However, we decided to extend our research after posting our protocol and before conducting our search to present all the results in one comprehensive article.

## Data Availability

To access the data of the studies, contact their respective authors. Review data are available as appendix files.

## Bibliography

[1] M. J. Maenner et al., ‘Prevalence and Characteristics of Autism Spectrum Disorder Among Children Aged 8 Years — Autism and Developmental Disabilities Monitoring Network, 11 Sites, United States, 2018’, MMWR. Surveillance Summaries, vol. 70, no. 11, pp. 1–16, Dec. 2021, doi: 10.15585/mmwr.ss7011a1.

[2] J. Zeidan et al., ‘Global prevalence of autism: A systematic review update’, Autism Research, vol. 15, no. 5, pp. 778–790, May 2022, doi: 10.1002/aur.2696.

[3] P. M. Dietz, C. E. Rose, D. McArthur, and M. Maenner, ‘National and State Estimates of Adults with Autism Spectrum Disorder’, J Autism Dev Disord, vol. 50, no. 12, pp. 4258–4266, Dec. 2020, doi: 10.1007/s10803-020-04494-4.

[4] J. P. Leigh and J. Du, ‘Brief Report: Forecasting the Economic Burden of Autism in 2015 and 2025 in the United States’, J Autism Dev Disord, vol. 45, no. 12, pp. 4135– 4139, Dec. 2015, doi: 10.1007/s10803-015-2521-7.

[5] C. Solomon, ‘Autism and Employment: Implications for Employers and Adults with ASD’, J Autism Dev Disord, vol. 50, no. 11, pp. 4209–4217, Nov. 2020, doi: 10.1007/s10803-020-04537-w.

[6] B. Oakley, E. Loth, and D. G. Murphy, ‘Autism and mood disorders’, International Review of Psychiatry, vol. 33, no. 3, pp. 280–299, Apr. 2021, doi: 10.1080/09540261.2021.1872506.

[7] F. Thabtah and D. Peebles, ‘Early Autism Screening: A Comprehensive Review’, Int J Environ Res Public Health, vol. 16, no. 18, p. 3502, Sep. 2019, doi: 10.3390/ijerph16183502.

[8] American Psychiatric Association, Diagnostic and Statistical Manual of Mental Disorders, vol. 21. American Psychiatric Association, 2013. doi: 10.1176/appi.books.9780890425596.

[9] S. J. Rogers, ‘Brief report: Early intervention in autism’, J Autism Dev Disord, vol. 26, no. 2, pp. 243–246, Apr. 1996, doi: 10.1007/BF02172020.

[10] L. Vllasaliu et al., ‘Diagnostic instruments for autism spectrum disorder (ASD)’, Cochrane Database of Systematic Reviews, no. 1, Jan. 2016, doi: 10.1002/14651858.CD012036.

[11] G. Baird, H. R. Douglas, a. director, and M. S. Murphy, ‘Recognising and diagnosing autism in children and young people: summary of NICE guidance’, BMJ, vol. 343, no. oct21 1, pp. d6360–d6360, Oct. 2011, doi: 10.1136/bmj.d6360.

[12] M. Xu, V. Calhoun, R. Jiang, W. Yan, and J. Sui, ‘Brain imaging-based machine learning in autism spectrum disorder: methods and applications’, J Neurosci Methods, vol. 361, p. 109271, Sep. 2021, doi: 10.1016/j.jneumeth.2021.109271.

[13] M. Liu, B. Li, and D. Hu, ‘Autism Spectrum Disorder Studies Using fMRI Data and Machine Learning: A Review’, Front Neurosci, vol. 15, p. 1111, Sep. 2021, doi: 10.3389/fnins.2021.697870.

[14] A. Chaddad et al., ‘Can Autism Be Diagnosed with Artificial Intelligence? A Narrative Review’, Diagnostics, vol. 11, no. 11, p. 2032, Nov. 2021, doi: 10.3390/diagnostics11112032.

[15] H. S. Nogay and H. Adeli, ‘Machine learning (ML) for the diagnosis of autism spectrum disorder (ASD) using brain imaging’, Rev Neurosci, vol. 31, no. 8, pp. 825– 841, Nov. 2020, doi: 10.1515/revneuro-2020-0043.

[16] S. J. Moon, J. Hwang, R. Kana, J. Torous, and J. W. Kim, ‘Accuracy of Machine Learning Algorithms for the Diagnosis of Autism Spectrum Disorder: Systematic Review and Meta-Analysis of Brain Magnetic Resonance Imaging Studies’, JMIR Ment Health, vol. 6, no. 12, p. e14108, Dec. 2019, doi: 10.2196/14108.

[17] C. Lefebvre, et al., ‘Searching for and selecting studies’, in Cochrane Handbook for Systematic Reviews of Interventions version 6.3, 6.3., J. Higgins, J. Thomas, J. Chandler, M. Cumpston, T. Li, M. Page, and V. Welch, Eds. Cochrane, 2022. Accessed: Sep. 13, 2022. [Online]. Available: www.training.cochrane.org/handbook

[18] D. Sharpe, ‘Of apples and oranges, file drawers and garbage: Why validity issues in meta-analysis will not go away’, Clin Psychol Rev, vol. 17, no. 8, pp. 881–901, Dec. 1997, doi: 10.1016/S0272-7358(97)00056-1.

[19] The U.S. Food and Drug Administration, ‘FDA permits marketing of artificial intelligence-based device to detect certain diabetes-related eye problems’, Apr. 11, 2018. https://www.fda.gov/news-events/press-announcements/fda-permits-marketing-artificial-intelligence-based-device-detect-certain-diabetes-related-eye (accessed Mar. 17, 2022).

[20] The Medical Futurist, ‘FDA-approved A.I.-based algorithms’, Mar. 17, 2022. https://medicalfuturist.com/fda-approved-ai-based-algorithms/ (accessed Mar. 17, 2022).

[21] S. Benjamens, P. Dhunnoo, and B. Meskó, ‘The state of artificial intelligence-based FDA-approved medical devices and algorithms: an online database’, NPJ Digit Med, vol. 3, no. 1, p. 118, Dec. 2020, doi: 10.1038/s41746-020-00324-0.

[22] G. Hinton, ‘Deep Learning—A Technology With the Potential to Transform Health Care’, JAMA, vol. 320, no. 11, p. 1101, Sep. 2018, doi: 10.1001/jama.2018.11100.

[23] C. J. Kelly, A. Karthikesalingam, M. Suleyman, G. Corrado, and D. King, ‘Key challenges for delivering clinical impact with artificial intelligence’, BMC Med, vol. 17, no. 1, p. 195, Dec. 2019, doi: 10.1186/s12916-019-1426-2.

[24] O. Esteban et al., ‘fMRIPrep: a robust preprocessing pipeline for functional MRI’, Nat Methods, vol. 16, no. 1, pp. 111–116, Jan. 2019, doi: 10.1038/s41592-018-0235-4.

[25] Ryan Holbrook, ‘Creating Features’, Kaggle, 2022. https://www.kaggle.com/code/ryanholbrook/creating-features (accessed Sep. 13, 2022).

[26] J. Brank et al., ‘Feature Selection’, in *Encyclopedia of Machine Learning*, C. Sammut and G. I. Webb, Eds. Boston, MA: Springer US, 2011, pp. 402–406. doi: 10.1007/978-0-387-30164-8_306.

[27] T. Dietterich, ‘Overfitting and undercomputing in machine learning’, ACM Comput Surv, vol. 27, no. 3, pp. 326–327, Sep. 1995, doi: 10.1145/212094.212114.

[28] F. Maleki, N. Muthukrishnan, K. Ovens, C. Reinhold, and R. Forghani, ‘Machine Learning Algorithm Validation’, Neuroimaging Clin N Am, vol. 30, no. 4, pp. 433– 445, Nov. 2020, doi: 10.1016/j.nic.2020.08.004.

[29] E. Tacconelli, ‘Systematic reviews: CRD’s guidance for undertaking reviews in health care’, Lancet Infect Dis, vol. 10, no. 4, p. 226, Apr. 2010, doi: 10.1016/S1473-3099(10)70065-7.

[30] M. D. F. McInnes et al., ‘Preferred Reporting Items for a Systematic Review and Meta-analysis of Diagnostic Test Accuracy Studies’, JAMA, vol. 319, no. 4, p. 388, Jan. 2018, doi: 10.1001/jama.2017.19163.

[31] M. L. Rethlefsen et al., ‘PRISMA-S: an extension to the PRISMA Statement for Reporting Literature Searches in Systematic Reviews’, Syst Rev, vol. 10, no. 1, p. 39, Dec. 2021, doi: 10.1186/s13643-020-01542-z.

[32] N. Sorden, ‘New MeSH browser available’, NLM Tech. Bull., vol. 413, 2016.

[33] L. Slater, ‘PubMed PubReMiner’, Journal of the Canadian Health Libraries Association / Journal de l’Association des bibliothèques de la santé du Canada, vol. 33, no. 2, p. 106, Jul. 2014, doi: 10.5596/c2012-014.

[34] J. McGowan, M. Sampson, D. M. Salzwedel, E. Cogo, V. Foerster, and C. Lefebvre, ‘PRESS Peer Review of Electronic Search Strategies: 2015 Guideline Statement’, J Clin Epidemiol, vol. 75, pp. 40–46, Jul. 2016, doi: 10.1016/j.jclinepi.2016.01.021.

[35] M. Hupe, ‘EndNote X9’, Journal of Electronic Resources in Medical Libraries, vol. 16, no. 3–4, pp. 117–119, Oct. 2019, doi: 10.1080/15424065.2019.1691963.

[36] M. Ouzzani, H. Hammady, Z. Fedorowicz, and A. Elmagarmid, ‘Rayyan—a web and mobile app for systematic reviews’, Syst Rev, vol. 5, no. 1, p. 210, Dec. 2016, doi: 10.1186/s13643-016-0384-4.

[37] P. F. Whiting, ‘QUADAS-2: A Revised Tool for the Quality Assessment of Diagnostic Accuracy Studies’, Ann Intern Med, vol. 155, no. 8, p. 529, Oct. 2011, doi: 10.7326/0003-4819-155-8-201110180-00009.

[38] A. Vabalas, E. Gowen, E. Poliakoff, and A. J. Casson, ‘Machine learning algorithm validation with a limited sample size’, PLoS One, vol. 14, no. 11, p. e0224365, Nov. 2019, doi: 10.1371/journal.pone.0224365.

[39] R Core Team, ‘R: A language and environment for statistical computing’, 2013.

[40] L. Furuya-Kanamori, P. Kostoulas, and S. A. R. Doi, ‘A new method for synthesizing test accuracy data outperformed the bivariate method’, J Clin Epidemiol, vol. 132, pp. 51–58, Apr. 2021, doi: 10.1016/j.jclinepi.2020.12.015.

[41] J. B. Reitsma, A. S. Glas, A. W. S. Rutjes, R. J. P. M. Scholten, P. M. Bossuyt, and A. H. Zwinderman, ‘Bivariate analysis of sensitivity and specificity produces informative summary measures in diagnostic reviews’, J Clin Epidemiol, vol. 58, no. 10, pp. 982–990, Oct. 2005, doi: 10.1016/j.jclinepi.2005.02.022.

[42] S. A. R. Doi, J. J. Barendregt, S. Khan, L. Thalib, and G. M. Williams, ‘Advances in the meta-analysis of heterogeneous clinical trials I: The inverse variance heterogeneity model’, Contemp Clin Trials, vol. 45, pp. 130–138, Nov. 2015, doi: 10.1016/j.cct.2015.05.009.

[43] J. P. T. Higgins and S. G. Thompson, ‘Quantifying heterogeneity in a meta-analysis’, Stat Med, vol. 21, no. 11, pp. 1539–1558, Jun. 2002, doi: 10.1002/sim.1186.

[44] P. Macaskill, Y. Takwoingi, J. Deeks, and C. Gatsonis, ‘Understanding meta-analysis’, in Cochrane Handbook for Systematic Reviews of Diagnostic Test Accuracy, 2nd ed., J. Deeks, P. Bossuyt, M. Leeflang, and Y. Takwoingi, Eds. London: Cochrane, 2022. [Online]. Available: www.training.cochrane.org/handbook

[45] F. Song et al., ‘Dissemination and publication of research findings: an updated review of related biases’, Health Technol Assess (Rockv), vol. 14, no. 8, pp. 1–115, Feb. 2010, doi: 10.3310/hta14080.

[46] S. Duval and R. Tweedie, ‘Trim and Fill: A Simple Funnel-Plot-Based Method of Testing and Adjusting for Publication Bias in Meta-Analysis’, Biometrics, vol. 56, no. 2, pp. 455–463, Jun. 2000, doi: 10.1111/j.0006-341X.2000.00455.x.

[47] P.-C. Bürkner and P. Doebler, ‘Testing for publication bias in diagnostic meta-analysis: a simulation study’, Stat Med, vol. 33, no. 18, pp. 3061–3077, Aug. 2014, doi: 10.1002/sim.6177.

[48] J. J. Deeks, P. Macaskill, and L. Irwig, ‘The performance of tests of publication bias and other sample size effects in systematic reviews of diagnostic test accuracy was assessed’, J Clin Epidemiol, vol. 58, no. 9, pp. 882–893, Sep. 2005, doi: 10.1016/j.jclinepi.2005.01.016.

[49] J. L. Brożek et al., ‘Grading quality of evidence and strength of recommendations in clinical practice guidelines: Part 2 of 3. The GRADE approach to grading quality of evidence about diagnostic tests and strategies’, Allergy, vol. 64, no. 8, pp. 1109– 1116, Aug. 2009, doi: 10.1111/j.1398-9995.2009.02083.x.

[50] X. Bi, J. Chen, Q. Sun, Y. Liu, Y. Wang, and X. Luo, ‘Analysis of Asperger Syndrome Using Genetic-Evolutionary Random Support Vector Machine Cluster’, Front Physiol, vol. 9, p. 1646, Nov. 2018, doi: 10.3389/fphys.2018.01646.

[51] R. W. Emerson et al., ‘Functional neuroimaging of high-risk 6-month-old infants predicts a diagnosis of autism at 24 months of age’, Sci Transl Med, vol. 9, no. 393, p. eaag2882, Jun. 2017, doi: 10.1126/scitranslmed.aag2882.

[52] C. Wang, Z. Xiao, B. Wang, and J. Wu, ‘Identification of Autism Based on SVM-RFE and Stacked Sparse Auto-Encoder’, IEEE Access, vol. 7, pp. 118030–118036, 2019, doi: 10.1109/ACCESS.2019.2936639.

[53] A. M. Mahmoud and H. Karamti, ‘Classifying a type of brain disorder in children: an effective fMRI based deep attempt’, Indonesian Journal of Electrical Engineering and Computer Science, vol. 22, no. 1, p. 260, Apr. 2021, doi: 10.11591/ijeecs.v22.i1.pp260-269.

[54] C. P. Chen et al., ‘Diagnostic classification of intrinsic functional connectivity highlights somatosensory, default mode, and visual regions in autism’, Neuroimage Clin, vol. 8, pp. 238–245, 2015, doi: 10.1016/j.nicl.2015.04.002.

[55] C. Wang, Z. Xiao, and J. Wu, ‘Functional connectivity-based classification of autism and control using SVM-RFECV on rs-fMRI data’, Physica Medica, vol. 65, pp. 99– 105, Sep. 2019, doi: 10.1016/j.ejmp.2019.08.010.

[56] T. Iidaka, ‘Resting state functional magnetic resonance imaging and neural network classified autism and control’, Cortex, vol. 63, pp. 55–67, Feb. 2015, doi: 10.1016/j.cortex.2014.08.011.

[57] Y. Zhang et al., ‘Self-Paced Learning and Privileged Information based Cascaded Multi-column Classification algorithm for ASD diagnosis’, in 2021 43rd Annual International Conference of the IEEE Engineering in Medicine & Biology Society (EMBC), Nov. 2021, pp. 3281–3284. doi: 10.1109/EMBC46164.2021.9630150.

[58] V. Subbaraju, S. Sundaram, and S. Narasimhan, ‘Identification of lateralized compensatory neural activities within the social brain due to autism spectrum disorder in adolescent males’, European Journal of Neuroscience, vol. 47, no. 6, pp. 631–642, Mar. 2018, doi: 10.1111/ejn.13634.

[59] F. Huang et al., ‘Self-weighted adaptive structure learning for ASD diagnosis via multi-template multi-center representation’, Med Image Anal, vol. 63, p. 101662, Jul. 2020, doi: 10.1016/j.media.2020.101662.

[60] X. Guo, K. C. Dominick, A. A. Minai, H. Li, C. A. Erickson, and L. J. Lu, ‘Diagnosing Autism Spectrum Disorder from Brain Resting-State Functional Connectivity Patterns Using a Deep Neural Network with a Novel Feature Selection Method’, Front Neurosci, vol. 11, p. 460, Aug. 2017, doi: 10.3389/fnins.2017.00460.

[61] N. Yahata et al., ‘A small number of abnormal brain connections predicts adult autism spectrum disorder’, Nat Commun, vol. 7, no. 1, p. 11254, Sep. 2016, doi: 10.1038/ncomms11254.

[62] X. Li, Y. Gu, N. Dvornek, L. H. Staib, P. Ventola, and J. S. Duncan, ‘Multi-site fMRI analysis using privacy-preserving federated learning and domain adaptation: ABIDE results’, Med Image Anal, vol. 65, p. 101765, Oct. 2020, doi: 10.1016/j.media.2020.101765.

[63] H. Sewani and R. Kashef, ‘An Autoencoder-Based Deep Learning Classifier for Efficient Diagnosis of Autism’, Children, vol. 7, no. 10, p. 182, Oct. 2020, doi: 10.3390/children7100182.

[64] N. A. Khan, S. A. Waheeb, A. Riaz, and X. Shang, ‘A Three-Stage Teacher, Student Neural Networks and Sequential Feed Forward Selection-Based Feature Selection Approach for the Classification of Autism Spectrum Disorder’, Brain Sci, vol. 10, no. 10, p. 754, Oct. 2020, doi: 10.3390/brainsci10100754.

[65] F. Huang, A. Elazab, L. OuYang, J. Tan, T. Wang, and B. Lei, ‘Sparse Low-rank Constrained Adaptive Structure Learning using Multi-template for Autism Spectrum Disorder Diagnosis’, in 2019 IEEE 16th International Symposium on Biomedical Imaging (ISBI 2019), Apr. 2019, pp. 1555–1558. doi: 10.1109/ISBI.2019.8759487.

[66] B. Li, A. Sharma, J. Meng, S. Purushwalkam, and E. Gowen, ‘Applying machine learning to identify autistic adults using imitation: An exploratory study’, PLoS One, vol. 12, no. 8, p. e0182652, Aug. 2017, doi: 10.1371/journal.pone.0182652.

[67] B. S. Mahanand, S. Vigneshwaran, S. Suresh, and N. Sundararajan, ‘An enhanced effect-size thresholding method for the diagnosis of Autism Spectrum Disorder using resting state functional MRI’, in 2016 Second International Conference on Cognitive Computing and Information Processing (CCIP), Aug. 2016, pp. 1–6. doi: 10.1109/CCIP.2016.7802874.

[68] D. K and V. R. Murthy Oruganti, ‘A Machine Learning Approach for Diagnosing Neurological Disorders using Longitudinal Resting-State fMRI’, in 2021 *11th International Conference on Cloud Computing*, Data Science & Engineering (Confluence), Jan. 2021, pp. 494–499. doi: 10.1109/Confluence51648.2021.9377173.

[69] F. Z. Benabdallah, A. D. El Maliani, and M. El Hassouni, ‘A CNN Based 3D Connectivity Matrices Features for Autism Detection: Application on ABIDE I’, in Ubiquitous Networking, 2021, pp. 293–302. doi: 10.1007/978-3-030-86356-2_24.

[70] Z. Dai, H. Zhang, F. Lin, S. Feng, Y. Wei, and J. Zhou, ‘The Classification System and Biomarkers for Autism Spectrum Disorder: A Machine Learning Approach’, in Bioinformatics Research and Applications, 2021, pp. 289–299. doi: 10.1007/978-3-030-91415-8_25.

[71] H. Huang, X. Liu, Y. Jin, S. Lee, C. Wee, and D. Shen, ‘Enhancing the representation of functional connectivity networks by fusing multi-view information for autism spectrum disorder diagnosis’, Hum Brain Mapp, vol. 40, no. 3, pp. 833–854, Feb. 2019, doi: 10.1002/hbm.24415.

[72] L. Boppana, N. Shabnam, and T. Srivatsava, ‘Deep Learning Approach for an early stage detection of Neurodevelopmental Disorders’, in 2021 IEEE 9th Region 10 Humanitarian Technology Conference (R10-HTC), Sep. 2021, pp. 1–6. doi: 10.1109/R10-HTC53172.2021.9641691.

[73] H. Chen et al., ‘Multivariate classification of autism spectrum disorder using frequency-specific resting-state functional connectivity—A multi-center study’, Prog Neuropsychopharmacol Biol Psychiatry, vol. 64, pp. 1–9, Jan. 2016, doi: 10.1016/j.pnpbp.2015.06.014.

[74] W. Yin, S. Mostafa, and F. Wu, ‘Diagnosis of Autism Spectrum Disorder Based on Functional Brain Networks with Deep Learning’, Journal of Computational Biology, vol. 28, no. 2, pp. 146–165, Feb. 2021, doi: 10.1089/cmb.2020.0252.

[75] M. Plitt, K. A. Barnes, and A. Martin, ‘Functional connectivity classification of autism identifies highly predictive brain features but falls short of biomarker standards’, Neuroimage Clin, vol. 7, pp. 359–366, 2015, doi: 10.1016/j.nicl.2014.12.013.

[76] J. S. Anderson et al., ‘Functional connectivity magnetic resonance imaging classification of autism’, Brain, vol. 134, no. 12, pp. 3742–3754, Dec. 2011, doi: 10.1093/brain/awr263.

[77] V. Subbaraju, M. B. Suresh, S. Sundaram, and S. Narasimhan, ‘Identifying differences in brain activities and an accurate detection of autism spectrum disorder using resting state functional-magnetic resonance imaging : A spatial filtering approach’, Med Image Anal, vol. 35, pp. 375–389, Jan. 2017, doi: 10.1016/j.media.2016.08.003.

[78] M. Jung et al., ‘Surface-based shared and distinct resting functional connectivity in attention-deficit hyperactivity disorder and autism spectrum disorder’, The British Journal of Psychiatry, vol. 214, no. 06, pp. 339–344, Jun. 2019, doi: 10.1192/bjp.2018.248.

[79] A. Mahmoud, H. Karamti, and F. Alrowais, ‘An Effective Sparse Autoencoders based Deep Learning Framework for fMRI Scans Classification’, in Proceedings of the 22nd International Conference on Enterprise Information Systems, 2020, pp. 540–547. doi: 10.5220/0009397605400547.

[80] S. Shrivastava, U. Mishra, N. Singh, A. Chandra, and S. Verma, ‘Control or Autism - Classification using Convolutional Neural Networks on Functional MRI’, in 2020 *11th International Conference on Computing*, Communication and Networking Technologies (ICCCNT), Jul. 2020, pp. 1–6. doi: 10.1109/ICCCNT49239.2020.9225506.

[81] X. Yang, P. T., and N. Zhang, ‘A Deep Neural Network Study of the ABIDE Repository on Autism Spectrum Classification’, International Journal of Advanced Computer Science and Applications, vol. 11, no. 4, pp. 1–6, 2020, doi: 10.14569/IJACSA.2020.0110401.

[82] B. Yamagata et al., ‘Machine learning approach to identify a resting-state functional connectivity pattern serving as an endophenotype of autism spectrum disorder’, Brain Imaging Behav, vol. 13, no. 6, pp. 1689–1698, Dec. 2019, doi: 10.1007/s11682-018-9973-2.

[83] Y. Wang, J. Wang, F.-X. Wu, R. Hayrat, and J. Liu, ‘AIMAFE: Autism spectrum disorder identification with multi-atlas deep feature representation and ensemble learning’, J Neurosci Methods, vol. 343, p. 108840, Sep. 2020, doi: 10.1016/j.jneumeth.2020.108840.

[84] M. Tang, P. Kumar, H. Chen, and A. Shrivastava, ‘Deep Multimodal Learning for the Diagnosis of Autism Spectrum Disorder’, J Imaging, vol. 6, no. 6, p. 47, Jun. 2020, doi: 10.3390/jimaging6060047.

[85] J. F. Agastinose Ronicko, J. Thomas, P. Thangavel, V. Koneru, G. Langs, and J. Dauwels, ‘Diagnostic classification of autism using resting-state fMRI data improves with full correlation functional brain connectivity compared to partial correlation’, J Neurosci Methods, vol. 345, p. 108884, Nov. 2020, doi: 10.1016/j.jneumeth.2020.108884.

[86] X. Yang, M. S. Islam, and A. M. A. Khaled, ‘Functional connectivity magnetic resonance imaging classification of autism spectrum disorder using the multisite ABIDE dataset’, in 2019 IEEE EMBS International Conference on Biomedical & Health Informatics (BHI), May 2019, pp. 1–4. doi: 10.1109/BHI.2019.8834653.

[87] Z. Jiao, H. Li, and Y. Fan, ‘Improving Diagnosis of Autism Spectrum Disorder and Disentangling its Heterogeneous Functional Connectivity Patterns Using Capsule Networks’, in 2020 IEEE 17th International Symposium on Biomedical Imaging (ISBI), Apr. 2020, pp. 1331–1334. doi: 10.1109/ISBI45749.2020.9098524.

[88] G. Spera et al., ‘Evaluation of Altered Functional Connections in Male Children With Autism Spectrum Disorders on Multiple-Site Data Optimized With Machine Learning’, Front Psychiatry, vol. 10, p. 620, Sep. 2019, doi: 10.3389/fpsyt.2019.00620.

[89] J. Zhuang, N. C. Dvornek, X. Li, P. Ventola, and J. S. Duncan, ‘Invertible Network for Classification and Biomarker Selection for ASD’, in *International Conference on Medical Image Computing and Computer-Assisted Intervention*, Springer, 2019, pp. 700–708. doi: 10.1007/978-3-030-32248-9_78.

[90] S. Gupta, Y. H. Chan, and J. C. Rajapakse, ‘Obtaining leaner deep neural networks for decoding brain functional connectome in a single shot’, Neurocomputing, vol. 453, pp. 326–336, Sep. 2021, doi: 10.1016/j.neucom.2020.04.152.

[91] J. Zhang, F. Feng, T. Han, X. Gong, and F. Duan, ‘Detection of Autism Spectrum Disorder using fMRI Functional Connectivity with Feature Selection and Deep Learning’, Cognit Comput, Jan. 2022, doi: 10.1007/s12559-021-09981-z.

[92] F. Almuqhim and F. Saeed, ‘ASD-SAENet: A Sparse Autoencoder, and Deep-Neural Network Model for Detecting Autism Spectrum Disorder (ASD) Using fMRI Data’, Front Comput Neurosci, vol. 15, p. 654315, Apr. 2021, doi: 10.3389/fncom.2021.654315.

[93] A. Jahedi, C. A. Nasamran, B. Faires, J. Fan, and R.-A. Müller, ‘Distributed Intrinsic Functional Connectivity Patterns Predict Diagnostic Status in Large Autism Cohort’, Brain Connect, vol. 7, no. 8, pp. 515–525, Oct. 2017, doi: 10.1089/brain.2017.0496.

[94] P. Lanka, D. Rangaprakash, M. N. Dretsch, J. S. Katz, T. S. Denney, and G. Deshpande, ‘Supervised machine learning for diagnostic classification from large-scale neuroimaging datasets’, Brain Imaging Behav, vol. 14, no. 6, pp. 2378–2416, Dec. 2020, doi: 10.1007/s11682-019-00191-8.

[95] Y. Zhao, F. Ge, S. Zhang, and T. Liu, ‘3D Deep Convolutional Neural Network Revealed the Value of Brain Network Overlap in Differentiating Autism Spectrum Disorder from Healthy Controls’, in Medical Image Computing and Computer Assisted Intervention – MICCAI 2018, 2018, pp. 172–180. doi: 10.1007/978-3-030-00931-1_20.

[96] H. Li, N. A. Parikh, and L. He, ‘A Novel Transfer Learning Approach to Enhance Deep Neural Network Classification of Brain Functional Connectomes’, Front Neurosci, vol. 12, p. 491, Jul. 2018, doi: 10.3389/fnins.2018.00491.

[97] T. Eslami, V. Mirjalili, A. Fong, A. R. Laird, and F. Saeed, ‘ASD-DiagNet: A Hybrid Learning Approach for Detection of Autism Spectrum Disorder Using fMRI Data’, Front Neuroinform, vol. 13, p. 70, Nov. 2019, doi: 10.3389/fninf.2019.00070.

[98] Z. Sherkatghanad et al., ‘Automated Detection of Autism Spectrum Disorder Using a Convolutional Neural Network’, Front Neurosci, vol. 13, p. 1325, Jan. 2020, doi: 10.3389/fnins.2019.01325.

[99] N. Chaitra, P. A. Vijaya, and G. Deshpande, ‘Diagnostic prediction of autism spectrum disorder using complex network measures in a machine learning framework’, Biomed Signal Process Control, vol. 62, p. 102099, Sep. 2020, doi: 10.1016/j.bspc.2020.102099.

[100] A. S. Heinsfeld, A. R. Franco, R. C. Craddock, A. Buchweitz, and F. Meneguzzi, ‘Identification of autism spectrum disorder using deep learning and the ABIDE dataset’, Neuroimage Clin, vol. 17, pp. 16–23, 2018, doi: 10.1016/j.nicl.2017.08.017.

[101] R. Bhaumik, A. Pradhan, S. Das, and D. K. Bhaumik, ‘Predicting Autism Spectrum Disorder Using Domain-Adaptive Cross-Site Evaluation’, Neuroinformatics, vol. 16, no. 2, pp. 197–205, Apr. 2018, doi: 10.1007/s12021-018-9366-0.

[102] J. Hu, L. Cao, T. Li, B. Liao, S. Dong, and P. Li, ‘Interpretable Learning Approaches in Resting-State Functional Connectivity Analysis: The Case of Autism Spectrum Disorder’, Comput Math Methods Med, vol. 2020, pp. 1–12, May 2020, doi: 10.1155/2020/1394830.

[103] W. Cancino, G. Africano, and S. Pertuz, ‘A Benchmark of Preprocessing Strategies for Autism Classification from Resting-State Functional Magnetic Resonance Imaging’, in 2021 *XXIII Symposium on Image*, Signal Processing and Artificial Vision (STSIVA), Sep. 2021, pp. 1–5. doi: 10.1109/STSIVA53688.2021.9592011.

[104] W. Jung, D.-W. Heo, E. Jeon, J. Lee, and H.-I. Suk, ‘Inter-regional High-Level Relation Learning from Functional Connectivity via Self-supervision’, in Medical Image Computing and Computer Assisted Intervention – MICCAI 2021, 2021, pp. 284–293. doi: 10.1007/978-3-030-87196-3_27.

[105] M. Wang, D. Zhang, J. Huang, D. Shen, and M. Liu, ‘Low-Rank Representation for Multi-center Autism Spectrum Disorder Identification’, in Med Image Comput Comput Assist Interv, vol. 11070, 2018, pp. 647–654. doi: 10.1007/978-3-030-00928-1_73.

[106] Y. Liu, L. Xu, J. Li, J. Yu, and X. Yu, ‘Attentional Connectivity-based Prediction of Autism Using Heterogeneous rs-fMRI Data from CC200 Atlas’, Exp Neurobiol, vol. 29, no. 1, pp. 27–37, Feb. 2020, doi: 10.5607/en.2020.29.1.27.

[107] T.-E. Kam, H.-I. Suk, and S.-W. Lee, ‘Multiple functional networks modeling for autism spectrum disorder diagnosis’, Hum Brain Mapp, vol. 38, no. 11, pp. 5804– 5821, Nov. 2017, doi: 10.1002/hbm.23769.

[108] J. M. Kernbach et al., ‘Shared endo-phenotypes of default mode dysfunction in attention deficit/hyperactivity disorder and autism spectrum disorder’, Transl Psychiatry, vol. 8, no. 1, p. 133, Dec. 2018, doi: 10.1038/s41398-018-0179-6.

[109] A. Abraham et al., ‘Deriving reproducible biomarkers from multi-site resting-state data: An Autism-based example’, Neuroimage, vol. 147, pp. 736–745, Feb. 2017, doi: 10.1016/j.neuroimage.2016.10.045.

[110] M. Leming, J. M. Górriz, and J. Suckling, ‘Ensemble Deep Learning on Large, Mixed-Site fMRI Datasets in Autism and Other Tasks’, Int J Neural Syst, vol. 30, no. 07, p. 2050012, Jul. 2020, doi: 10.1142/S0129065720500124.

[111] N. Okamoto and H. Akama, ‘Extended Invariant Information Clustering Is Effective for Leave-One-Site-Out Cross-Validation in Resting State Functional Connectivity Modeling’, Front Neuroinform, vol. 15, p. 709179, Dec. 2021, doi: 10.3389/fninf.2021.709179.

[112] X. Xing, J. Ji, and Y. Yao, ‘Convolutional Neural Network with Element-wise Filters to Extract Hierarchical Topological Features for Brain Networks’, in 2018 IEEE International Conference on Bioinformatics and Biomedicine (BIBM), Dec. 2018, pp. 780–783. doi: 10.1109/BIBM.2018.8621472.

[113] A. J. Fredo, A. Jahedi, M. Reiter, and R.-A. Müller, ‘Diagnostic classification of autism using resting-state fMRI data and conditional random forest’, in 40th Annual International Conference of the IEEE Engineering in Medicine and Biology Society, Jul. 2018, vol. 12, no. 2.76, pp. 6–41.

[114] I. Mhiri and I. Rekik, ‘Joint functional brain network atlas estimation and feature selection for neurological disorder diagnosis with application to autism’, Med Image Anal, vol. 60, p. 101596, Feb. 2020, doi: 10.1016/j.media.2019.101596.

[115] G. Xu and Y. Liang, ‘A Novel Two-stage Prediction Model to Classify Functional Connectivity for Brain Disease Diagnosis’, in 2021 IEEE 4th International Conference on Computer and Communication Engineering Technology (CCET), Aug. 2021, pp. 63–68. doi: 10.1109/CCET52649.2021.9544424.

[116] P. Kassraian-Fard, C. Matthis, J. H. Balsters, M. H. Maathuis, and N. Wenderoth, ‘Promises, Pitfalls, and Basic Guidelines for Applying Machine Learning Classifiers to Psychiatric Imaging Data, with Autism as an Example’, Front Psychiatry, vol. 7, p. 177, Dec. 2016, doi: 10.3389/fpsyt.2016.00177.

[117] L. Dodero, H. Q. Minh, M. S. Biagio, V. Murino, and D. Sona, ‘Kernel-based classification for brain connectivity graphs on the Riemannian manifold of positive definite matrices’, in 2015 IEEE 12th International Symposium on Biomedical Imaging (ISBI), Apr. 2015, pp. 42–45. doi: 10.1109/ISBI.2015.7163812.

[118] J. A. Nielsen et al., ‘Multisite functional connectivity MRI classification of autism: ABIDE results’, Front Hum Neurosci, vol. 7, 2013, doi: 10.3389/fnhum.2013.00599.

[119] K. Niu et al., ‘Multichannel Deep Attention Neural Networks for the Classification of Autism Spectrum Disorder Using Neuroimaging and Personal Characteristic Data’, Complexity, vol. 2020, pp. 1–9, Jan. 2020, doi: 10.1155/2020/1357853.

[120] M. A. Reiter, A. Jahedi, A. R. J. Fredo, I. Fishman, B. Bailey, and R.-A. Müller, ‘Performance of machine learning classification models of autism using resting-state fMRI is contingent on sample heterogeneity’, Neural Comput Appl, vol. 33, no. 8, pp. 3299–3310, Apr. 2021, doi: 10.1007/s00521-020-05193-y.

[121] X. Bi et al., ‘The Genetic-Evolutionary Random Support Vector Machine Cluster Analysis in Autism Spectrum Disorder’, IEEE Access, vol. 7, pp. 30527–30535, 2019, doi: 10.1109/ACCESS.2019.2902889.

[122] X. Bi, Y. Wang, Q. Shu, Q. Sun, and Q. Xu, ‘Classification of Autism Spectrum Disorder Using Random Support Vector Machine Cluster’, Front Genet, vol. 9, p. 18, Feb. 2018, doi: 10.3389/fgene.2018.00018.

[123] X. Bi, Y. Liu, Q. Jiang, Q. Shu, Q. Sun, and J. Dai, ‘The Diagnosis of Autism Spectrum Disorder Based on the Random Neural Network Cluster’, Front Hum Neurosci, vol. 12, p. 257, Jun. 2018, doi: 10.3389/fnhum.2018.00257.

[124] A. Kazeminejad and R. C. Sotero, ‘Topological Properties of Resting-State fMRI Functional Networks Improve Machine Learning-Based Autism Classification’, Front Neurosci, vol. 12, p. 1018, Jan. 2019, doi: 10.3389/fnins.2018.01018.

[125] P. Barttfeld et al., ‘State-dependent changes of connectivity patterns and functional brain network topology in autism spectrum disorder’, Neuropsychologia, vol. 50, no. 14, pp. 3653–3662, Dec. 2012, doi: 10.1016/j.neuropsychologia.2012.09.047.

[126] J.-W. Sun, R. Fan, Q. Wang, Q.-Q. Wang, X.-Z. Jia, and H.-B. Ma, ‘Identify abnormal functional connectivity of resting state networks in Autism spectrum disorder and apply to machine learning-based classification’, Brain Res, vol. 1757, p. 147299, Apr. 2021, doi: 10.1016/j.brainres.2021.147299.

[127] C. Zu et al., ‘Identifying disease-related subnetwork connectome biomarkers by sparse hypergraph learning’, Brain Imaging Behav, vol. 13, no. 4, pp. 879–892, Aug. 2019, doi: 10.1007/s11682-018-9899-8.

[128] C. Shi, J. Zhang, and X. Wu, ‘An fMRI Feature Selection Method Based on a Minimum Spanning Tree for Identifying Patients with Autism’, Symmetry (Basel), vol. 12, no. 12, p. 1995, Dec. 2020, doi: 10.3390/sym12121995.

[129] S. Gupta, J. C. Rajapakse, and R. E. Welsch, ‘Ambivert degree identifies crucial brain functional hubs and improves detection of Alzheimer’s Disease and Autism Spectrum Disorder’, Neuroimage Clin, vol. 25, p. 102186, 2020, doi: 10.1016/j.nicl.2020.102186.

[130] R. Anirudh and J. J. Thiagarajan, ‘Bootstrapping Graph Convolutional Neural Networks for Autism Spectrum Disorder Classification’, in *ICASSP 2019 -2019 IEEE International Conference on Acoustics*, Speech and Signal Processing (ICASSP), May 2019, pp. 3197–3201. doi: 10.1109/ICASSP.2019.8683547.

[131] L. Shao, C. Fu, Y. You, and D. Fu, ‘Classification of ASD based on fMRI data with deep learning’, Cogn Neurodyn, vol. 15, no. 6, pp. 961–974, Dec. 2021, doi: 10.1007/s11571-021-09683-0.

[132] S. Mostafa, L. Tang, and F.-X. Wu, ‘Diagnosis of Autism Spectrum Disorder Based on Eigenvalues of Brain Networks’, IEEE Access, vol. 7, pp. 128474–128486, 2019, doi: 10.1109/ACCESS.2019.2940198.

[133] Y. Chen, A. Liu, X. Fu, J. Wen, and X. Chen, ‘An Invertible Dynamic Graph Convolutional Network for Multi-Center ASD Classification’, Front Neurosci, vol. 15, p. 828512, Feb. 2022, doi: 10.3389/fnins.2021.828512.

[134] Y. Wang, J. Liu, Y. Xiang, J. Wang, Q. Chen, and J. Chong, ‘MAGE: Automatic diagnosis of autism spectrum disorders using multi-atlas graph convolutional networks and ensemble learning’, Neurocomputing, vol. 469, pp. 346–353, Jan. 2022, doi: 10.1016/j.neucom.2020.06.152.

[135] Y. Song, T. M. Epalle, and H. Lu, ‘Characterizing and Predicting Autism Spectrum Disorder by Performing Resting-State Functional Network Community Pattern Analysis’, Front Hum Neurosci, vol. 13, p. 203, Jun. 2019, doi: 10.3389/fnhum.2019.00203.

[136] L. Zhang, J.-R. Wang, and Y. Ma, ‘Graph Convolutional Networks via Low-Rank Subspace for Multi-Site rs-fMRI ASD Diagnosis’, in 2021 *14th International Congress on Image and Signal Processing*, BioMedical Engineering and Informatics (CISP-BMEI), Oct. 2021, pp. 1–6. doi: 10.1109/CISP-BMEI53629.2021.9624374.

[137] Z. Rakhimberdina, X. Liu, and T. Murata, ‘Population Graph-Based Multi-Model Ensemble Method for Diagnosing Autism Spectrum Disorder’, Sensors, vol. 20, no. 21, p. 6001, Oct. 2020, doi: 10.3390/s20216001.

[138] M. Cao et al., ‘Using DeepGCN to identify the autism spectrum disorder from multi-site resting-state data’, Biomed Signal Process Control, vol. 70, p. 103015, Sep. 2021, doi: 10.1016/j.bspc.2021.103015.

[139] C. Yang, P. Wang, J. Tan, Q. Liu, and X. Li, ‘Autism spectrum disorder diagnosis using graph attention network based on spatial-constrained sparse functional brain networks’, Comput Biol Med, vol. 139, p. 104963, Dec. 2021, doi: 10.1016/j.compbiomed.2021.104963.

[140] S. Parisot et al., ‘Disease prediction using graph convolutional networks: Application to Autism Spectrum Disorder and Alzheimer’s disease’, Med Image Anal, vol. 48, pp. 117–130, Aug. 2018, doi: 10.1016/j.media.2018.06.001.

[141] S. Ataei, N. Attar, S. Aliakbary, and F. Bakouie, ‘Graph theoretical approach for screening autism on brain complex networks’, SN Appl Sci, vol. 1, no. 9, p. 1122, Sep. 2019, doi: 10.1007/s42452-019-1079-y.

[142] S. Parisot et al., ‘Spectral Graph Convolutions for Population-Based Disease Prediction’, in *International conference on medical image computing and computer-assisted intervention*, Springer, 2017, pp. 177–185. doi: 10.1007/978-3-319-66179-7_21.

[143] C. J. Brown, J. Kawahara, and G. Hamarneh, ‘Connectome priors in deep neural networks to predict autism’, in 2018 IEEE 15th International Symposium on Biomedical Imaging (ISBI 2018), Apr. 2018, pp. 110–113. doi: 10.1109/ISBI.2018.8363534.

[144] D. Yao et al., ‘Triplet Graph Convolutional Network for Multi-scale Analysis of Functional Connectivity Using Functional MRI’, in Graph Learning in Medical Imaging, 2019, pp. 70–78. doi: 10.1007/978-3-030-35817-4_9.

[145] H. Jiang, P. Cao, M. Xu, J. Yang, and O. Zaiane, ‘Hi-GCN: A hierarchical graph convolution network for graph embedding learning of brain network and brain disorders prediction’, Comput Biol Med, vol. 127, p. 104096, Dec. 2020, doi: 10.1016/j.compbiomed.2020.104096.

[146] E. Tolan and Z. Isik, ‘Graph Theory Based Classification of Brain Connectivity Network for Autism Spectrum Disorder’, in Bioinformatics and Biomedical Engineering, 2018, pp. 520–530. doi: 10.1007/978-3-319-78723-7_45.

[147] M. Sadeghi, R. Khosrowabadi, F. Bakouie, H. Mahdavi, C. Eslahchi, and H. Pouretemad, ‘Screening of autism based on task-free fMRI using graph theoretical approach’, Psychiatry Res Neuroimaging, vol. 263, pp. 48–56, May 2017, doi: 10.1016/j.pscychresns.2017.02.004.

[148] L. Dodero, F. Sambataro, V. Murino, and D. Sona, ‘Kernel-Based Analysis of Functional Brain Connectivity on Grassmann Manifold’, in Medical Image Computing and Computer-Assisted Intervention – MICCAI 2015, 2015, pp. 604–611. doi: 10.1007/978-3-319-24574-4_72.

[149] S. I. Ktena et al., ‘Metric learning with spectral graph convolutions on brain connectivity networks’, Neuroimage, vol. 169, pp. 431–442, Apr. 2018, doi: 10.1016/j.neuroimage.2017.12.052.

[150] A. Kazeminejad and R. C. Sotero, ‘The Importance of Anti-correlations in Graph Theory Based Classification of Autism Spectrum Disorder’, Front Neurosci, vol. 14, p. 676, Aug. 2020, doi: 10.3389/fnins.2020.00676.

[151] K. Byeon, J. Kwon, J. Hong, and H. Park, ‘Artificial Neural Network Inspired by Neuroimaging Connectivity: Application in Autism Spectrum Disorder’, in 2020 IEEE International Conference on Big Data and Smart Computing (BigComp), Feb. 2020, pp. 575–578. doi: 10.1109/BigComp48618.2020.00013.

[152] S. M. Jain, ‘Detection of Autism using Magnetic Resonance Imaging data and Graph Convolutional Neural Networks’, Rochester Institute of Technology, Rochester, 2018.

[153] E. Jun and H.-I. Suk, *Connectomics in NeuroImaging*, vol. 10511. Cham: Springer International Publishing, 2017. doi: 10.1007/978-3-319-67159-8.

[154] T. Price, C.-Y. Wee, W. Gao, and D. Shen, ‘Multiple-Network Classification of Childhood Autism Using Functional Connectivity Dynamics’, in *International Conference on Medical Image Computing and Computer-Assisted Intervention*, Springer, 2014, pp. 177–184. doi: 10.1007/978-3-319-10443-0_23.

[155] Z. Xiao, C. Wang, N. Jia, and J. Wu, ‘SAE-based classification of school-aged children with autism spectrum disorders using functional magnetic resonance imaging’, Multimed Tools Appl, vol. 77, no. 17, pp. 22809–22820, Sep. 2018, doi: 10.1007/s11042-018-5625-1.

[156] M. Naghashzadeh, M. Yazdi, and A. Zolghadrasli, ‘Classification of autism spectrum disorders individuals and controls using phase and envelope features from resting-state fMRI data’, Comput Methods Biomech Biomed Eng Imaging Vis, vol. 10, no. 1, pp. 55–66, Jan. 2022, doi: 10.1080/21681163.2021.1972343.

[157] M. I. Al-Hiyali, N. Yahya, I. Faye, Z. Khan, and K. Alsaih, ‘Classification of BOLD FMRI Signals using Wavelet Transform and Transfer Learning for Detection of Autism Spectrum Disorder’, in 2020 IEEE-EMBS Conference on Biomedical Engineering and Sciences (IECBES), Mar. 2021, pp. 94–98. doi: 10.1109/IECBES48179.2021.9398803.

[158] F. Zhao, Z. Chen, I. Rekik, S.-W. Lee, and D. Shen, ‘Diagnosis of Autism Spectrum Disorder Using Central-Moment Features From Low- and High-Order Dynamic Resting-State Functional Connectivity Networks’, Front Neurosci, vol. 14, p. 258, Apr. 2020, doi: 10.3389/fnins.2020.00258.

[159] X. Ma, X.-H. Wang, and L. Li, ‘Identifying individuals with autism spectrum disorder based on the principal components of whole-brain phase synchrony’, Neurosci Lett, vol. 742, p. 135519, Jan. 2021, doi: 10.1016/j.neulet.2020.135519.

[160] J. Liu, Y. Sheng, W. Lan, R. Guo, Y. Wang, and J. Wang, ‘Improved ASD classification using dynamic functional connectivity and multi-task feature selection’, Pattern Recognit Lett, vol. 138, pp. 82–87, Oct. 2020, doi: 10.1016/j.patrec.2020.07.005.

[161] A. S. Karampasi, A. D. Savva, V. Ch. Korfiatis, I. Kakkos, and G. K. Matsopoulos, ‘Informative Biomarkers for Autism Spectrum Disorder Diagnosis in Functional Magnetic Resonance Imaging Data on the Default Mode Network’, Applied Sciences, vol. 11, no. 13, p. 6216, Jul. 2021, doi: 10.3390/app11136216.

[162] E. Jun, E. Kang, J. Choi, and H.-I. Suk, ‘Modeling regional dynamics in low-frequency fluctuation and its application to Autism spectrum disorder diagnosis’, Neuroimage, vol. 184, pp. 669–686, Jan. 2019, doi: 10.1016/j.neuroimage.2018.09.043.

[163] G. Fan et al., ‘Abnormal Brain Regions in Two-Group Cross-Location Dynamics Model of Autism’, IEEE Access, vol. 8, pp. 94526–94534, 2020, doi: 10.1109/ACCESS.2020.2995209.

[164] M. A. Bayram, İ. Özer, and F. Temurtas, ‘Deep Learning Methods for Autism Spectrum Disorder Diagnosis Based on fMRI Images’, Sakarya University Journal of Computer and Information Sciences, vol. 4, no. 1, pp. 142–155, Apr. 2021, doi: 10.35377/saucis.04.01.879735.

[165] Y. Liu, L. Xu, J. Yu, J. Li, and X. Yu, ‘Identification of autism spectrum disorder using multi-regional resting-state data through an attention learning approach’, Biomed Signal Process Control, vol. 69, p. 102833, Aug. 2021, doi: 10.1016/j.bspc.2021.102833.

[166] P. S. Dammu and R. S. Bapi, ‘Employing Temporal Properties of Brain Activity for Classifying Autism Using Machine Learning’, in Pattern Recognition and Machine Intelligence, 2019, pp. 193–200. doi: 10.1007/978-3-030-34872-4_22.

[167] C.-Y. Wee, P.-T. Yap, and D. Shen, ‘Diagnosis of Autism Spectrum Disorders Using Temporally Distinct Resting-State Functional Connectivity Networks’, CNS Neurosci Ther, vol. 22, no. 3, pp. 212–219, Mar. 2016, doi: 10.1111/cns.12499.

[168] M. A. Aghdam, A. Sharifi, and M. M. Pedram, ‘Diagnosis of Autism Spectrum Disorders in Young Children Based on Resting-State Functional Magnetic Resonance Imaging Data Using Convolutional Neural Networks’, J Digit Imaging, vol. 32, no. 6, pp. 899–918, Dec. 2019, doi: 10.1007/s10278-019-00196-1.

[169] A. D. Savva, A. S. Karampasi, and G. K. Matsopoulos, ‘Deriving resting-state fMRI biomarkers for classification of autism spectrum disorder’, in *Autism 360°*, Elsevier, 2020, pp. 101–123. doi: 10.1016/B978-0-12-818466-0.00006-X.

[170] N. C. Dvornek, P. Ventola, K. A. Pelphrey, and J. S. Duncan, Machine Learning in Medical Imaging, vol. 10541. Cham: Springer International Publishing, 2017. doi: 10.1007/978-3-319-67389-9.

[171] R. Tejwani, A. Liska, H. You, J. Reinen, and P. Das, ‘Autism Classification Using Brain Functional Connectivity Dynamics and Machine Learning’, arXiv preprint arXiv:1712.08041, Dec. 2017.

[172] M. D. Schirmer et al., ‘Neuropsychiatric disease classification using functional connectomics - results of the connectomics in neuroimaging transfer learning challenge’, Med Image Anal, vol. 70, p. 101972, May 2021, doi: 10.1016/j.media.2021.101972.

[173] L. Rabany et al., ‘Dynamic functional connectivity in schizophrenia and autism spectrum disorder: Convergence, divergence and classification’, Neuroimage Clin, vol. 24, p. 101966, 2019, doi: 10.1016/j.nicl.2019.101966.

[174] B. Huang, ‘Diagnosis of autism spectrum disorder by causal influence strength learned from resting-state fMRI data’, in *Neural Engineering Techniques for Autism Spectrum Disorder*, Elsevier, 2021, pp. 237–267. doi: 10.1016/B978-0-12-822822-7.00012-0.

[175] H. Haghighat, M. Mirzarezaee, B. Nadjar Araabi, and A. Khadem, ‘An age-dependent Connectivity-based computer aided diagnosis system for Autism Spectrum Disorder using Resting-state fMRI’, Biomed Signal Process Control, vol. 71, p. 103108, Jan. 2022, doi: 10.1016/j.bspc.2021.103108.

[176] M. S. Ahammed, S. Niu, M. R. Ahmed, J. Dong, X. Gao, and Y. Chen, ‘DarkASDNet: Classification of ASD on Functional MRI Using Deep Neural Network’, Front Neuroinform, vol. 15, p. 635657, Jun. 2021, doi: 10.3389/fninf.2021.635657.

[177] M. S. Ahammed, S. Niu, M. R. Ahmed, J. Dong, X. Gao, and Y. Chen, ‘Bag-of-Features Model for ASD fMRI Classification using SVM’, in 2021 Asia-Pacific Conference on Communications Technology and Computer Science (ACCTCS), Jan. 2021, pp. 52–57. doi: 10.1109/ACCTCS52002.2021.00019.

[178] Z. Xiao, J. Wu, C. Wang, N. Jia, and X. Yang, ‘Computer-aided diagnosis of school-aged children with ASD using full frequency bands and enhanced SAE: A multi-institution study’, Exp Ther Med, vol. 17, no. 5, pp. 4055–4063, Mar. 2019, doi: 10.3892/etm.2019.7448.

[179] O. Dekhil et al., ‘Identifying Personalized Autism Related Impairments Using Resting Functional MRI and ADOS Reports’, in Medical Image Computing and Computer Assisted Intervention – MICCAI 2018, 2018, pp. 240–248. doi: 10.1007/978-3-030-00931-1_28.

[180] S. Vigneshwaran, B. S. Mahanand, S. Suresh, and N. Sundararajan, ‘Using regional homogeneity from functional MRI for diagnosis of ASD among males’, in 2015 International Joint Conference on Neural Networks (IJCNN), Jul. 2015, pp. 1–8. doi: 10.1109/IJCNN.2015.7280562.

[181] S. Rane, E. Jolly, A. Park, H. Jang, and C. Craddock, ‘Developing predictive imaging biomarkers using whole-brain classifiers: Application to the ABIDE I dataset’, Res Ideas Outcomes, vol. 3, p. e12733, Mar. 2017, doi: 10.3897/rio.3.e12733.

[182] F. Zhao, H. Zhang, I. Rekik, Z. An, and D. Shen, ‘Diagnosis of Autism Spectrum Disorders Using Multi-Level High-Order Functional Networks Derived From Resting-State Functional MRI’, Front Hum Neurosci, vol. 12, p. 184, May 2018, doi: 10.3389/fnhum.2018.00184.

[183] Y. Liang, B. Liu, and H. Zhang, ‘A Convolutional Neural Network Combined With Prototype Learning Framework for Brain Functional Network Classification of Autism Spectrum Disorder’, IEEE Transactions on Neural Systems and Rehabilitation Engineering, vol. 29, pp. 2193–2202, 2021, doi: 10.1109/TNSRE.2021.3120024.

[184] J. Wang, Q. Wang, H. Zhang, J. Chen, S. Wang, and D. Shen, ‘Sparse Multiview Task-Centralized Ensemble Learning for ASD Diagnosis Based on Age- and Sex-Related Functional Connectivity Patterns’, IEEE Trans Cybern, vol. 49, no. 8, pp. 3141–3154, Aug. 2019, doi: 10.1109/TCYB.2018.2839693.

[185] J. Wang, Q. Wang, S. Wang, and D. Shen, ‘Sparse Multi-view Task-Centralized Learning for ASD Diagnosis’, in Mach Learn Med Imaging, vol. 10541, 2017, pp. 159–167. doi: 10.1007/978-3-319-67389-9_19.

[186] S. Sartipi, M. G. Shayesteh, and H. Kalbkhani, ‘Diagnosing of Autism Spectrum Disorder based on GARCH Variance Series for rs-fMRI data’, in 2018 9th International Symposium on Telecommunications (IST), Dec. 2018, pp. 86–90. doi: 10.1109/ISTEL.2018.8661147.

[187] A. Bernas, A. P. Aldenkamp, and S. Zinger, ‘Wavelet coherence-based classifier: A resting-state functional MRI study on neurodynamics in adolescents with high-functioning autism’, Comput Methods Programs Biomed, vol. 154, pp. 143–151, Feb. 2018, doi: 10.1016/j.cmpb.2017.11.017.

[188] M. A. Syed, Z. Yang, X. P. Hu, and G. Deshpande, ‘Investigating Brain Connectomic Alterations in Autism Using the Reproducibility of Independent Components Derived from Resting State Functional MRI Data’, Front Neurosci, vol. 11, p. 459, Sep. 2017, doi: 10.3389/fnins.2017.00459.

[189] L. Q. Uddin et al., ‘Salience Network–Based Classification and Prediction of Symptom Severity in Children With Autism’, JAMA Psychiatry, vol. 70, no. 8, p. 869, Aug. 2013, doi: 10.1001/jamapsychiatry.2013.104.

[190] M. Yang et al., ‘Large-Scale Brain Functional Network Integration for Discrimination of Autism Using a 3-D Deep Learning Model’, Front Hum Neurosci, vol. 15, p. 687288, Jun. 2021, doi: 10.3389/fnhum.2021.687288.

[191] S. Ghiassian, R. Greiner, P. Jin, and M. R. G. Brown, ‘Using Functional or Structural Magnetic Resonance Images and Personal Characteristic Data to Identify ADHD and Autism’, PLoS One, vol. 11, no. 12, p. e0166934, Dec. 2016, doi: 10.1371/journal.pone.0166934.

[192] M. Khosla, K. Jamison, A. Kuceyeski, and M. R. Sabuncu, ‘3D Convolutional Neural Networks for Classification of Functional Connectomes’, in *Deep Learning in Medical Image Analysis and Multimodal Learning for Clinical Decision Support*, Springer, 2018, pp. 137–145. doi: 10.1007/978-3-030-00889-5_16.

[193] A. Karampasi et al., ‘A Machine Learning fMRI Approach in the Diagnosis of Autism’, in 2020 IEEE International Conference on Big Data (Big Data), Dec. 2020, pp. 3628– 3631. doi: 10.1109/BigData50022.2020.9378453.

[194] Y. You, H. Liu, S. Zhang, and L. Shao, ‘Classification of Autism Based on fMRI Data with Feature-Fused Convolutional Neural Network’, in *Cyberspace Data and Intelligence, and Cyber-Living*, Syndrome, and Health, 2020, pp. 77–88. doi: 10.1007/978-981-33-4336-8_7.

[195] A. Rathore, S. Palande, J. S. Anderson, B. A. Zielinski, P. T. Fletcher, and B. Wang, ‘Autism Classification Using Topological Features and Deep Learning: A Cautionary Tale’, in Med Image Comput Comput Assist Interv, vol. 11766, 2019, pp. 736–744. doi: 10.1007/978-3-030-32248-9_82.

[196] A. Sadiq, M. I. Al-Hiyali, N. Yahya, T. B. Tang, and D. M. Khan, ‘Non-Oscillatory Connectivity Approach for Classification of Autism Spectrum Disorder Subtypes Using Resting-State fMRI’, IEEE Access, vol. 10, pp. 14049–14061, 2022, doi: 10.1109/ACCESS.2022.3146719.

[197] R. M. Thomas, S. Gallo, L. Cerliani, P. Zhutovsky, A. El-Gazzar, and G. van Wingen, ‘Classifying Autism Spectrum Disorder Using the Temporal Statistics of Resting-State Functional MRI Data With 3D Convolutional Neural Networks’, Front Psychiatry, vol. 11, p. 440, May 2020, doi: 10.3389/fpsyt.2020.00440.

[198] L. Herath, D. Meedeniya, M. A. J. C. Marasingha, and V. Weerasinghe, ‘Autism spectrum disorder diagnosis support model using Inception V3’, in 2021 International Research Conference on Smart Computing and Systems Engineering (SCSE), Sep. 2021, vol. 4, pp. 1–7. doi: 10.1109/SCSE53661.2021.9568314.

[199] M. R. Ahmed, Y. Zhang, Y. Liu, and H. Liao, ‘Single Volume Image Generator and Deep Learning-Based ASD Classification’, IEEE J Biomed Health Inform, vol. 24, no. 11, pp. 3044–3054, Nov. 2020, doi: 10.1109/JBHI.2020.2998603.

[200] N. Dominic, T. W. Cenggoro, A. Budiarto, and B. Pardamean, ‘Transfer learning using inception-ResNet-v2 model to the augmented neuroimages data for autism spectrum disorder classification’, Communications in Mathematical Biology and Neuroscience, vol. 2021, p. Article ID 39, 2021, doi: 10.28919/cmbn/5565.

[201] N. Adluru et al., ‘Classification in DTI using shapes of white matter tracts’, in 2009 Annual International Conference of the IEEE Engineering in Medicine and Biology Society, Sep. 2009, pp. 2719–2722. doi: 10.1109/IEMBS.2009.5333386.

[202] Z. Zhang and W. Zheng, ‘The Discriminative Power of White Matter Microstructures for Autism Diagnosis’, IFAC-PapersOnLine, vol. 53, no. 5, pp. 446–451, 2020, doi: 10.1016/j.ifacol.2021.04.121.

[203] F. Zhang et al., ‘Fiber clustering based white matter connectivity analysis for prediction of Autism Spectrum Disorder using diffusion tensor imaging’, in 2016 IEEE 13th International Symposium on Biomedical Imaging (ISBI), Apr. 2016, pp. 564–567. doi: 10.1109/ISBI.2016.7493331.

[204] M. Ingalhalikar, D. Parker, L. Bloy, T. P. L. Roberts, and R. Verma, ‘Diffusion based abnormality markers of pathology: Toward learned diagnostic prediction of ASD’, Neuroimage, vol. 57, no. 3, pp. 918–927, Aug. 2011, doi: 10.1016/j.neuroimage.2011.05.023.

[205] F. Zhang et al., ‘Whole brain white matter connectivity analysis using machine learning: An application to autism’, Neuroimage, vol. 172, pp. 826–837, May 2018, doi: 10.1016/j.neuroimage.2017.10.029.

[206] M. Mostapha, M. F. Casanova, G. Gimel’farb, and A. El-Baz, ‘Towards Non-invasive Image-Based Early Diagnosis of Autism’, in Medical Image Computing and Computer-Assisted Intervention – MICCAI 2015, 2015, pp. 160–168. doi: 10.1007/978-3-319-24571-3_20.

[207] Y. ElNakieb et al., ‘The Role of Diffusion Tensor MR Imaging (DTI) of the Brain in Diagnosing Autism Spectrum Disorder: Promising Results’, Sensors, vol. 21, no. 24, p. 8171, Dec. 2021, doi: 10.3390/s21248171.

[208] N. Lange et al., ‘Atypical diffusion tensor hemispheric asymmetry in autism’, Autism Research, vol. 3, no. 6, pp. 350–358, Dec. 2010, doi: 10.1002/aur.162.

[209] Y. A. Elnakieb et al., ‘Computer Aided Autism Diagnosis Using Diffusion Tensor Imaging’, IEEE Access, vol. 8, pp. 191298–191308, 2020, doi: 10.1109/ACCESS.2020.3032066.

[210] J. Kang, Y. Jin, G. Liang, and X. Li, ‘Accurate assessment of low-function autistic children based on EEG feature fusion’, Journal of Clinical Neuroscience, vol. 90, pp. 351–358, Aug. 2021, doi: 10.1016/j.jocn.2021.06.022.

[211] E. Grossi, C. Olivieri, and M. Buscema, ‘Diagnosis of autism through EEG processed by advanced computational algorithms: A pilot study’, Comput Methods Programs Biomed, vol. 142, pp. 73–79, Apr. 2017, doi: 10.1016/j.cmpb.2017.02.002.

[212] M. Ahmadlou, H. Adeli, and A. Adeli, ‘Fuzzy Synchronization Likelihood-wavelet methodology for diagnosis of autism spectrum disorder’, J Neurosci Methods, vol. 211, no. 2, pp. 203–209, Nov. 2012, doi: 10.1016/j.jneumeth.2012.08.020.

[213] H. Behnam, A. Sheikhani, M. R. Mohammadi, M. Noroozian, and P. Golabi, ‘Abnormalities in Connectivity of Quantitative Electroencephalogram Background Activity in Autism Disorders especially in Left Hemisphere and Right Temporal’, in Tenth International Conference on Computer Modeling and Simulation (uksim 2008), Apr. 2008, pp. 82–87. doi: 10.1109/UKSIM.2008.68.

[214] W. Jamal, S. Das, I.-A. Oprescu, K. Maharatna, F. Apicella, and F. Sicca, ‘Classification of autism spectrum disorder using supervised learning of brain connectivity measures extracted from synchrostates’, J Neural Eng, vol. 11, no. 4, p. 046019, Aug. 2014, doi: 10.1088/1741-2560/11/4/046019.

[215] T. Wadhera and D. Kakkar, ‘Social cognition and functional brain network in autism spectrum disorder: Insights from EEG graph-theoretic measures’, Biomed Signal Process Control, vol. 67, p. 102556, May 2021, doi: 10.1016/j.bspc.2021.102556.

[216] F. H. Duffy and H. Als, ‘A stable pattern of EEG spectral coherence distinguishes children with autism from neuro-typical controls - a large case control study’, BMC Med, vol. 10, no. 1, p. 64, Dec. 2012, doi: 10.1186/1741-7015-10-64.

[217] G. S. Bajestani, M. Behrooz, A. G. Khani, M. Nouri-Baygi, and A. Mollaei, ‘Diagnosis of autism spectrum disorder based on complex network features’, Comput Methods Programs Biomed, vol. 177, pp. 277–283, Aug. 2019, doi: 10.1016/j.cmpb.2019.06.006.

[218] N. Satheesh Kumar, J. Mohanalin, and J. Mahil, ‘Recognition of autism in children via electroencephalogram behaviour using particle swarm optimization based ANFIS classifier’, Multimed Tools Appl, vol. 79, no. 13–14, pp. 8747–8766, Apr. 2020, doi: 10.1007/s11042-018-6290-0.

[219] D. Abdolzadegan, M. H. Moattar, and M. Ghoshuni, ‘A robust method for early diagnosis of autism spectrum disorder from EEG signals based on feature selection and DBSCAN method’, Biocybern Biomed Eng, vol. 40, no. 1, pp. 482–493, Jan. 2020, doi: 10.1016/j.bbe.2020.01.008.

[220] W. J. Bosl, T. Loddenkemper, and C. A. Nelson, ‘Nonlinear EEG biomarker profiles for autism and absence epilepsy’, Neuropsychiatr Electrophysiol, vol. 3, no. 1, p. 1, Dec. 2017, doi: 10.1186/s40810-017-0023-x.

[221] T. Heunis et al., ‘Recurrence quantification analysis of resting state EEG signals in autism spectrum disorder – a systematic methodological exploration of technical and demographic confounders in the search for biomarkers’, BMC Med, vol. 16, no. 1, p. 101, Dec. 2018, doi: 10.1186/s12916-018-1086-7.

[222] T. A. Manoharan and M. Radhakrishnan, ‘Region-Wise Brain Response Classification of ASD Children Using EEG and BiLSTM RNN’, Clin EEG Neurosci, p. 155005942110549, Nov. 2021, doi: 10.1177/15500594211054990.

[223] W. J. Bosl, H. Tager-Flusberg, and C. A. Nelson, ‘EEG Analytics for Early Detection of Autism Spectrum Disorder: A data-driven approach’, Sci Rep, vol. 8, no. 1, p. 6828, Dec. 2018, doi: 10.1038/s41598-018-24318-x.

[224] S. Ibrahim, R. Djemal, and A. Alsuwailem, ‘Electroencephalography (EEG) signal processing for epilepsy and autism spectrum disorder diagnosis’, Biocybern Biomed Eng, vol. 38, no. 1, pp. 16–26, 2018, doi: 10.1016/j.bbe.2017.08.006.

[225] J. Castelhano, P. Tavares, S. Mouga, G. Oliveira, and M. Castelo-Branco, ‘Stimulus dependent neural oscillatory patterns show reliable statistical identification of autism spectrum disorder in a face perceptual decision task’, Clinical Neurophysiology, vol. 129, no. 5, pp. 981–989, May 2018, doi: 10.1016/j.clinph.2018.01.072.

[226] A. K. Subudhi, M. Mohanty, S. K. Sahoo, S. K. Mohanty, and B. Mohanty, ‘Automated Delimitation and Classification of Autistic Disorder Using EEG Signal’, IETE J Res, pp. 1–9, Nov. 2020, doi: 10.1080/03772063.2020.1844076.

[227] J. Eldridge, A. E. Lane, M. Belkin, and S. Dennis, ‘Robust features for the automatic identification of autism spectrum disorder in children’, J Neurodev Disord, vol. 6, no. 1, p. 12, Dec. 2014, doi: 10.1186/1866-1955-6-12.

[228] A. Gui et al., ‘Attentive brain states in infants with and without later autism’, Transl Psychiatry, vol. 11, no. 1, p. 196, Dec. 2021, doi: 10.1038/s41398-021-01315-9.

[229] E. Askari, S. K. Setarehdan, A. Sheikhani, M. R. Mohammadi, and M. Teshnehlab, ‘Computational model for detection of abnormal brain connections in children with autism’, J Integr Neurosci, vol. 17, no. 3, pp. 237–248, Aug. 2018, doi: 10.31083/JIN-180075.

[230] R. Djemal, K. AlSharabi, S. Ibrahim, and A. Alsuwailem, ‘EEG-Based Computer Aided Diagnosis of Autism Spectrum Disorder Using Wavelet, Entropy, and ANN’, Biomed Res Int, vol. 2017, pp. 1–9, 2017, doi: 10.1155/2017/9816591.

[231] T.-H. Pham et al., ‘Autism Spectrum Disorder Diagnostic System Using HOS Bispectrum with EEG Signals’, Int J Environ Res Public Health, vol. 17, no. 3, p. 971, Feb. 2020, doi: 10.3390/ijerph17030971.

[232] F. Salehi, M. Jaloli, R. Coben, and A. M. Nasrabadi, ‘Estimating brain effective connectivity from EEG signals of patients with autism disorder and healthy individuals by reducing volume conduction effect’, Cogn Neurodyn, vol. 16, no. 3, pp. 519–529, Jun. 2022, doi: 10.1007/s11571-021-09730-w.

[233] F. A. Alturki, K. AlSharabi, A. M. Abdurraqeeb, and M. Aljalal, ‘EEG Signal Analysis for Diagnosing Neurological Disorders Using Discrete Wavelet Transform and Intelligent Techniques’, Sensors, vol. 20, no. 9, p. 2505, Apr. 2020, doi: 10.3390/s20092505.

[234] X. Li, E. Cai, L. Qin, and J. Kang, ‘[Abnormal electroencephalogram features extraction of autistic children based on wavelet transform combined with empirical modal decomposition].’, Sheng Wu Yi Xue Gong Cheng Xue Za Zhi, vol. 35, no. 4, pp. 524–529, 2018, doi: 10.7507/1001-5515.201705067.

[235] E. Askari, S. K. Setarehdan, A. Sheikhani, M. R. Mohammadi, and M. Teshnehlab, ‘Modeling the connections of brain regions in children with autism using cellular neural networks and electroencephalography analysis’, Artif Intell Med, vol. 89, pp. 40–50, Jul. 2018, doi: 10.1016/j.artmed.2018.05.003.

[236] Y. Jayawardana, M. Jaime, and S. Jayarathna, ‘Analysis of Temporal Relationships between ASD and Brain Activity through EEG and Machine Learning’, in 2019 IEEE 20th International Conference on Information Reuse and Integration for Data Science (IRI), Jul. 2019, pp. 151–158. doi: 10.1109/IRI.2019.00035.

[237] M. Baygin et al., ‘Automated ASD detection using hybrid deep lightweight features extracted from EEG signals’, Comput Biol Med, vol. 134, p. 104548, Jul. 2021, doi: 10.1016/j.compbiomed.2021.104548.

[238] A. Sheikhani, H. Behnam, M. Noroozian, M. R. Mohammadi, and M. Mohammadi, ‘Abnormalities of quantitative electroencephalography in children with Asperger disorder in various conditions’, Res Autism Spectr Disord, vol. 3, no. 2, pp. 538–546, Apr. 2009, doi: 10.1016/j.rasd.2008.11.002.

[239] M. N. A. Tawhid, S. Siuly, and H. Wang, ‘Diagnosis of autism spectrum disorder from EEG using a time–frequency spectrogram image-based approach’, Electron Lett, vol. 56, no. 25, pp. 1372–1375, Dec. 2020, doi: 10.1049/el.2020.2646.

[240] E. Abdulhay et al., ‘Computer-aided autism diagnosis via second-order difference plot area applied to EEG empirical mode decomposition’, Neural Comput Appl, vol. 32, no. 15, pp. 10947–10956, Aug. 2020, doi: 10.1007/s00521-018-3738-0.

[241] D. Haputhanthri et al., ‘An EEG based Channel Optimized Classification Approach for Autism Spectrum Disorder’, in 2019 Moratuwa Engineering Research Conference (MERCon), Jul. 2019, pp. 123–128. doi: 10.1109/MERCon.2019.8818814.

[242] M. I. Kamel et al., ‘EEG based Autism Diagnosis Using Regularized Fisher Linear Discriminant Analysis’, *International Journal of Image*, Graphics and Signal Processing, vol. 4, no. 3, pp. 35–41, Apr. 2012, doi: 10.5815/ijigsp.2012.03.06.

[243] W. Khazaal Shams and A. W. Abdul Rahman, ‘Characterizing autistic disorder based on Principle Component Analysis’, in 2011 IEEE Symposium on Industrial Electronics and Applications, Sep. 2011, pp. 653–657. doi: 10.1109/ISIEA.2011.6108797.

[244] E. A. Alsaggaf and M. I. Kamel, ‘Using EEGs to diagnose autism disorder by classification algorithm’, Life Sci J, vol. 11, no. 6, pp. 305–308, 2014, doi: 10.7537/marslsj110614.40.

[245] N. Razali and A. Wahab, ‘2D affective space model (ASM) for detecting autistic children’, in 2011 IEEE 15th International Symposium on Consumer Electronics (ISCE), Jun. 2011, pp. 536–541. doi: 10.1109/ISCE.2011.5973888.

[246] A. Salekin and N. Russo, ‘Understanding autism’, in Proceedings of the Workshop on Medical Cyber Physical Systems and Internet of Medical Things, May 2021, pp. 12–16. doi: 10.1145/3446913.3460317.

[247] M. Radhakrishnan, K. Ramamurthy, K. K. Choudhury, D. Won, and T. A. Manoharan, ‘Performance Analysis of Deep Learning Models for Detection of Autism Spectrum Disorder from EEG Signals’, Traitement du Signal, vol. 38, no. 3, pp. 853– 863, Jun. 2021, doi: 10.18280/ts.380332.

[248] A. Sheikhani, H. Behnam, M. R. Mohammadi, M. Noroozian, and P. Golabi, ‘Analysis of quantitative Electroencephalogram background activity in Autism disease patients with Lempel-Ziv complexity and Short Time Fourier Transform measure’, in 2007 4th IEEE/EMBS International Summer School and Symposium on Medical Devices and Biosensors, Aug. 2007, pp. 111–114. doi: 10.1109/ISSMDBS.2007.4338305.

[249] Q. Mohi-Ud-Din and A. K. Jayanthy, ‘EEG feature extraction using wavelet transform for classifying autism spectrum disorder’, Mater Today Proc, Mar. 2021, doi: 10.1016/j.matpr.2021.01.803.

[250] J. Zhao, J. Song, X. Li, and J. Kang, ‘A study on EEG feature extraction and classification in autistic children based on singular spectrum analysis method’, Brain Behav, vol. 10, no. 12, p. e01721, Dec. 2020, doi: 10.1002/brb3.1721.

[251] M. Ranjani and P. Supraja, ‘Classifying the Autism and Epilepsy Disorder Based on EEG Signal Using Deep Convolutional Neural Network (DCNN)’, in 2021 International Conference on Advance Computing and Innovative Technologies in Engineering (ICACITE), Mar. 2021, pp. 880–886. doi: 10.1109/ICACITE51222.2021.9404634.

[252] A. R. Aslam, N. Hafeez, H. Heidari, and M. A. Bin Altaf, ‘An 8.62 μ W Processor for Autism Spectrum Disorder Classification using Shallow Neural Network’, in 2021 IEEE 3rd International Conference on Artificial Intelligence Circuits and Systems (AICAS), Jun. 2021, pp. 1–4. doi: 10.1109/AICAS51828.2021.9458412.

[253] N. Ghoreishi, A. Goshvarpour, S. Zare-Molekabad, N. Khorshidi, and S. Baratzade, ‘Classification of autistic children using polar-based lagged state-space indices of EEG signals’, Signal Image Video Process, vol. 15, no. 8, pp. 1805–1812, Nov. 2021, doi: 10.1007/s11760-021-01928-z.

[254] G. Bouallegue, R. Djemal, S. A. Alshebeili, and H. Aldhalaan, ‘A Dynamic Filtering DF-RNN Deep-Learning-Based Approach for EEG-Based Neurological Disorders Diagnosis’, IEEE Access, vol. 8, pp. 206992–207007, 2020, doi: 10.1109/ACCESS.2020.3037995.

[255] N. A. Ali, ‘Autism spectrum disorder classification on electroencephalogram signal using deep learning algorithm’, IAES International Journal of Artificial Intelligence (IJ-AI), vol. 9, no. 1, p. 91, Mar. 2020, doi: 10.11591/ijai.v9.i1.pp91-99.

[256] M. Ahmadlou, H. Adeli, and A. Adeli, ‘Fractality and a Wavelet-Chaos-Neural Network Methodology for EEG-Based Diagnosis of Autistic Spectrum Disorder’, Journal of Clinical Neurophysiology, vol. 27, no. 5, pp. 328–333, Oct. 2010, doi: 10.1097/WNP.0b013e3181f40dc8.

[257] A. Sheikhani, H. Behnam, M. R. Mohammadi, M. Noroozian, and M. Mohammadi, ‘Detection of Abnormalities for Diagnosing of Children with Autism Disorders Using of Quantitative Electroencephalography Analysis’, J Med Syst, vol. 36, no. 2, pp. 957–963, Apr. 2012, doi: 10.1007/s10916-010-9560-6.

[258] D. Alie, M. H. Mahoor, W. I. Mattson, D. R. Anderson, and D. S. Messinger, ‘Analysis of eye gaze pattern of infants at risk of autism spectrum disorder using Markov models’, in 2011 IEEE Workshop on Applications of Computer Vision (WACV), Jan. 2011, pp. 282–287. doi: 10.1109/WACV.2011.5711515.

[259] S. Ozdemir, I. Akin-Bulbul, I. Kok, and S. Ozdemir, ‘Development of a visual attention based decision support system for autism spectrum disorder screening’, International Journal of Psychophysiology, vol. 173, pp. 69–81, Mar. 2022, doi: 10.1016/j.ijpsycho.2022.01.004.

[260] R. Carette, F. Cilia, G. Dequen, J. Bosche, J.-L. Guerin, and L. Vandromme, ‘Automatic Autism Spectrum Disorder Detection Thanks to Eye-Tracking and Neural Network-Based Approach’, in Internet of Things (IoT) Technologies for HealthCare, 2018, pp. 75–81. doi: 10.1007/978-3-319-76213-5_11.

[261] R. Carette, M. Elbattah, F. Cilia, G. Dequen, J.-L. Guérin, and J. Bosche, ‘Learning to Predict Autism Spectrum Disorder based on the Visual Patterns of Eye-tracking Scanpaths’, in Proceedings of the 12th International Joint Conference on Biomedical Engineering Systems and Technologies, 2019, pp. 103–112. doi: 10.5220/0007402601030112.

[262] J. S. Oliveira et al., ‘Computer-aided autism diagnosis based on visual attention models using eye tracking’, Sci Rep, vol. 11, no. 1, p. 10131, Dec. 2021, doi: 10.1038/s41598-021-89023-8.

[263] E. Khalaji, ‘A machine learning approach for detecting high-functioning autism using web-based eye-tracking data’, Middle East Technical University, Ankara, 2021.

[264] V. Yaneva, L. A. Ha, S. Eraslan, Y. Yesilada, and R. Mitkov, ‘Detecting Autism Based on Eye-Tracking Data from Web Searching Tasks’, in Proceedings of the 15th International Web for All Conference, Apr. 2018, pp. 1–10. doi: 10.1145/3192714.3192819.

[265] V. Yaneva, L. A. Ha, S. Eraslan, Y. Yesilada, and R. Mitkov, ‘Detecting High-Functioning Autism in Adults Using Eye Tracking and Machine Learning’, IEEE Transactions on Neural Systems and Rehabilitation Engineering, vol. 28, no. 6, pp. 1254–1261, Jun. 2020, doi: 10.1109/TNSRE.2020.2991675.

[266] S. Eraslan, Y. Yesilada, V. Yaneva, and S. Harper, ‘Autism detection based on eye movement sequences on the web’, in Proceedings of the 17th International Web for All Conference, Apr. 2020, pp. 1–10. doi: 10.1145/3371300.3383340.

[267] M. Jiang and Q. Zhao, ‘Learning Visual Attention to Identify People with Autism Spectrum Disorder’, in 2017 IEEE International Conference on Computer Vision (ICCV), Oct. 2017, pp. 3287–3296. doi: 10.1109/ICCV.2017.354.

[268] G. Arru, P. Mazumdar, and F. Battisti, ‘Exploiting Visual Behaviour for Autism Spectrum Disorder Identification’, in 2019 IEEE International Conference on Multimedia & Expo Workshops (ICMEW), Jul. 2019, pp. 637–640. doi: 10.1109/ICMEW.2019.00123.

[269] S. Rahman, S. Rahman, O. Shahid, Md. T. Abdullah, and J. A. Sourov, ‘Classifying Eye-Tracking Data Using Saliency Maps’, in 2020 25th International Conference on Pattern Recognition (ICPR), Jan. 2021, pp. 9288–9295. doi: 10.1109/ICPR48806.2021.9412308.

[270] S. Chen and Q. Zhao, ‘Attention-Based Autism Spectrum Disorder Screening With Privileged Modality’, in 2019 IEEE/CVF International Conference on Computer Vision (ICCV), Oct. 2019, pp. 1181–1190. doi: 10.1109/ICCV.2019.00127.

[271] G. Wan et al., ‘Applying Eye Tracking to Identify Autism Spectrum Disorder in Children’, J Autism Dev Disord, vol. 49, no. 1, pp. 209–215, Jan. 2019, doi: 10.1007/s10803-018-3690-y.

[272] S. Liaqat et al., ‘Predicting ASD diagnosis in children with synthetic and image-based eye gaze data’, Signal Process Image Commun, vol. 94, p. 116198, May 2021, doi: 10.1016/j.image.2021.116198.

[273] M. Alcañiz et al., ‘Eye gaze as a biomarker in the recognition of autism spectrum disorder using virtual reality and machine learning: A proof of concept for diagnosis’, Autism Research, vol. 15, no. 1, pp. 131–145, Jan. 2022, doi: 10.1002/aur.2636.

[274] Y. Lin, Y. Gu, Y. Xu, S. Hou, R. Ding, and S. Ni, ‘Autistic spectrum traits detection and early screening: A machine learning based eye movement study’, Journal of Child and Adolescent Psychiatric Nursing, vol. 35, no. 1, pp. 83–92, Feb. 2022, doi: 10.1111/jcap.12346.

[275] W. Liu, M. Li, and L. Yi, ‘Identifying children with autism spectrum disorder based on their face processing abnormality: A machine learning framework’, Autism Research, vol. 9, no. 8, pp. 888–898, Aug. 2016, doi: 10.1002/aur.1615.

[276] W. Liu, X. Yu, B. Raj, L. Yi, X. Zou, and M. Li, ‘Efficient autism spectrum disorder prediction with eye movement: A machine learning framework’, in 2015 International Conference on Affective Computing and Intelligent Interaction (ACII), Sep. 2015, pp. 649–655. doi: 10.1109/ACII.2015.7344638.

[277] J. Kang, X. Han, J.-F. Hu, H. Feng, and X. Li, ‘The study of the differences between low-functioning autistic children and typically developing children in the processing of the own-race and other-race faces by the machine learning approach’, Journal of Clinical Neuroscience, vol. 81, pp. 54–60, Nov. 2020, doi: 10.1016/j.jocn.2020.09.039.

[278] Z. Zhao, H. Tang, X. Zhang, X. Qu, X. Hu, and J. Lu, ‘Classification of Children With Autism and Typical Development Using Eye-Tracking Data From Face-to-Face Conversations: Machine Learning Model Development and Performance Evaluation’, J Med Internet Res, vol. 23, no. 8, p. e29328, Aug. 2021, doi: 10.2196/29328.

[279] A. Lu and M. Perkowski, ‘Deep Learning Approach for Screening Autism Spectrum Disorder in Children with Facial Images and Analysis of Ethnoracial Factors in Model Development and Application’, Brain Sci, vol. 11, no. 11, p. 1446, Oct. 2021, doi: 10.3390/brainsci11111446.

[280] M. Beary, A. Hadsell, R. Messersmith, and M.-P. Hosseini, ‘Diagnosis of Autism in Children using Facial Analysis and Deep Learning’, arXiv preprint arXiv:2008.02890, Aug. 2020.

[281] P. V. K. Sandeep and N. S. Kumar, ‘Autism Detection in Children with Facial Cues using Dense Net Deep Learning Architecture’, Design Engineering, vol. 2021, no. 9, pp. 7506–7520, 2021.

[282] S. Ram Arumugam, S. Ganesh Karuppasamy, S. Gowr, O. Manoj, and K. Kalaivani, ‘A Deep Convolutional Neural Network based Detection System for Autism Spectrum Disorder in Facial images’, in 2021 *Fifth International Conference on I-SMAC (IoT in Social, Mobile*, Analytics and Cloud) (I-SMAC), Nov. 2021, pp. 1255– 1259. doi: 10.1109/I-SMAC52330.2021.9641046.

[283] S. R. Arumugam, R. Balakrishna, R. Khilar, O. Manoj, and C. S. Shylaja, ‘Prediction of Autism Spectrum Disorder in Children using Face Recognition’, in 2021 2nd International Conference on Smart Electronics and Communication (ICOSEC), Oct. 2021, pp. 1246–1250. doi: 10.1109/ICOSEC51865.2021.9591679.

[284] T. Akter et al., ‘Improved Transfer-Learning-Based Facial Recognition Framework to Detect Autistic Children at an Early Stage’, Brain Sci, vol. 11, no. 6, p. 734, May 2021, doi: 10.3390/brainsci11060734.

[285] K. K. Mujeeb Rahman and M. M. Subashini, ‘Identification of Autism in Children Using Static Facial Features and Deep Neural Networks’, Brain Sci, vol. 12, no. 1, p. 94, Jan. 2022, doi: 10.3390/brainsci12010094.

[286] Y. Khosla, P. Ramachandra, and N. Chaitra, ‘Detection of autistic individuals using facial images and deep learning’, in 2021 IEEE International Conference on Computation System and Information Technology for Sustainable Solutions (CSITSS), Dec. 2021, pp. 1–5. doi: 10.1109/CSITSS54238.2021.9683205.

[287] R. Sadik, S. Anwar, and M. L. Reza, ‘AutismNet: Recognition of Autism Spectrum Disorder from Facial Expressions using MobileNet Architecture’, International Journal of Advanced Trends in Computer Science and Engineering, vol. 10, no. 1, pp. 327–334, Feb. 2021, doi: 10.30534/ijatcse/2021/471012021.

[288] B. Banire, D. Al Thani, M. Qaraqe, and B. Mansoor, ‘Face-Based Attention Recognition Model for Children with Autism Spectrum Disorder’, J Healthc Inform Res, vol. 5, no. 4, pp. 420–445, Dec. 2021, doi: 10.1007/s41666-021-00101-y.

[289] B. Li et al., ‘A Facial Affect Analysis System for Autism Spectrum Disorder’, in 2019 IEEE International Conference on Image Processing (ICIP), Sep. 2019, pp. 4549– 4553. doi: 10.1109/ICIP.2019.8803604.

[290] D.-Y. Song, C.-C. Topriceanu, D. C. Ilie-Ablachim, M. Kinali, and S. Bisdas, ‘Machine learning with neuroimaging data to identify autism spectrum disorder: a systematic review and meta-analysis’, Neuroradiology, vol. 63, no. 12, pp. 2057– 2072, Dec. 2021, doi: 10.1007/s00234-021-02774-z.

[291] K. Gao, Y. Sun, S. Niu, and L. Wang, ‘Unified framework for early stage status prediction of autism based on infant structural magnetic resonance imaging’, Autism Research, vol. 14, no. 12, pp. 2512–2523, Dec. 2021, doi: 10.1002/aur.2626.

[292] L. Q. Uddin et al., ‘Multivariate Searchlight Classification of Structural Magnetic Resonance Imaging in Children and Adolescents with Autism’, Biol Psychiatry, vol. 70, no. 9, pp. 833–841, Nov. 2011, doi: 10.1016/j.biopsych.2011.07.014.

[293] Y. Kong, J. Gao, Y. Xu, Y. Pan, J. Wang, and J. Liu, ‘Classification of autism spectrum disorder by combining brain connectivity and deep neural network classifier’, Neurocomputing, vol. 324, pp. 63–68, Jan. 2019, doi: 10.1016/j.neucom.2018.04.080.

[294] J. M. Górriz et al., ‘A Machine Learning Approach to Reveal the NeuroPhenotypes of Autisms’, Int J Neural Syst, vol. 29, no. 07, p. 1850058, Sep. 2019, doi: 10.1142/S0129065718500582.

[295] W. Sato et al., ‘Reduced Gray Matter Volume in the Social Brain Network in Adults with Autism Spectrum Disorder’, Front Hum Neurosci, vol. 11, p. 395, Aug. 2017, doi: 10.3389/fnhum.2017.00395.

[296] F. Segovia et al., ‘Identifying endophenotypes of autism: a multivariate approach’, Front Comput Neurosci, vol. 8, p. 60, Jun. 2014, doi: 10.3389/fncom.2014.00060.

[297] C. Ecker et al., ‘Investigating the predictive value of whole-brain structural MR scans in autism: A pattern classification approach’, Neuroimage, vol. 49, no. 1, pp. 44–56, Jan. 2010, doi: 10.1016/j.neuroimage.2009.08.024.

[298] L. Wang, C.-Y. Wee, X. Tang, P.-T. Yap, and D. Shen, ‘Multi-task feature selection via supervised canonical graph matching for diagnosis of autism spectrum disorder’, Brain Imaging Behav, vol. 10, no. 1, pp. 33–40, Mar. 2016, doi: 10.1007/s11682-015-9360-1.

[299] N. Denier, G. Steinberg, L. T. Elst, and T. Bracht, ‘The role of head circumference and cerebral volumes to phenotype male adults with autism spectrum disorder’, Brain Behav, vol. 12, no. 3, p. e2460, Mar. 2022, doi: 10.1002/brb3.2460.

[300] S. Vigneshwaran, B. S. Mahanand, S. Suresh, and R. Savitha, ‘Autism spectrum disorder detection using projection based learning meta-cognitive RBF network’, in The 2013 International Joint Conference on Neural Networks (IJCNN), Aug. 2013, pp. 1–8. doi: 10.1109/IJCNN.2013.6706777.

[301] M. J. Leming, S. Baron-Cohen, and J. Suckling, ‘Single-participant structural similarity matrices lead to greater accuracy in classification of participants than function in autism in MRI’, Mol Autism, vol. 12, no. 1, p. 34, Dec. 2021, doi: 10.1186/s13229-021-00439-5.

[302] Z. Fan, J. Su, K. Gao, D. Hu, and L.-L. Zeng, ‘A Federated Deep Learning Framework for 3D Brain MRI Images’, in 2021 International Joint Conference on Neural Networks (IJCNN), Jul. 2021, pp. 1–6. doi: 10.1109/IJCNN52387.2021.9534376.

[303] V. Subbaraju, S. Sundaram, S. Narasimhan, and M. B. Suresh, ‘Accurate detection of autism spectrum disorder from structural MRI using extended metacognitive radial basis function network’, Expert Syst Appl, vol. 42, no. 22, pp. 8775–8790, Dec. 2015, doi: 10.1016/j.eswa.2015.07.031.

[304] S. Calderoni, A. Retico, L. Biagi, R. Tancredi, F. Muratori, and M. Tosetti, ‘Female children with autism spectrum disorder: An insight from mass-univariate and pattern classification analyses’, Neuroimage, vol. 59, no. 2, pp. 1013–1022, Jan. 2012, doi: 10.1016/j.neuroimage.2011.08.070.

[305] Y. Fu et al., ‘A novel pipeline leveraging surface-based features of small subcortical structures to classify individuals with autism spectrum disorder’, Prog Neuropsychopharmacol Biol Psychiatry, vol. 104, p. 109989, Jan. 2021, doi: 10.1016/j.pnpbp.2020.109989.

[306] I. Bilgen, G. Guvercin, and I. Rekik, ‘Machine learning methods for brain network classification: Application to autism diagnosis using cortical morphological networks’, J Neurosci Methods, vol. 343, p. 108799, Sep. 2020, doi: 10.1016/j.jneumeth.2020.108799.

[307] O. Graa and I. Rekik, ‘Multi-view learning-based data proliferator for boosting classification using highly imbalanced classes’, J Neurosci Methods, vol. 327, p. 108344, Nov. 2019, doi: 10.1016/j.jneumeth.2019.108344.

[308] C. Ecker et al., ‘Describing the Brain in Autism in Five Dimensions--Magnetic Resonance Imaging-Assisted Diagnosis of Autism Spectrum Disorder Using a Multiparameter Classification Approach’, Journal of Neuroscience, vol. 30, no. 32, pp. 10612–10623, Aug. 2010, doi: 10.1523/JNEUROSCI.5413-09.2010.

[309] Y. Jiao, R. Chen, X. Ke, K. Chu, Z. Lu, and E. H. Herskovits, ‘Predictive models of autism spectrum disorder based on brain regional cortical thickness’, Neuroimage, vol. 50, no. 2, pp. 589–599, Apr. 2010, doi: 10.1016/j.neuroimage.2009.12.047.

[310] L. Squarcina et al., ‘Automatic classification of autism spectrum disorder in children using cortical thickness and support vector machine’, Brain Behav, vol. 11, no. 8, p. e2238, Aug. 2021, doi: 10.1002/brb3.2238.

[311] A. Demirhan, ‘The effect of feature selection on multivariate pattern analysis of structural brain MR images’, Physica Medica, vol. 47, pp. 103–111, Mar. 2018, doi: 10.1016/j.ejmp.2018.03.002.

[312] S. Tummala, ‘Deep Learning Framework using Siamese Neural Network for Diagnosis of Autism from Brain Magnetic Resonance Imaging’, in 2021 6th International Conference for Convergence in Technology (I2CT), Apr. 2021, pp. 1–5. doi: 10.1109/I2CT51068.2021.9418143.

[313] R. Nur Syahindah Husna, A. R. Syafeeza, N. Abdul Hamid, Y. C. Wong, and R. Atikah Raihan, ‘Functional Magnetic Resonance Imaging for Autism Spectrum Disorder Detection Using Deep Learning’, J Teknol, vol. 83, no. 3, pp. 45–52, Apr. 2021, doi: 10.11113/jurnalteknologi.v83.16389.

[314] X. Guo et al., ‘Diagnosing autism spectrum disorder in children using conventional MRI and apparent diffusion coefficient based deep learning algorithms’, Eur Radiol, vol. 32, no. 2, pp. 761–770, Feb. 2022, doi: 10.1007/s00330-021-08239-4.

[315] H. Shahamat and M. Saniee Abadeh, ‘Brain MRI analysis using a deep learning based evolutionary approach’, Neural Networks, vol. 126, pp. 218–234, Jun. 2020, doi: 10.1016/j.neunet.2020.03.017.

[316] X. Chen, Z. Wang, F. A. Cheikh, and M. Ullah, ‘3D-Resnet Fused Attention for Autism Spectrum Disorder Classification’, in Image and Graphics, 2021, pp. 607– 617. doi: 10.1007/978-3-030-87358-5_49.

[317] F. Ke, S. Choi, Y. H. Kang, K.-A. Cheon, and S. W. Lee, ‘Exploring the Structural and Strategic Bases of Autism Spectrum Disorders With Deep Learning’, IEEE Access, vol. 8, pp. 153341–153352, 2020, doi: 10.1109/ACCESS.2020.3016734.

[318] C.-Y. Wee, L. Wang, F. Shi, P.-T. Yap, and D. Shen, ‘Diagnosis of autism spectrum disorders using regional and interregional morphological features’, Hum Brain Mapp, vol. 35, no. 7, pp. 3414–3430, Jul. 2014, doi: 10.1002/hbm.22411.

[319] S. Haar, S. Berman, M. Behrmann, and I. Dinstein, ‘Anatomical Abnormalities in Autism?’, Cerebral Cortex, vol. 26, no. 4, pp. 1440–1452, Apr. 2016, doi: 10.1093/cercor/bhu242.

[320] W. H. L. Pinaya, A. Mechelli, and J. R. Sato, ‘Using deep autoencoders to identify abnormal brain structural patterns in neuropsychiatric disorders: A large-scale multi-sample study’, Hum Brain Mapp, vol. 40, no. 3, pp. 944–954, Feb. 2019, doi: 10.1002/hbm.24423.

[321] E. P. K. Pua, G. Ball, C. Adamson, S. Bowden, and M. L. Seal, ‘Quantifying individual differences in brain morphometry underlying symptom severity in Autism Spectrum Disorders’, Sci Rep, vol. 9, no. 1, p. 9898, Dec. 2019, doi: 10.1038/s41598-019-45774-z.

[322] G. J. Katuwal, N. D. Cahill, S. A. Baum, and A. M. Michael, ‘The predictive power of structural MRI in Autism diagnosis’, in 2015 37th Annual International Conference of the IEEE Engineering in Medicine and Biology Society (EMBC), Aug. 2015, pp. 4270–4273. doi: 10.1109/EMBC.2015.7319338.

[323] M. Mishra and U. C. Pati, ‘Autism Spectrum Disorder Detection using Surface Morphometric Feature of sMRI in Machine Learning’, in 2021 8th International Conference on Smart Computing and Communications (ICSCC), Jul. 2021, pp. 17–20. doi: 10.1109/ICSCC51209.2021.9528240.

[324] M. T. Ali et al., ‘Autism Classification Using SMRI: A Recursive Features Selection Based on Sampling from Multi-Level High Dimensional Spaces’, in 2021 IEEE 18th International Symposium on Biomedical Imaging (ISBI), Apr. 2021, pp. 267–270. doi: 10.1109/ISBI48211.2021.9433973.

[325] H. Sharif and R. A. Khan, ‘A novel machine learning based framework for detection of Autism Spectrum Disorder (ASD)’, arXiv preprint arXiv:1903.11323, Mar. 2019, doi: 10.1080/08839514.2021.2004655.

[326] H. C. Hazlett et al., ‘Early brain development in infants at high risk for autism spectrum disorder’, Nature, vol. 542, no. 7641, pp. 348–351, Feb. 2017, doi: 10.1038/nature21369.

[327] M. T. Ali et al., ‘The Role of Structure MRI in Diagnosing Autism’, Diagnostics, vol. 12, no. 1, p. 165, Jan. 2022, doi: 10.3390/diagnostics12010165.

[328] M. R. Sabuncu and E. Konukoglu, ‘Clinical Prediction from Structural Brain MRI Scans: A Large-Scale Empirical Study’, Neuroinformatics, vol. 13, no. 1, pp. 31–46, Jan. 2015, doi: 10.1007/s12021-014-9238-1.

[329] E. Ferrari et al., ‘Dealing with confounders and outliers in classification medical studies: The Autism Spectrum Disorders case study’, Artif Intell Med, vol. 108, p. 101926, Aug. 2020, doi: 10.1016/j.artmed.2020.101926.

[330] I. Gori et al., ‘Gray Matter Alterations in Young Children with Autism Spectrum Disorders: Comparing Morphometry at the Voxel and Regional Level’, Journal of Neuroimaging, vol. 25, no. 6, pp. 866–874, Nov. 2015, doi: 10.1111/jon.12280.

[331] J. Germann et al., ‘Involvement of the habenula in the pathophysiology of autism spectrum disorder’, Sci Rep, vol. 11, no. 1, p. 21168, Dec. 2021, doi: 10.1038/s41598-021-00603-0.

[332] G. Li, M. Liu, Q. Sun, D. Shen, and L. Wang, ‘Early Diagnosis of Autism Disease by Multi-channel CNNs’, in Mach Learn Med Imaging, vol. 11046, 2018, pp. 303–309. doi: 10.1007/978-3-030-00919-9_35.

[333] T. Chen et al., ‘The Development of a Practical Artificial Intelligence Tool for Diagnosing and Evaluating Autism Spectrum Disorder: Multicenter Study’, JMIR Med Inform, vol. 8, no. 5, p. e15767, May 2020, doi: 10.2196/15767.

[334] X. Zhang et al., ‘Siamese Verification Framework for Autism Identification During Infancy Using Cortical Path Signature Features’, in 2020 IEEE 17th International Symposium on Biomedical Imaging (ISBI), Apr. 2020, pp. 1–4. doi: 10.1109/ISBI45749.2020.9098385.

[335] C. Alvarez-Jimenez, N. Múnera-Garzón, M. A. Zuluaga, N. F. Velasco, and E. Romero, ‘Autism spectrum disorder characterization in children by capturing local-regional brain changes in MRI’, Med Phys, vol. 47, no. 1, pp. 119–131, Jan. 2020, doi: 10.1002/mp.13901.

[336] E. Varol, B. Gaonkar, G. Erus, R. Schultz, and C. Davatzikos, ‘Feature ranking based nested support vector machine ensemble for medical image classification’, in 2012 9th IEEE International Symposium on Biomedical Imaging (ISBI), May 2012, pp. 146–149. doi: 10.1109/ISBI.2012.6235505.

[337] R. Yang, F. Ke, H. Liu, M. Zhou, and H.-M. Cao, ‘Exploring sMRI Biomarkers for Diagnosis of Autism Spectrum Disorders Based on Multi Class Activation Mapping Models’, IEEE Access, vol. 9, pp. 124122–124131, 2021, doi: 10.1109/ACCESS.2021.3069211.

[338] J. Gao et al., ‘Multisite Autism Spectrum Disorder Classification Using Convolutional Neural Network Classifier and Individual Morphological Brain Networks’, Front Neurosci, vol. 14, p. 629630, Jan. 2021, doi: 10.3389/fnins.2020.629630.

[339] M. Madine, I. Rekik, and N. Werghi, ‘Diagnosing Autism Using T1-W MRI With Multi-Kernel Learning and Hypergraph Neural Network’, in 2020 IEEE International Conference on Image Processing (ICIP), Oct. 2020, pp. 438–442. doi: 10.1109/ICIP40778.2020.9190924.

[340] M. Soussia and I. Rekik, ‘Unsupervised Manifold Learning Using High-Order Morphological Brain Networks Derived From T1-w MRI for Autism Diagnosis’, Front Neuroinform, vol. 12, p. 70, Oct. 2018, doi: 10.3389/fninf.2018.00070.

[341] N. Georges, I. Mhiri, and I. Rekik, ‘Identifying the best data-driven feature selection method for boosting reproducibility in classification tasks’, Pattern Recognit, vol. 101, p. 107183, May 2020, doi: 10.1016/j.patcog.2019.107183.

[342] G. Chanel, S. Pichon, L. Conty, S. Berthoz, C. Chevallier, and J. Grèzes, ‘Classification of autistic individuals and controls using cross-task characterization of fMRI activity’, Neuroimage Clin, vol. 10, pp. 78–88, 2016, doi: 10.1016/j.nicl.2015.11.010.

[343] P. Odriozola, L. Q. Uddin, C. J. Lynch, J. Kochalka, T. Chen, and V. Menon, ‘Insula response and connectivity during social and non-social attention in children with autism’, Soc Cogn Affect Neurosci, vol. 11, no. 3, pp. 433–444, Mar. 2016, doi: 10.1093/scan/nsv126.

[344] M. A. Just, V. L. Cherkassky, A. Buchweitz, T. A. Keller, and T. M. Mitchell, ‘Identifying Autism from Neural Representations of Social Interactions: Neurocognitive Markers of Autism’, PLoS One, vol. 9, no. 12, p. e113879, Dec. 2014, doi: 10.1371/journal.pone.0113879.

[345] X. Li, N. C. Dvornek, J. Zhuang, P. Ventola, and J. S. Duncan, ‘Brain Biomarker Interpretation in ASD Using Deep Learning and fMRI’, in Med Image Comput Comput Assist Interv, vol. 11072, 2018, pp. 206–214. doi: 10.1007/978-3-030-00931-1_24.

[346] X. Li et al., ‘BrainGNN: Interpretable Brain Graph Neural Network for fMRI Analysis’, Med Image Anal, vol. 74, p. 102233, Dec. 2021, doi: 10.1016/j.media.2021.102233.

[347] X. Li, N. C. Dvornek, Y. Zhou, J. Zhuang, P. Ventola, and J. S. Duncan, ‘Graph Neural Network for Interpreting Task-fMRI Biomarkers’, in Med Image Comput Comput Assist Interv, vol. 11768, 2019, pp. 485–493. doi: 10.1007/978-3-030-32254-0_54.

[348] X. Li et al., ‘2-Channel convolutional 3D deep neural network (2CC3D) for fMRI analysis: ASD classification and feature learning’, in 2018 IEEE 15th International Symposium on Biomedical Imaging (ISBI 2018), Apr. 2018, pp. 1252–1255. doi: 10.1109/ISBI.2018.8363798.

[349] N. C. Dvornek, D. Yang, P. Ventola, and J. S. Duncan, ‘Learning Generalizable Recurrent Neural Networks from Small Task-fMRI Datasets’, in Med Image Comput Comput Assist Interv, vol. 11072, 2018, pp. 329–337. doi: 10.1007/978-3-030-00931-1_38.

[350] D. L. Murdaugh, S. V. Shinkareva, H. R. Deshpande, J. Wang, M. R. Pennick, and R. K. Kana, ‘Differential Deactivation during Mentalizing and Classification of Autism Based on Default Mode Network Connectivity’, PLoS One, vol. 7, no. 11, p. e50064, Nov. 2012, doi: 10.1371/journal.pone.0050064.

[351] H. Wang, C. Chen, and H. Fushing, ‘Extracting Multiscale Pattern Information of fMRI Based Functional Brain Connectivity with Application on Classification of Autism Spectrum Disorders’, PLoS One, vol. 7, no. 10, p. e45502, Oct. 2012, doi: 10.1371/journal.pone.0045502.

[352] R. Haweel et al., ‘Functional Magnetic Resonance Imaging Based Framework for Autism Diagnosis’, in 2019 Fifth International Conference on Advances in Biomedical Engineering (ICABME), Oct. 2019, pp. 1–4. doi: 10.1109/ICABME47164.2019.8940348.

[353] R. Haweel et al., ‘A robust DWT–CNN-based CAD system for early diagnosis of autism using task-based fMRI’, Med Phys, vol. 48, no. 5, pp. 2315–2326, May 2021, doi: 10.1002/mp.14692.

[354] R. Haweel et al., ‘A Novel DWT-Based Discriminant Features Extraction From Task-Based Fmri: An Asd Diagnosis Study Using Cnn’, in 2021 IEEE 18th International Symposium on Biomedical Imaging (ISBI), Apr. 2021, pp. 196–199. doi: 10.1109/ISBI48211.2021.9433768.

[355] R. Haweel, N. Seada, S. Ghoniemy, N. S. Alghamdi, and A. El-Baz, ‘A CNN Deep Local and Global ASD Classification Approach with Continuous Wavelet Transform Using Task-Based FMRI’, Sensors, vol. 21, no. 17, p. 5822, Aug. 2021, doi: 10.3390/s21175822.

[356] L. Xu et al., ‘Characterizing autism spectrum disorder by deep learning spontaneous brain activity from functional near-infrared spectroscopy’, J Neurosci Methods, vol. 331, p. 108538, Feb. 2020, doi: 10.1016/j.jneumeth.2019.108538.

[357] L. Xu, X. Geng, X. He, J. Li, and J. Yu, ‘Prediction in Autism by Deep Learning Short-Time Spontaneous Hemodynamic Fluctuations’, Front Neurosci, vol. 13, p. 1120, Nov. 2019, doi: 10.3389/fnins.2019.01120.

[358] J. Li, L. Qiu, L. Xu, E. V. Pedapati, C. A. Erickson, and U. Sunar, ‘Characterization of autism spectrum disorder with spontaneous hemodynamic activity’, Biomed Opt Express, vol. 7, no. 10, p. 3871, Oct. 2016, doi: 10.1364/BOE.7.003871.

[359] A. Crippa et al., ‘Use of Machine Learning to Identify Children with Autism and Their Motor Abnormalities’, J Autism Dev Disord, vol. 45, no. 7, pp. 2146–2156, Jul. 2015, doi: 10.1007/s10803-015-2379-8.

[360] Z. Zhao et al., ‘Identifying Autism with Head Movement Features by Implementing Machine Learning Algorithms’, J Autism Dev Disord, vol. 52, no. 7, pp. 3038–3049, Jul. 2022, doi: 10.1007/s10803-021-05179-2.

[361] Z. Zhao et al., ‘Applying Machine Learning to Identify Autism With Restricted Kinematic Features’, IEEE Access, vol. 7, pp. 157614–157622, 2019, doi: 10.1109/ACCESS.2019.2950030.

[362] W. Li, Z. Wang, L. Zhang, L. Qiao, and D. Shen, ‘Remodeling Pearson’s Correlation for Functional Brain Network Estimation and Autism Spectrum Disorder Identification’, Front Neuroinform, vol. 11, p. 55, Aug. 2017, doi: 10.3389/fninf.2017.00055.

[363] A. Anzulewicz, K. Sobota, and J. T. Delafield-Butt, ‘Toward the Autism Motor Signature: Gesture patterns during smart tablet gameplay identify children with autism’, Sci Rep, vol. 6, no. 1, p. 31107, Aug. 2016, doi: 10.1038/srep31107.

[364] W. Liu, T. Zhou, C. Zhang, X. Zou, and M. Li, ‘Response to name: A dataset and a multimodal machine learning framework towards autism study’, in 2017 Seventh International Conference on Affective Computing and Intelligent Interaction (ACII), Oct. 2017, pp. 178–183. doi: 10.1109/ACII.2017.8273597.

[365] Q. Tariq, J. Daniels, J. N. Schwartz, P. Washington, H. Kalantarian, and D. P. Wall, ‘Mobile detection of autism through machine learning on home video: A development and prospective validation study’, PLoS Med, vol. 15, no. 11, p. e1002705, Nov. 2018, doi: 10.1371/journal.pmed.1002705.

[366] C. Wu et al., ‘Machine Learning Based Autism Spectrum Disorder Detection from Videos’, in 2020 *IEEE International Conference on E-health Networking*, Application & Services (HEALTHCOM), Mar. 2021, vol. 2020, pp. 1–6. doi: 10.1109/HEALTHCOM49281.2021.9398924.

[367] M. Ruan, ‘Image and Video-Based Autism Spectrum Disorder Detection via Deep Learning’, Benjamin M. Statler College of Engineering and Mineral Resources, Morgantown, West Virginia, 2020.

[368] M. J. Maenner, M. Yeargin-Allsopp, K. Van Naarden Braun, D. L. Christensen, and L. A. Schieve, ‘Development of a Machine Learning Algorithm for the Surveillance of Autism Spectrum Disorder’, PLoS One, vol. 11, no. 12, p. e0168224, Dec. 2016, doi: 10.1371/journal.pone.0168224.

[369] E. Duchesnay et al., ‘Feature selection and classification of imbalanced datasets’, Neuroimage, vol. 57, no. 3, pp. 1003–1014, Aug. 2011, doi: 10.1016/j.neuroimage.2011.05.011.

[370] S. Liu, F. Ge, L. Zhao, T. Wang, D. Ni, and T. Liu, ‘NAS-optimized topology-preserving transfer learning for differentiating cortical folding patterns’, Med Image Anal, vol. 77, p. 102316, Apr. 2022, doi: 10.1016/j.media.2021.102316.

[371] M. Rakić, M. Cabezas, K. Kushibar, A. Oliver, and X. Lladó, ‘Improving the detection of autism spectrum disorder by combining structural and functional MRI information’, Neuroimage Clin, vol. 25, p. 102181, 2020, doi: 10.1016/j.nicl.2020.102181.

[372] O. Dekhil et al., ‘A Personalized Autism Diagnosis CAD System Using a Fusion of Structural MRI and Resting-State Functional MRI Data’, Front Psychiatry, vol. 10, p. 392, Jul. 2019, doi: 10.3389/fpsyt.2019.00392.

[373] S. Itani and D. Thanou, ‘Combining anatomical and functional networks for neuropathology identification: A case study on autism spectrum disorder’, Med Image Anal, vol. 69, p. 101986, Apr. 2021, doi: 10.1016/j.media.2021.101986.

[374] O. Dekhil et al., ‘A Comprehensive Framework for Differentiating Autism Spectrum Disorder From Neurotypicals by Fusing Structural MRI and Resting State Functional MRI’, Semin Pediatr Neurol, vol. 34, p. 100805, Jul. 2020, doi: 10.1016/j.spen.2020.100805.

[375] Y. Chen et al., ‘Attention-Based Node-Edge Graph Convolutional Networks for Identification of Autism Spectrum Disorder Using Multi-Modal MRI Data’, in Pattern Recognition and Computer Vision, 2021, pp. 374–385. doi: 10.1007/978-3-030-88010-1_31.

[376] Y. Zhou, F. Yu, and T. Duong, ‘Multiparametric MRI Characterization and Prediction in Autism Spectrum Disorder Using Graph Theory and Machine Learning’, PLoS One, vol. 9, no. 6, p. e90405, Jun. 2014, doi: 10.1371/journal.pone.0090405.

[377] M. Akhavan Aghdam, A. Sharifi, and M. M. Pedram, ‘Combination of rs-fMRI and sMRI Data to Discriminate Autism Spectrum Disorders in Young Children Using Deep Belief Network’, J Digit Imaging, vol. 31, no. 6, pp. 895–903, Dec. 2018, doi: 10.1007/s10278-018-0093-8.

[378] B. Sen, N. C. Borle, R. Greiner, and M. R. G. Brown, ‘A general prediction model for the detection of ADHD and Autism using structural and functional MRI’, PLoS One, vol. 13, no. 4, p. e0194856, Apr. 2018, doi: 10.1371/journal.pone.0194856.

[379] K. M and S. Jaganathan, ‘Graph Convolutional Model to Diagnose Autism Spectrum Disorder Using Rs-Fmri Data’, in 2021 *5th International Conference on Computer*, Communication and Signal Processing (ICCCSP), May 2021, pp. 1–5. doi: 10.1109/ICCCSP52374.2021.9465490.

[380] A. Brahim and N. Farrugia, ‘Graph Fourier transform of fMRI temporal signals based on an averaged structural connectome for the classification of neuroimaging’, Artif Intell Med, vol. 106, p. 101870, Jun. 2020, doi: 10.1016/j.artmed.2020.101870.

[381] A. Eill et al., ‘Functional Connectivities Are More Informative Than Anatomical Variables in Diagnostic Classification of Autism’, Brain Connect, vol. 9, no. 8, pp. 604–612, Oct. 2019, doi: 10.1089/brain.2019.0689.

[382] A. Crimi, L. Dodero, V. Murino, and D. Sona, ‘Case-control discrimination through effective brain connectivity’, in 2017 IEEE 14th International Symposium on Biomedical Imaging (ISBI 2017), Apr. 2017, pp. 970–973. doi: 10.1109/ISBI.2017.7950677.

[383] G. Deshpande, L. E. Libero, K. R. Sreenivasan, H. D. Deshpande, and R. K. Kana, ‘Identification of neural connectivity signatures of autism using machine learning’, Front Hum Neurosci, vol. 7, p. 670, 2013, doi: 10.3389/fnhum.2013.00670.

[384] A. Irimia, X. Lei, C. M. Torgerson, Z. J. Jacokes, S. Abe, and J. D. Van Horn, ‘Support Vector Machines, Multidimensional Scaling and Magnetic Resonance Imaging Reveal Structural Brain Abnormalities Associated With the Interaction Between Autism Spectrum Disorder and Sex’, Front Comput Neurosci, vol. 12, p. 93, Nov. 2018, doi: 10.3389/fncom.2018.00093.

[385] J. I. Kim et al., ‘Classification of Preschoolers with Low-Functioning Autism Spectrum Disorder Using Multimodal MRI Data’, J Autism Dev Disord, Jan. 2022, doi: 10.1007/s10803-021-05368-z.

[386] C. J. Goch et al., ‘Global Changes in the Connectome in Autism Spectrum Disorders’, in *Computational Diffusion MRI and Brain Connectivity*, Springer, 2014, pp. 239–247. doi: 10.1007/978-3-319-02475-2_22.

[387] S. Payabvash et al., ‘White Matter Connectome Edge Density in Children with Autism Spectrum Disorders: Potential Imaging Biomarkers Using Machine-Learning Models’, Brain Connect, vol. 9, no. 2, pp. 209–220, Mar. 2019, doi: 10.1089/brain.2018.0658.

[388] S. Thapaliya, S. Jayarathna, and M. Jaime, ‘Evaluating the EEG and Eye Movements for Autism Spectrum Disorder’, in 2018 IEEE International Conference on Big Data (Big Data), Dec. 2018, pp. 2328–2336. doi: 10.1109/BigData.2018.8622501.

[389] S. Zhang, D. Chen, Y. Tang, and L. Zhang, ‘Children ASD Evaluation Through Joint Analysis of EEG and Eye-Tracking Recordings With Graph Convolution Network’, Front Hum Neurosci, vol. 15, p. 651349, May 2021, doi: 10.3389/fnhum.2021.651349.

[390] J. Kang, X. Han, J. Song, Z. Niu, and X. Li, ‘The identification of children with autism spectrum disorder by SVM approach on EEG and eye-tracking data’, Comput Biol Med, vol. 120, p. 103722, May 2020, doi: 10.1016/j.compbiomed.2020.103722.

[391] P. Lu, X. Li, L. Hu, and L. Lu, ‘Integrating genomic and resting State fMRI for efficient autism spectrum disorder classification’, Multimed Tools Appl, vol. 81, no. 14, pp. 19183–19194, Jun. 2022, doi: 10.1007/s11042-020-10473-9.

[392] A. Vabalas, E. Gowen, E. Poliakoff, and A. J. Casson, ‘Applying Machine Learning to Kinematic and Eye Movement Features of a Movement Imitation Task to Predict Autism Diagnosis’, Sci Rep, vol. 10, no. 1, p. 8346, Dec. 2020, doi: 10.1038/s41598-020-65384-4.

[393] L. E. Libero, T. P. DeRamus, A. C. Lahti, G. Deshpande, and R. K. Kana, ‘Multimodal neuroimaging based classification of autism spectrum disorder using anatomical, neurochemical, and white matter correlates’, Cortex, vol. 66, pp. 46–59, May 2015, doi: 10.1016/j.cortex.2015.02.008.

[394] C. Cameron et al., ‘The Neuro Bureau Preprocessing Initiative: open sharing of preprocessed neuroimaging data and derivatives’, Front Neuroinform, vol. 7, 2013, doi: 10.3389/conf.fninf.2013.09.00041.

[395] A. Di Martino et al., ‘The autism brain imaging data exchange: towards a large-scale evaluation of the intrinsic brain architecture in autism’, Mol Psychiatry, vol. 19, no. 6, pp. 659–667, Jun. 2014, doi: 10.1038/mp.2013.78.

[396] D. Hall, M. F. Huerta, M. J. McAuliffe, and G. K. Farber, ‘Sharing Heterogeneous Data: The National Database for Autism Research’, Neuroinformatics, vol. 10, no. 4, pp. 331–339, Oct. 2012, doi: 10.1007/s12021-012-9151-4.

[397] Y. Sheu, ‘Illuminating the Black Box: Interpreting Deep Neural Network Models for Psychiatric Research’, Front Psychiatry, vol. 11, Oct. 2020, doi: 10.3389/fpsyt.2020.551299.

[398] Z. C. Lipton, ‘The mythos of model interpretability’, Commun ACM, vol. 61, no. 10, pp. 36–43, Sep. 2018, doi: 10.1145/3233231.

[399] R. Guidotti, A. Monreale, S. Ruggieri, F. Turini, F. Giannotti, and D. Pedreschi, ‘A Survey of Methods for Explaining Black Box Models’, ACM Comput Surv, vol. 51, no. 5, pp. 1–42, Sep. 2019, doi: 10.1145/3236009.

[400] F. Doshi-Velez and B. Kim, ‘Towards A Rigorous Science of Interpretable Machine Learning’, arXiv preprint arXiv:1702.08608, Feb. 2017.

[401] A. Barredo Arrieta et al., ‘Explainable Artificial Intelligence (XAI): Concepts, taxonomies, opportunities and challenges toward responsible AI’, Information Fusion, vol. 58, pp. 82–115, Jun. 2020, doi: 10.1016/j.inffus.2019.12.012.

[402] M. Kimura and M. Tanaka, ‘New Perspective of Interpretability of Deep Neural Networks’, in 2020 3rd International Conference on Information and Computer Technologies (ICICT), Mar. 2020, pp. 78–85. doi: 10.1109/ICICT50521.2020.00020.

[403] M. R. Arbabshirani, S. Plis, J. Sui, and V. D. Calhoun, ‘Single subject prediction of brain disorders in neuroimaging: Promises and pitfalls’, Neuroimage, vol. 145, pp. 137–165, Jan. 2017, doi: 10.1016/j.neuroimage.2016.02.079.

[404] A. Brooks, ‘How much does an MRI cost without insurance in 2023?’, Feb. 20, 2023. https://www.talktomira.com/post/how-much-does-an-mri-cost-without-insurance-in-2021

[405] M. Allen, ‘fMRI vs. SPECT scan for the brain: Know your options’, Jan. 21, 2023. https://www.cognitivefxusa.com/blog/fmri-vs-spect-scan-for-brain

[406] CostHelper, ‘How much does an EEG cost?’, Dec. 22, 2020. https://health.costhelper.com/eeg.html

[407] U. C. Mehta, I. Patel, and F. V. Castello, ‘EEG Sedation for Children with Autism’, Journal of Developmental & Behavioral Pediatrics, vol. 25, no. 2, pp. 102–104, Apr. 2004, doi: 10.1097/00004703-200404000-00005.

[408] X. Ying, ‘An Overview of Overfitting and its Solutions’, J Phys Conf Ser, vol. 1168, p. 022022, Feb. 2019, doi: 10.1088/1742-6596/1168/2/022022.

[409] T. Hernandez-Boussard, S. Bozkurt, J. P. A. Ioannidis, and N. H. Shah, ‘MINIMAR (MINimum Information for Medical AI Reporting): Developing reporting standards for artificial intelligence in health care’, Journal of the American Medical Informatics Association, vol. 27, no. 12, pp. 2011–2015, Dec. 2020, doi: 10.1093/jamia/ocaa088.

[410] J. Mongan, L. Moy, and C. E. Kahn, ‘Checklist for Artificial Intelligence in Medical Imaging (CLAIM): A Guide for Authors and Reviewers’, Radiol Artif Intell, vol. 2, no. 2, p. e200029, Mar. 2020, doi: 10.1148/ryai.2020200029.

[411] V. Sounderajah et al., ‘Developing a reporting guideline for artificial intelligence-centred diagnostic test accuracy studies: the STARD-AI protocol’, BMJ Open, vol. 11, no. 6, p. e047709, Jun. 2021, doi: 10.1136/bmjopen-2020-047709.

[412] C. Meng, L. Trinh, N. Xu, J. Enouen, and Y. Liu, ‘Interpretability and fairness evaluation of deep learning models on MIMIC-IV dataset’, Sci Rep, vol. 12, no. 1, p. 7166, May 2022, doi: 10.1038/s41598-022-11012-2.

[413] R. Gadepally, ‘Overfitting and Generalization of AI in Medical Imaging’, Apr. 11, 2022. https://www.acrdsi.org/DSIBlog/2022/04/11/Overfitting-and-Generalization-of-AI-in-Medical-Imaging

[414] M. D. Wilkinson et al., ‘The FAIR Guiding Principles for scientific data management and stewardship’, Sci Data, vol. 3, no. 1, p. 160018, Mar. 2016, doi: 10.1038/sdata.2016.18.

[415] M. D. Kohli, R. M. Summers, and J. R. Geis, ‘Medical Image Data and Datasets in the Era of Machine Learning—Whitepaper from the 2016 C-MIMI Meeting Dataset Session’, J Digit Imaging, vol. 30, no. 4, pp. 392–399, Aug. 2017, doi: 10.1007/s10278-017-9976-3.

[416] N. Kela-Madar and I. Kela, ‘The Machine-Human Collaboration in Healthcare Innovation’, in *Toward Super-Creativity -Improving Creativity in Humans, Machines, and Human -Machine Collaborations*, IntechOpen, 2020. doi: 10.5772/intechopen.88951.

[417] V. Volovici, N. L. Syn, A. Ercole, J. J. Zhao, and N. Liu, ‘Steps to avoid overuse and misuse of machine learning in clinical research’, Nat Med, vol. 28, no. 10, pp. 1996– 1999, Oct. 2022, doi: 10.1038/s41591-022-01961-6.

