## Supplementary material for "Automated diagnosis of autism: State of the art": Multimedia Appendix 1: Search strategy

Embase (Elsevier) <1947 to Feb 2022>

#1 ‘autism’/exp
#2 'asperger syndrom*':ab,ti,kw OR 'autis*':ab,ti,kw OR 'child* development* disorder*':ab,ti,kw
#3 #1 OR #2
#4 'artificial intelligence*':ab,ti,kw OR 'deep learning*':ab,ti,kw OR 'machine learning*':ab,ti,kw OR 'machine intelligence*':ab,ti,kw OR 'supervised learning*':ab,ti,kw OR 'unsupervised learning*':ab,ti,kw OR ‘semi-supervised learning*’:ab,ti,kw OR ‘reinforcement learning*’:ab,ti,kw OR 'regression algorithm*':ab,ti,kw OR 'learning vector quantization*':ab,ti,kw OR 'self organizing map*':ab,ti,kw OR 'regularization algorithm*':ab,ti,kw OR 'iterative dichotomiser*':ab,ti,kw OR 'decision stump*':ab,ti,kw OR 'clustering algorithm*':ab,ti,kw OR 'k-means*':ab,ti,kw OR 'k-medians*':ab,ti,kw OR 'hierarchical clustering*':ab,ti,kw OR 'apriori*':ab,ti,kw OR 'auto encoder*':ab,ti,kw OR 'autoencoder*':ab,ti,kw OR 'markov*':ab,ti,kw OR 'classification algorithm*':ab,ti,kw OR 'locally weighted learning*':ab,ti,kw OR 'support vector machine*':ab,ti,kw OR 'svm':ab,ti,kw OR 'decision tree*':ab,ti,kw OR 'component analy*':ab,ti,kw OR 'pca':ab,ti,kw OR 'ica':ab,ti,kw OR 'neural network*':ab,ti,kw OR 'hierarchical learning*':ab,ti,kw OR 'latent class analy*':ab,ti,kw OR 'latent class model*':ab,ti,kw OR 'latent variable model*':ab,ti,kw OR 'boltzmann':ab,ti,kw OR 'deep belief network*':ab,ti,kw OR ‘lasso regression*':ab,ti,kw OR ‘lda':ab,ti,kw OR ‘adtree':ab,ti,kw OR ‘ridge regression':ab,ti,kw OR ‘enet':ab,ti,kw OR ‘fnn':ab,ti,kw OR ‘firefly algorithm*':ab,ti,kw OR ‘multivariate adaptive regression splines*’:ab,ti,kw OR ‘locally estimated scatterplot smoothing*’:ab,ti,kw OR ‘instance base*’:ab,ti,kw OR ‘nearest neighbor*’:ab,ti,kw OR ‘regularization algorithm*’:ab,ti,kw OR ‘least absolute shrinkage select*’:ab,ti,kw OR ‘least angle regression*’:ab,ti,kw OR ‘eclat*’:ab,ti,kw OR ‘back propagation*’:ab,ti,kw OR ‘hopfield network*’:ab,ti,kw OR ‘radial basis function network*’:ab,ti,kw OR ‘dimensionality reduction algorithm*’:ab,ti,kw OR ‘ensemble algorithm*’:ab,ti,kw OR ‘adaboost’:ab,ti,kw OR ‘stacked generalization*’:ab,ti,kw OR ‘relu’:ab,ti,kw OR ‘naive bayes’:ab,ti,kw
#5 #3 AND #4

MEDLINE (OvidSP) <1996 to Feb 2022>

1 exp Autism Spectrum Disorder/
2 (asperger syndrom* or autis* or child* development* disorder*).mp.
3 1 or 2
4 exp Algorithms/ or exp Neural Networks, Computer/ or exp Decision Trees/
5 (artificial intelligence* or deep learning* or machine learning* or machine intelligence* or supervised learning* or unsupervised learning* or semi-supervised learning* or reinforcement learning* or regression algorithm* or learning vector quantization* or self organizing map* or regularization algorithm* or iterative dichotomiser* or decision stump* or clustering algorithm* or k-means* or k-medians* or hierarchical clustering* or apriori* or auto encoder* or auto-encoder* or markov* or classification algorithm* or locally weighted learning* or support vector machine* or svm or decision tree* or component analy* or pca or ica or neural network* or hierarchical learning* or latent class analy* or latent class model* or latent variable model* or boltzmann or deep belief network* or lasso regression* or lda or adtree or ridge regression or enet or fnn or firefly algorithm* or multivariate adaptive regression splines* or locally estimated

scatterplot smoothing* or instance base* or nearest neighbor* or regularization algorithm* or least absolute shrinkage select* or least angle regression* or eclat* or back propagation* or hopfield network* or radial basis function network* or dimensionality reduction algorithm* or ensemble algorithm* or adaboost or stacked generalization* or relu or naive bayes).mp.
6 4 or 5
7 3 and 6

APA PsycINFO (OvidSP) <1987 to Feb 2022>
1 exp Autism Spectrum Disorders/
2 (asperger syndrom* or autis* or child* development* disorder*).mp.
3 1 or 2
4 exp Artificial Intelligence/
5 (artificial intelligence* or deep learning* or machine learning* or machine intelligence* or supervised learning* or unsupervised learning* or semi-supervised learning* or reinforcement learning* or regression algorithm* or learning vector quantization* or self organizing map* or regularization algorithm* or iterative dichotomiser* or decision stump* or clustering algorithm* or k-means* or k-medians* or hierarchical clustering* or apriori* or auto encoder* or auto-encoder* or markov* or classification algorithm* or locally weighted learning* or support vector machine* or svm or decision tree* or component analy* or pca or ica or neural network* or hierarchical learning* or latent class analy* or latent class model* or latent variable model* or boltzmann or deep belief network* or lasso regression* or lda or adtree or ridge regression or enet or fnn or firefly algorithm* or multivariate adaptive regression splines* or locally estimated scatterplot smoothing* or instance base* or nearest neighbor* or regularization algorithm* or least absolute shrinkage select* or least angle regression* or eclat* or back propagation* or hopfield network* or radial basis function network* or dimensionality reduction algorithm* or ensemble algorithm* or adaboost or stacked generalization* or relu or naive bayes).mp.
6 4 or 5
7 3 and 6

IEEE Xplore (Institute of Electrical and Electronics Engineers) <1988 to Feb 2022>
(asperger syndrome OR autis* OR child development disorder) AND (artificial intelligence OR deep learning OR machine learning OR machine intelligence OR supervised learning OR unsupervised learning OR semi-supervised learning OR reinforcement learning OR regression algorithm OR learning vector quantization OR self organizing map OR regularization algorithm OR iterative dichotomiser OR decision stump OR clustering algorithm OR k-means OR kmedians OR hierarchical clustering OR apriori OR auto encoder OR auto-encoder OR markov OR classification algorithm OR locally weighted learning OR support vector machine OR svm OR decision tree* OR component analysis OR pca OR ica OR neural network* OR hierarchical learning OR latent class analysis OR latent class model OR latent variable model OR Boltzmann OR deep belief network OR lasso regression OR lda OR adtree OR ridge regression OR enet OR fnn OR firefly algorithm OR multivariate adaptive regression splines OR locally estimated scatterplot smoothing OR instance base OR nearest neighbors OR regularization algorithm OR least absolute shrinkage selection OR least angle regression OR eclat OR back propagation OR hopfield network OR radial basis function network OR dimensionality reduction algorithm OR ensemble algorithm OR adaboost OR stacked generalization OR relu OR naive bayes)

### Scopus (Elsevier) <1996 to Feb 2022>

TITLE-ABS-KEY(“asperger syndrome*” OR “autis*” OR “child development disorder*”) AND TITLE-ABS-KEY(“artificial intelligence*” OR “deep learning*” OR “machine learning*” OR “machine intelligence*” OR “supervised learning*” OR “unsupervised learning*” OR “semisupervised learning*” OR “reinforcement learning*” OR “regression algorithm*” OR “learning vector quantization*” OR “self organizing map*” OR “regularization algorithm*” OR “iterative dichotomiser*” OR “decision stump*” OR “clustering algorithm*” OR “k-means” OR “kmedians” OR “hierarchical clustering*” OR “apriori” OR “auto encoder*” OR “auto-encoder*” OR “markov” OR “classification algorithm*” OR “locally weighted learning*” OR “support vector machine*” OR “svm” OR ”decision tree*” OR “component analy*” OR “pca” OR “ica” OR “neural network*” OR “hierarchical learning*” OR “latent class analy*” OR “latent class model*” OR “latent variable model*” OR “boltzmann” OR “deep belief network*” OR “lasso regression*” OR “lda” OR “adtree*” OR “ridge regression*” OR “enet” OR “fnn” OR “firefly algorithm*” OR “multivariate adaptive regression splines*” OR “locally estimated scatterplot smoothing*” OR “instance base*” OR “instance-base*” OR “nearest neighbor*” OR “regularization algorithm*” OR “least absolute shrinkage select*” OR “least angle regression*” OR “eclat” OR “back propagation*” OR “hopfield network*” OR “radial basis function network*” OR “dimensionality reduction algorithm*” OR “ensemble algorithm*” OR “adaboost” OR “stacked generalization*” OR “relu” OR “naive bayes*”)

### Web of Science (Core Collection) <1900 to Feb 2022>

#1 TS=(“asperger syndrome*” OR “autis*” OR “child development disorder*”)
#2 TS=(“artificial intelligence*” OR “deep learning*” OR “machine learning*” OR “machine intelligence*” OR “supervised learning*” OR “unsupervised learning*” OR “semi-supervised learning*” OR “reinforcement learning*” OR “regression algorithm*” OR “learning vector quantization*” OR “self organizing map*” OR “regularization algorithm*” OR “iterative dichotomiser*” OR “decision stump*” OR “clustering algorithm*” OR “k-means” OR “kmedians” OR “hierarchical clustering*” OR “apriori” OR “auto encoder*” OR “auto-encoder*” OR “markov” OR “classification algorithm*” OR “locally weighted learning*” OR “support vector machine*” OR “svm” OR ”decision tree*” OR “component analy*” OR “pca” OR “ica” OR “neural network*” OR “hierarchical learning*” OR “latent class analy*” OR “latent class model*” OR “latent variable model*” OR “boltzmann” OR “deep belief network*” OR “lasso regression*” OR “lda” OR “adtree*” OR “ridge regression*” OR “enet” OR “fnn” OR “firefly algorithm*” OR “multivariate adaptive regression splines*” OR “locally estimated scatterplot smoothing*” OR “instance base*” OR “instance-base*” OR “nearest neighbor*” OR “regularization algorithm*” OR “least absolute shrinkage select*” OR “least angle regression*” OR “eclat” OR “back propagation*” OR “hopfield network*” OR “radial basis function network*” OR “dimensionality reduction algorithm*” OR “ensemble algorithm*” OR “adaboost” OR “stacked generalization*” OR “relu” OR “naive bayes*”)
#3 #1 AND #2

### OpenGrey <Feb 2022>

(“autis*” OR “asperger*” OR “child* development* disorder*”) AND (“artificial intelligence*” OR “deep learning*” OR “machine learning*” OR “machine intelligence*” OR “supervised learning*” OR “unsupervised learning*” OR “semi-supervised learning*” OR “reinforcement learning*” OR “regression algorithm*” OR “learning vector quantization*” OR “self organizing map*” OR “regularization algorithm*” OR “iterative dichotomiser*” OR “decision stump*” OR “clustering algorithm*” OR “k-means” OR “k-medians” OR “hierarchical clustering*” OR “apriori” OR “auto encoder*” OR “auto-encoder*” OR “markov” OR “classification algorithm*” OR “locally weighted learning*” OR “support vector machine*” OR “svm” OR ”decision tree*” OR “component analy*” OR “pca” OR “ica” OR “neural network*” OR “hierarchical learning*” OR “latent class analy*” OR “latent class model*” OR “latent variable model*” OR “boltzmann” OR “deep belief network*” OR “lasso regression*” OR “lda” OR “adtree*” OR “ridge regression*” OR “enet” OR “fnn” OR “firefly algorithm*” OR “multivariate adaptive regression splines*” OR “locally estimated scatterplot smoothing*” OR “instance base*” OR “instancebase*” OR “nearest neighbor*” OR “regularization algorithm*” OR “least absolute shrinkage select*” OR “least angle regression*” OR “eclat” OR “back propagation*” OR “Hopfield network*” OR “radial basis function network*” OR “dimensionality reduction algorithm*” OR “ensemble algorithm*” OR “adaboost” OR “stacked generalization*” OR “relu” OR “naïve bayes*”)

CRL (Center for Research Libraries Online Catalogue) <1949 to Feb 2022>

((autis*) or (asperger*) or (child development disorder*))

OATD (Open Access Theses and Dissertations) <Up to Feb 2022>

title:(((autis*) OR ("asperger syndrom*") OR ("child development disorder*")) AND ((“artificial intelligence*”) OR (“deep learning*”) OR (“machine learning*”) OR (“machine intelligence*”) OR (“supervised learning*”) OR (“unsupervised learning*”) OR (“semi-supervised learning*”) OR (“reinforcement learning*”) OR (“regression algorithm*”) OR (“learning vector quantization*”) OR (“self organizing map*”) OR (“regularization algorithm*”) OR (“iterative dichotomiser*”) OR (“decision stump*”) OR (“clustering algorithm*”) OR (“k-means”) OR (“kmedians”) OR (“hierarchical clustering*”) OR (“apriori”) OR (“auto encoder*”) OR (“autoencoder*”) OR (“markov”) OR (“classification algorithm*”) OR (“locally weighted learning*”) OR (“support vector machine*”) OR (“svm”) OR (”decision tree*”) OR (“component analy*”) OR (“pca”) OR (“ica”) OR (“neural network*”) OR (“hierarchical learning*”) OR (“latent class analy*”) OR (“latent class model*”) OR (“latent variable model*”) OR (“boltzmann”) OR (“deep belief network*”) OR (“lasso regression*”) OR (“lda”) OR (“adtree*”) OR (“ridge regression*”) OR (“enet”) OR (“fnn”) OR (“firefly algorithm*”) OR (“multivariate adaptive regression splines*”) OR (“locally estimated scatterplot smoothing*”) OR (“instance base*”) OR (“instancebase*”) OR (“nearest neighbor*”) OR (“regularization algorithm*”) OR (“least absolute shrinkage select*”) OR (“least angle regression*”) OR (“eclat”) OR (“back propagation*”) OR (“hopfield network*”) OR (“radial basis function network*”) OR (“dimensionality reduction algorithm*”) OR (“ensemble algorithm*”) OR (“adaboost”) OR (“stacked generalization*”) OR (“relu”) OR (“naive bayes*”)))
