## Supplementary material for "Automated diagnosis of autism: State of the art": Multimedia Appendix 2: Characteristics of included studies

#### rs-fMRI

| Parcellation[1] | Feature Selection[2] | ASD/TD | Validation | Accuracy | Sen/Spe | Age | Classifier[3] | AI Spec[4] | Risk of bias | Reference |
|---|---|---|---|---|---|---|---|---|---|---|
| ROI-based functional connectivity | | | | | | | | | | |
| AAL116 | LLFS | 63/72* | 70:30 | 97.5 | - | B | SVM | RBF kernel | ✓✓✓✓ | [50] |
| Self-generated | t test | 11/48*** | LOOCV | 96.6 | 81.8/100 | A | SVM | Linear kernel | ✓✓✓? | [51] |
| AAL116 | RFE-SSAE | 501/553* | 30-fold | 94.9 | 94/95.6 | B+C | LR | Simple LR | ✕✓✓✓ | [52] |
| RBS | AE | 155/148* | 10-fold | 91.1 | 89.9/92 | B | DRBM | - | ✕✓✓✓ | **[53]** |
| Power150 | LLFS | 126/126* | 66:33 | 90.8 | 89/93 | B+C | Ensemble | RF | ✓✓✓✓ | [54] |
| Clements35 | RFE | 255/276* | 10-fold | 90.6 | 90.6/90.6 | B+C | SVM | Linear kernel | ✕✓✓✓ | **[55]** |
| AAL90 | Threshold | 312/328* | 50-fold | 90.3 | 92.3/88.4 | B | DNN | PNN | ✕✓✓✓ | [56] |
| - | t test-MRMR-LLFS | 23/27* | 5-fold | 89.7 | 87.6/92 | B+C | Ensemble | - | ?✕✕✓ | [57] |
| AAL90 | SMF-t test | 505/530* | 90:10 | 88 | 91.5/86.5 | B+C | SVM | Linear kernel | ✕✓✕✓ | [58] |
| Multi atlas | SLR | 159/197* | 10-fold | 89.1 | 91/87.7 | B | Ensemble | - | ?✓✓✓ | [59] |
| AAL116 | None | 55/55* | 5-fold | 86.4 | - | B | AE + Softmax | Stacked SAE | ✕✓✕✓ | [60] |
| Multi atlas | L1SCCA | 74/107 | LOOCV | 85 | 80/89 | C | LR | Sparse LR | ✓✓✕✓ | [61] |
| HO110 | None | 186/184* | 5-fold | 84.9 | - | - | FCNN | - | ?✓✓✓ | [62] |
| Whole brain | AE | 505/530* | 10-fold | 84.1 | 80/75.3 | B+C | CNN | Standard | ✕✓✓✓ | **[63]** |
| Multi atlas | FFS | 505/530* | 10-fold | 83 | 83/84 | B+C | LR | Simple LR | ✕✓✓✓ | [64] |
| Multi atlas | SLR | 36/46* | 10-fold | 81.7 | 71.8/89.5 | B+C | SVM | Linear kernel | ?✕✕✓ | [65] |
| AAL116 | t test | 45/47* | LOOCV | 81.5 | 84.4/78.7 | B | SVM | Linear kernel | ✕✓✓✓ | [66] |
| AAL90 | Threshold | 443/435* | 90:10 | 81.1 | 80.4/81.7 | B+C | SVM | Gaussian kernel | ✕✓✓✓ | (16) |
| AAL116 | F score | 23/15* | 62:38 | 80.8 | - | - | SVM | Linear kernel | ?✓?✓ | **[68]** |
| DOS160 | Extra tree | 403/468* | 10-fold | 80.5 | 63.7/100 | B+C | SVM | Linear kernel | ✕✓✓✓ | [69] |
| AAL116 | LASSO | 539/573* | 85:15 | 79.6 | 83.6/75.6 | B+C | Ensemble | - | ✕✕✓✓ | [70] |
| AAL116 | LLFS-t test-RFE | 45/47* | LOOCV | 79.4 | 82.2/76.6 | B | SVM | Multi kernel | ✓✓✓✓ | [71] |
| CC400 | None | 505/530* | 80:20 | 79.2 | 69.6/85 | B+C | CNN | AlexNet | ✕✓✓✓ | [72] |
| Self-generated | F score | 112/128* | LOOCV | 79.2 | 77.8/80.5 | B | SVM | Linear kernel | ✕✓✓✓ | [73] |
| Power150 | Threshold | 403/468* | 10-fold | 79.2 | - | B+C | AE + Softmax | - | ✕✕✓✓ | [74] |
| DES162 | RFE | 59/59 | 10-fold | 79.1 | 73.3/85 | C | LR | Simple LR | ✓✓✕✓ | [75] |
| Whole brain | t test | 40/40 | LOOCV | 79 | 83/75 | B+C | LR | Simple LR | ✓✓✓✓ | [76] |
| AAL90 | SMF | 505/530* | LOOCV | 77.3 | - | B+C | SVM | Gaussian kernel | ✕✓✕✓ | [77] |
| DK70 | t test-RFE | 86/125* | 10-fold | 76.3 | 79.2/63.9 | B | SVM | Linear kernel | ?✕✓✓ | [78] |

| | | | | | | | | | | |
|---|---|---|---|---|---|---|---|---|---|---|
| RBS | None | 505/530* | 70:30 | 76 | 78/67 | B+C | AE + Softmax | Stacked SAE | ×✓✓✓ | [79] |
| CC400 | None | 505/530* | 10-fold | 76 | 70/81.7 | B+C | CNN | Self-proposed | ×✓✓✓ | [80] |
| CC400 | None | 505/530* | 5-fold | 75.3 | 74/76.6 | B+C | FCNN | - | ×✓✓✓ | [81] |
| SBC36 | LASSO | 15/45 | LOOCV | 75 | 76.7/73.3 | C | LR | Sparse LR | ✓✓✓✓ | [82] |
| Multi atlas | None | 419/530* | 10-fold | 74.5 | 80.7/66.7 | B+C | AE + Softmax | Stacked DAE | ×✓✓✓ | [83] |
| AAL116 | None | 505/530* | 90:10 | 74 | 94.9/69.9 | B+C | CNN | ResNet-18 + FCNN | ×✓✓✓ | [84] |
| Multi atlas | None | 300/300* | 5-fold | 72.5 | 71.7/73.3 | B | Ensemble | CRF + CNN | ×✓✓✓ | [85] |
| CC400 | None | 505/530* | 5-fold | 72 | 70.9/73 | B+C | LR | Ridge LR | ×✓✓✓ | [86] |
| CC200 | None | 505/530* | 10-fold | 71 | 73/66 | B+C | DNN | CapsNet | ×?✓✓ | [87] |
| HO110 | Threshold | 102/88* | LOOCV | 71 | 69/74 | B | SVM | Linear kernel | ×✓×✓ | [88] |
| CC200 | None | 505/530* | 10-fold | 71 | 71/71 | B+C | CNN | Self-proposed | ×✓✓✓ | [89] |
| Power150 | LEAN-LLFS | 387/436* | 5-fold | 70.9 | - | B+C | FCNN | - | ×✓✓✓ | [90] |
| CC200 | F score | 505/530* | 5-fold | 70.9 | 70.7/75.5 | B+C | AE + Softmax | - | ×✓✓✓ | [91] |
| CC200 | None | 505/530* | 10-fold | 70.8 | 62.2/79.1 | B+C | AE + Softmax | Stacked SAE | ×✓✓✓ | **[92]** |
| Power150 | None | 126/126* | 72:28 | 70.8 | - | B+C | Ensemble | CRF | ×✓✓✓ | [93] |
| CC200 | RFE | 442/556* | 80:20 | 70.7 | - | B+C | SVM | RBF kernel | ×✓✓✓ | [94] |
| DLA200 | SLR | 100/100* | 10-fold | 70.5 | 74/67 | B+C | CNN | Standard | ?×✓✓ | [95] |
| AAL90 | None | 38/23* | 5-fold | 70.4 | 72.5/67 | B+C | AE + Softmax | Stacked SAE | ✓✓×✓ | [96] |
| CC200 | None | 505/530* | 5-fold | 70.3 | 68.3/72.2 | B+C | AE + Softmax | - | ×✓✓✓ | [97] |
| CC400 | None | 505/530* | 10-fold | 70.2 | 77.5/61.8 | B+C | CNN | Standard | ×✓✓✓ | [98] |
| CC200 | RFE | 432/556* | 90:10 | 70.1 | - | B+C | SVM | Linear kernel | ×✓✓✓ | [99] |
| CC200 | None | 505/530* | 10-fold | 70 | 74/63 | B+C | AE + Softmax | Stacked DAE | ×✓✓✓ | [100] |
| Brodmann's areas | Elastic Nets | 167/205* | 10-fold | 70 | 65/75 | B | SVM | Linear kernel | ×✓✓✓ | [101] |
| AAL116 | None | 403/468* | 5-fold | 69.8 | 63.1/75.6 | B+C | FCNN | - | ×✓✓✓ | [102] |
| BASC64 | BOF | 75/100* | 5-fold | 69.7 | 54.6/80.9 | B+C | SVM | RBF kernel | ×✓✓✓ | [103] |
| AAL116 | Masking | 510/536* | 5-fold | 69.2 | 64.7/73.5 | B+C | AE + Softmax | Stacked SAE | ××?✓ | [104] |
| AAL116 | SLR | 250/218* | 5-fold | 69.1 | 70.2/66.4 | B+C | KNN | - | ×✓✓✓ | [105] |
| CC200 | Extra tree | 506/548* | 10-fold | 67.7 | 66.3/68.9 | B+C | SVM | Linear kernel | ×✓✓✓ | [106] |
| AAL116 | t test | 119/144* | 10-fold | 67.4 | 58.3/75 | B | DRBM | - | ?×✓✓ | [107] |
| HBM-Bzdok12 | IBP | 369/349* | 90:10 | 67.3 | - | B+C | SVM | Linear kernel | ✓✓✓✓ | [108] |
| DLA39 | MSDL | 403/468* | LOOCV | 67 | 61/72.3 | B+C | SVM | Linear kernel | ×✓×✓ | [109] |
| AAL116 | None | 1711/15903 | 70:15:15 | 67 | - | All | Ensemble | - | ××✓✓ | [110] |
| HO110 | EIIC | 539/573* | LOOCV | 67 | - | B+C | Ensemble | - | ×✓×✓ | **[111]** |
| AAL90 | EW | 527/569* | 5-fold | 66.9 | 66.4/70.4 | B+C | CNN | Self-proposed | ××✓✓ | [112] |
| Multi atlas | None | 200/200* | 80:20 | 65 | 65/65 | B+C | Ensemble | CRF | ×✓✓✓ | [113] |
| AAL116 | LLFS | 272/245* | LOOCV | 65 | - | - | SVM | Linear kernel | ?✓×✓ | **[114]** |

| | | | | | | | | | | |
|---|---|---|---|---|---|---|---|---|---|---|
| DOS160 | SDAE | 457/483* | 10-fold | 64 | 63.8/64.4 | B+C | SVM | RBF kernel | ✕✕✓✓ | [115] |
| CC200 | Threshold | 77/77* | 10-fold | 63 | 62/64 | C | SVM | Linear kernel | ✓✓✓✓ | [116] |
| Power150 | Graph based | 42/37* | LOOCV | 60.7 | - | B | SVM | Gaussian kernel | ?✕✕✓ | [117] |
| Whole brain | t test | 447/517* | LOOCV | 60 | 62/58 | B+C | LR | Simple LR | ✕✓✕✓ | [118] |
| **ROI-based functional connectivity + Clinical data** | | | | | | | | | | |
| Multi atlas | None | 408/401* | 10-fold | 73.2 | 74.5/71.7 | B+C | DNN | DANN | ✕✕✓✓ | [119] |
| HO110 | None | 200/200* | 80:20 | 65 | 65/65 | B | Ensemble | CRF | ✓✓✓✓ | [120] |
| **Graph metrics** | | | | | | | | | | |
| AAL116 | GERSMC | 103/106* | 70:30 | 96.8 | - | B | SVM | RBF kernel | ✓✓✓✓ | [121] |
| AAL116 | LLFS | 45/39* | 70:30 | 96.2 | - | B | SVM | RBF kernel | ✓✕✓✓ | [122] |
| AAL90 | LLFS | 50/42* | 80:20 | 95 | - | B | Ensemble | - | ✓✕✓✓ | [123] |
| AAL116 | SSFA | 28/19* | 10-fold | 95 | 97/91 | - | SVM | Gaussian kernel | ✕✓✕✓ | [124] |
| DOS160 | Graph based | 12/12 | LOOCV | 91 | - | C | SVM | Linear kernel | ✕✓✓✓ | [125] |
| 6 RSN | ICA-Graph based | 103/192* | LOOCV | 88.7 | 77.4/100 | B+C | SVM | RBF kernel | ✓✕✕✓ | [126] |
| AAL116 | Graph based | 45/47 | 10-fold | 87.7 | 87.8/87.7 | B | SVM | Linear kernel | ✕✕✕✓ | [127] |
| AAL116 | Prim algorithm | 59/46* | 5-fold | 86.7 | 87.5/85.7 | B+C | SVM | RBF kernel | ✓✕✕✓ | [128] |
| Power150 | LLFS | 73/88* | 5-fold | 81.2 | - | B | FCNN | - | ✕✓?✓ | [129] |
| HO110 | Graph based | 403/468* | 10-fold | 79.9 | - | B+C | Ensemble | - | ✕✓✓✓ | [130] |
| HO110 | None | 403/468* | 10-fold | 79.5 | 78.3/81.2 | B+C | GNN | GFT + FCNN | ✕✓✓✓ | [131] |
| Power150 | Threshold-RFE | 403/468* | 10-fold | 77.7 | - | B+C | LDA | - | ✕✓✓✓ | [132] |
| HO110 | LLFS | 416/451* | 80:20 | 76.3 | 81/71.3 | B+C | GNN | ID + FCNN | ✕✓✓✓ | [133] |
| Multi atlas | RFE | 419/530* | 10-fold | 75.9 | 79.2/71.5 | B+C | GNN | MT + Ensemble | ✕✓✓✓ | [134] |
| AAL116 | Threshold-RFE | 119/116* | LOOCV | 74.9 | 71.2/78 | B+C | LDA | - | ✕✓✕✓ | [135] |
| HO110 | None | 221/253* | LOOCV | 73.5 | 78.7/71.5 | B+C | GNN | LR-GCN + FCNN | ✕✓✕✓ | [136] |
| Whole brain | Graph based | 403/468* | 10-fold | 73.1 | 76/69 | B+C | Ensemble | - | ✕✓✓✓ | [137] |
| HO110 | RFE | 539/573* | 10-fold | 73 | 68.8/76.9 | B+C | GNN | GFT + FCNN | ✕✓✓✓ | [138] |
| HO110 | t test-LASSO | 403/468* | 10-fold | 72.4 | 71.2/75 | B+C | DNN | GAT | ✕✓✓✓ | [139] |
| HO110 | RFE-PCA-AE | 403/468* | 10-fold | 70.4 | - | B+C | GNN | GFT + FCNN | ✕✓✓✓ | [140] |
| Power150 | Graph based | 42/37 | 5-fold | 69.8 | - | B | Ensemble | RF | ✓✕✕✓ | [141] |
| HO110 | None | 403/468* | 10-fold | 69.5 | - | B+C | GNN | GFT + FCNN | ✕✓✓✓ | [142] |
| AAL90 | EW | 474/539* | 5-fold | 68.7 | 69.2/68.3 | B+C | CNN | Self-proposed | ✕✓✓✓ | [143] |
| Multi atlas | None | 485/544* | 60:10:30 | 67.3 | 70.4/64.2 | B+C | GNN | MT + KNN | ✕✓✓✓ | [144] |
| AAL116 | None | 402/464* | 10-fold | 67.2 | 65.9/68.4 | B+C | GNN | hi-GCN + RBF-SVM | ✕✓✓✓ | [145] |
| Whole brain | Graph based-LLFS | 42/37* | LOOCV | 67 | 70/63.9 | B | Ensemble | - | ?✓✕✓ | [146] |
| AAL116 | Graph based | 29/31* | External | 65.5 | - | C | SVM | Linear kernel | ✓✓?✓ | [147] |
| Power150 | Graph based | 42/37* | LOOCV | 63.3 | 73.8/51.4 | - | SVM | Grass kernel | ?✕✕✓ | [148] |

| | | | | | | | | | | |
|---|---|---|---|---|---|---|---|---|---|---|
| HO110 | None | 403/468* | 5-fold | 62.9 | - | B+C | GNN | s-GCN + FCNN | ✕✓✓✓ | [149] |
| CC200 | Graph based | 493/530* | 17-fold | 59.2 | 61.4/57.4 | B+C | FCNN | - | ✕✓✓✓ | [150] |
| **Graph metrics + Clinical data** | | | | | | | | | | |
| BNA246 | Graph based | 270/305* | 5-fold | 74.5 | 63.5/84.3 | B+C | CNN | Self-proposed | ✕✓✓✓ | [151] |
| Multi atlas | None | 539/573* | 90:10 | 70.2 | - | B+C | GNN | GFT + FCNN | ✕✓✓✓ | [152] |
| **Dynamic functional connectivity** | | | | | | | | | | |
| AAL116 | HMM | 47/73* | 10-fold | 90.1 | 85.5/93.2 | B | SVM | Linear kernel | ✕✓✕✓ | [153] |
| Multi-network | Self-proposed | 30/30* | LOOCV | 90 | 87/93 | B | SVM | Multi kernel | ?✓✓✓ | [154] |
| Whole brain | None | 42/42* | 28-fold | 89.3 | 88.1/83.3 | B | AE + Softmax | SAE | ✕✕✕✓ | [155] |
| TFP-IP | SSFA | 48/51* | 5-fold | 88 | 87/88 | B | KNN | - | ✕✓✕✓ | [156] |
| AAL116 | WT | 41/41* | 70:15:15 | 85.9 | 79.3/92.6 | - | KNN | - | ?✕?✓ | [157] |
| AAL116 | t test-LASSO | 45/47* | 6-fold | 83 | 82/84 | B | SVM | Linear kernel | ✓✓✕✓ | [158] |
| AAL116 | PCA | 49/41* | LOOCV | 78.9 | 85.7/70.7 | B+C | SVM | Linear kernel | ✓✓✓✓ | [159] |
| AAL116 | MTFS-EM | 403/468* | 10-fold | 76.8 | 72.5/79.9 | B+C | SVM | Multi kernel | ✕✓✓✓ | [160] |
| CC200 | RFE | 399/472* | 10-fold | 76.6 | 78.6/74.3 | B+C | SVM | Linear kernel | ✕✓✓✓ | [161] |
| AAL116 | HMM | 121/171* | 10-fold | 75.9 | 83.3/70.6 | B | SVM | Linear kernel | ✕✓✓✓ | [162] |
| CC200 | HMM | 145/157* | 10-fold | 74.9 | - | B+C | SVM | Linear kernel | ✓✓✓✓ | [163] |
| BASC64 | MSTEPS | 403/468* | 10-fold | 74.7 | 73/76.3 | B+C | LSTM | - | ✕✓✓✓ | [164] |
| CC200 | AE | 322/352* | LOOCV | 74.7 | 73/75 | B+C | LSTM | - | ✕✓✕✓ | [165] |
| AAL116 | LLFS | 423/446* | 10-fold | 73.6 | 75/72 | B+C | Ensemble | - | ✕✓?✓ | [166] |
| AAL116 | LLFS-LASSO | 45/47* | 10-fold | 71.4 | 60.5/81.6 | B | SVM | RBF kernel | ✓✓✕✓ | [167] |
| Whole brain | WT | 210/249* | 10-fold | 70.5 | 67.9/74.2 | B | CNN | Self-proposed | ✕✕✓✓ | [168] |
| HO110 | LLFS | 539/573* | 80:20 | 69.8 | 77.1/60.6 | B+C | SVM | Polynomial kernel | ✕✓✓✓ | [169] |
| CC200 | None | 539/573* | 10-fold | 68.5 | - | B+C | LSTM | - | ✕✓✓✓ | [170] |
| CC200 | Manual | 147/146* | 5-fold | 61.1 | 61.8/60 | B+C | SVM | Linear kernel | ?✓?✓ | [171] |
| AAL116 | BS | 25/25 | External | 53 | 50/55 | B | LSTM | - | ✕✓✓✓ | [172] |
| Multi atlas | ICA-PCA-LLFS | 32/34 | LOOCV | 50 | - | C | LDA | - | ✓✓✓✓ | [173] |
| **Effective connectivity** | | | | | | | | | | |
| AAL116 | t test | 48/30* | LOOCV | 87 | 80/92 | B+C | SVM | RBF kernel | ✕✓✓✓ | [174] |
| HO110 | DR-ICA-t test | 127/135* | LOOCV | 69 | 63.5/74.6 | B+C | LDA | - | ✕✓✕✓ | [175] |
| **3D fMRI** | | | | | | | | | | |
| Whole brain | None | 79/105* | - | 94.7 | 92.5/96.2 | B+C | CNN | DarkNet-19 | ✕✓?✓ | [176] |
| Whole brain | BOF | 19/19* | 70:30 | 81 | 79/83 | B | SVM | Linear kernel | ?✓✓✓ | [177] |
| **Power spectral density** | | | | | | | | | | |
| Whole brain | None | 117/81* | LOOCV | 96.2 | 98/93.6 | B | AE + Softmax | SAE | ✕✓✕✓ | [178] |

| | | | | | | | | | | |
|---|---|---|---|---|---|---|---|---|---|---|
| Whole brain | AE | 123/160*** | LOOCV | 92 | 93/89 | B | SVM | RBF kernel | ✓✕✕✓ | [179] |
| **Regional homogeneity** | | | | | | | | | | |
| AAL116 | Chi square | 443/435* | 75:25 | 68.9 | - | B+C | DNN | PBL-McRBFN | ✕✓✓✓ | [180] |
| Whole brain | Self-proposed | 539/573* | 5-fold | 62 | - | B+C | SVM | Linear kernel | ✕✕✓✓ | [181] |
| **High-order functional connectivity** | | | | | | | | | | |
| AAL116 | LASSO | 54/46* | 10-fold | 81 | 82/80 | B | SVM | Linear kernel | ✓✓✕✓ | [182] |
| CC200 | LLFS | 511/561* | 10-fold | 77.3 | 78/77.8 | B+C | DNN | PTN | ✕✓✓✓ | [183] |
| AAL116 | t test | 77/105 | 10-fold | 72.6 | 79/64 | B+C | Ensemble | Sparce-MVTC | ✕✓✓✓ | [184] |
| AAL116 | t test | 134/160* | 10-fold | 68.8 | 73.6/62.5 | B+C | Ensemble | Sparce-MVTC | ✕✕✓✓ | [185] |
| **Wavelet-based dynamics features** | | | | | | | | | | |
| HO110 | GARCH-t test | 222/246* | 5-fold | 75.3 | - | - | SVM | Linear kernel | ?✕✓✓ | [186] |
| 7 RSN | ICA-DR | 24/30 | LOOCV | 86.7 | 91.7/83.3 | B | SVM | Polynomial kernel | ?✓✓✓ | [187] |
| **Independent components** | | | | | | | | | | |
| Whole brain | ICA | 392/407* | - | 89.5 | 89.3/89.7 | B+C | Ensemble | - | ✕✓?✓ | [188] |
| Whole brain | ICA-DR | 20/20*** | LOOCV | 83 | 67/100 | B | LR | Simple LR | ✓✓✓✓ | [189] |
| 8 RSN | LLFS | 79/105* | 10-fold | 77.7 | 78.6/76.9 | B+C | CNN | Standard | ✕✓✓✓ | [190] |
| **Histogram of oriented gradients + Characteristics** | | | | | | | | | | |
| HO110 | MRMR | 538/573* | 80:20 | 65 | 71.3/58.3 | B+C | SVM | RBF kernel | ✕✓✓✓ | [191] |
| **ROI-based functional connectivity + Normalized image** | | | | | | | | | | |
| Multi atlas | None | 542/625* | 77:33 | 71.7 | - | B+C | CNN | 3D CNN | ✕✓✓✓ | [192] |
| **ROI-based functional connectivity + Clinical data + Information theory** | | | | | | | | | | |
| CC200 | RFE | 399/472* | 10-fold | 72.5 | 79.2/64.7 | B+C | SVM | Linear kernel | ✕✓✓✓ | [193] |
| **ROI-based functional connectivity + Amplitude of low-frequency fluctuation** | | | | | | | | | | |
| AAL90 | None | 99/85* | 10-fold | 68.5 | 69.5/67.6 | B+C | CNN | Standard | ✕✓✓✓ | [194] |
| **Topology** | | | | | | | | | | |
| CC200 | None | 505/530* | 5-fold | 69.2 | - | B+C | FCNN | - | ✕✓✓✓ | [195] |
| **Non-oscillatory connectivity** | | | | | | | | | | |
| AAL116 | t test | 36/36* | 10-fold | 80 | 80/80 | B+C | SVM | Polynomial kernel | ✕✕✕✓ | [196] |
| **Combined features** | | | | | | | | | | |
| 9 measures | None | 620/542* | 5-fold | 64 | - | B+C | SVM | Linear kernel | ✕✓✓✓ | [197] |
| **Normalized image** | | | | | | | | | | |
| EPI images | None | 69/69* | 70:15:15 | 98.4 | - | B | CNN | Inception V3 | ✓✕✓✓ | [198] |
| Glass brain-Stat_map | LLFS | 529/573* | 85:15 | 82.7 | - | B+C | Ensemble | - | ✕✓✓✓ | [199] |

| | | | | | | | | | | |
|---|---|---|---|---|---|---|---|---|---|---|
| EPI images | None | 74/98* | 70:15:15 | 57.8 | 57.2/61.3 | B+C | CNN | InceptionResNet V2 | ✗✓✓✓ | [200] |

#### DWI/DTI

| Feature Selection[2] | ASD/TD | Validation | Accuracy | Sen/Spe | Age | Classifier[3] | AI Spec[4] | Risk of bias | Reference |
|---|---|---|---|---|---|---|---|---|---|
| **Fractional anisotropy** | | | | | | | | | |
| PCA | 41/32 | LOOCV | 75.3 | 71.9/71.9 | - | SVM | Linear kernel | ?✓✗? | [201] |
| **Fiber density + Fiber bundle cross-section** | | | | | | | | | |
| t test-LASSO-RFE | 26/26*** | 10-fold | 73.1 | 71.1/75.1 | C | SVM | Linear kernel | ✓✓✗✓ | **[202]** |
| **Fractional anisotropy + Mean diffusivity** | | | | | | | | | |
| S2n | 70/80 | LOOCV | 81.3 | - | B | SVM | Polynomial kernel | ✓✓✗? | [203] |
| S2n | 45/30 | LOOCV | 80 | 74/84 | B | SVM | RBF kernel | ✗✓✗✓ | [204] |
| S2n | 70/79 | 10-fold | 78.3 | 84.8/72.9 | B | SVM | Polynomial kernel | ✓✓✓✓ | [205] |
| **Fractional anisotropy + Axial diffusivity + Radial diffusivity + Spherical harmonics** | | | | | | | | | |
| t test | 19/19 | LOOCV | 86.8 | - | A | Ensemble | RF | ✓✓✓? | [206] |
| **Fractional anisotropy + Mean diffusivity + Axial diffusivity + Radial diffusivity + Skewness** | | | | | | | | | |
| RFE | 125/100* | 5-fold | 99 | - | B+C | SVM | Linear kernel | ✗✓✓✓ | [207] |
| None | 30/30 | 70:30 | 94.7 | 91.7/100 | B+C | QDA | - | ✓✓✓✓ | [208] |
| S2n | 124/139*** | LOOCV | 73 | 70/76 | B | SVM | Linear kernel | ✗✓✗✓ | [209] |
| **Graph metrics** | | | | | | | | | |
| Graph based-LLFS | 43/51* | LOOCV | 68 | 70/65.3 | B | Ensemble | - | ?✓✗✓ | **[146]** |
| Graph based | 42/37* | LOOCV | 68 | - | B | SVM | Gaussian kernel | ?✗✗✓ | [117] |

#### EEG

| Feature Selection[2] | Channels | ASD/TD | Validation | Accuracy | Sen/Spe | Age | Classifier[3] | AI Spec[4] | Risk of Bias | Reference |
|---|---|---|---|---|---|---|---|---|---|---|
| **Entropy + Spatiotemporal features** | | | | | | | | | | |
| MI | 128 | 48/48 | 10-fold | 95.7 | - | B | SVM | Gaussian kernel | ✓✓✓? | [210] |
| LLFS | 18 | 15/10 | LOOCV | 92.8 | - | B | Ensemble | RF | ✗✓✓✓ | [211] |
| **Frequency-domain + Spatiotemporal features** | | | | | | | | | | |
| Manual | 19 | 9/9 | 78:22 | 95.5 | - | B | PNN | - | ?✓✓✓ | [212] |
| Manual | 19 | 10/9 | - | 89.5 | - | B | KNN | - | ✗✓?✓ | [213] |
| **Complex networks** | | | | | | | | | | |
| Graph based | 128 | 12/12 | LOOCV | 94.7 | 85.7/100 | B | SVM | Polynomial kernel | ?✓✓✗ | [214] |
| MI | 21 | 28/28 | 10-fold | 92.3 | - | B | SVM | Gaussian kernel | ✓✓✗✓ | **[215]** |
| PCA | 24 | 430/554 | 50:50 | 86 | - | B | DFA | - | ✓✓✓✓ | **[216]** |

| | | | | | | | | | | |
|---|---|---|---|---|---|---|---|---|---|---|
| Graph based | 10 | 30/30 | LOOCV | 81.7 | 83.3/80 | B | KNN | - | ✕?✕✕ | [217] |
| **Entropy + Frequency-domain + Time-domain + Non-linear features** | | | | | | | | | | |
| None | - | 48/50 | 50:50 | 97.9 | 96/95.8 | B | PSN-ANFIS | - | ??✓✕ | [218] |
| MI | 10 | 34/11 | 70:30 | 94.7 | 99.1/- | B | SVM | Linear kernel | ✕✕✓✓ | [219] |
| **Entropy + Non-linear features** | | | | | | | | | | |
| RFE | 19 | 18/23 | 10-fold | 97 | 100/94 | B | SVM | Linear kernel | ✕✓✕✓ | [220] |
| PCA | 17 | 7/7 | LOOCV | 92.9 | 100/85.7 | B | SVM | RBF kernel | ✕✓✓✓ | [221] |
| None | 19 | 20/20 | 75:25 | 91.9 | - | B | RNN | - | ?✓✓✓ | **[222]** |
| RFE | 19 | 21/102 | LOOCV | 90.2 | 95/89 | A | SVM | RBF kernel | ✓✓✕✓ | [223] |
| **Entropy + Frequency-domain + Non-linear features** | | | | | | | | | | |
| None | 64 | 9/10 | 10-fold | 84.5 | - | B | KNN | - | ✕✓✕? | [224] |
| **Time-domain features** | | | | | | | | | | |
| Manual | 40 | 10/10 | 3-fold | 98.6 | - | C | SVM | Linear kernel | ✓✓✕✓ | [225] |
| **Event-related potentials** | | | | | | | | | | |
| ICA | 19 | 41/32 | - | 90.4 | 90.4/89.8 | B | SVM | Linear kernel | ✓✓?? | [226] |
| Manual | 128 | 19/30 | LOOCV | 79 | 68/87 | B | NB | - | ✕✓✓✓ | [227] |
| GA | 128 | 19/112 | 70:30 | 70 | 40/100 | A | SVM | Linear kernel | ✕✓✓✓ | [228] |
| **Entropy + Frequency-domain features** | | | | | | | | | | |
| Manual | 14 | 89/94 | 10-fold | 99.7 | 99.9/95.9 | B | SVM | Polynomial kernel | ✓✓✓✓ | [229] |
| None | 16 | 8/4 | 10-fold | 99.7 | - | B | FCNN | - | ✕✓✕? | [230] |
| t test | 64 | 40/37 | 10-fold | 98.7 | 100/97.3 | B | PNN | - | ✓✓✓✓ | [231] |
| LLFS | 19 | 10/18 | LOOCV | 96 | 90/100 | - | SVM | RBF kernel | ?✓✓✕ | [232] |
| Manual | 23 | 9/10 | 10-fold | 91.2 | - | B | FCNN | - | ✕✓✕? | [233] |
| - | 128 | 25/25 | - | 87 | - | B | SVM | Linear kernel | ?✓?✓ | [234] |
| Manual | 16 | 100/100 | 10-fold | 86 | 85.9/89.7 | B | SVM | Polynomial kernel | ✓✓✓✓ | [235] |
| **Frequency-domain features** | | | | | | | | | | |
| None | 32 | 8/9 | 10-fold | 98.3 | 98/98 | B | Ensemble | RF | ✕✓✕✓ | [236] |
| LLFS | 64 | 61/61 | 10-fold | 96.4 | 97.8/93.2 | B | SVM | Polynomial kernel | ✓✓✓✓ | [237] |
| Manual | 19 | 15/11 | - | 96.2 | - | B | KNN | - | ✓??✓ | [238] |
| PCA | 16 | 12/4 | 10-fold | 95.2 | 97.1/90.9 | B+C | SVM | Linear kernel | ✕✓✕? | [239] |
| PCA | 64 | 60/60 | - | 94.4 | 100/88.9 | B | FCNN | - | ?✓?? | [240] |
| Manual | 32 | 10/5 | k-fold | 93.3 | - | B | Ensemble | RF | ?✕✕✓ | [241] |
| LLFS | 16 | 8/4 | 10-fold | 90 | - | B | FLDA | - | ✕✓✕? | [242] |
| PCA | 8 | 6/6 | - | 90 | - | B | FCNN | - | ?✓?✕ | **[243]** |

| | | | | | | | | | | |
|---|---|---|---|---|---|---|---|---|---|---|
| LLFS | - | 8/10 | 18-fold | 88.9 | 88.9 | B | FLDA | - | ??✗✗ | [244] |
| Manual | 8 | 6/6 | 5-fold | 86.6 | - | - | FCNN | - | ??✗✗ | [245] |
| Manual | 128 | 36/69 | 5-fold | 85.2 | - | B+C | PTN | - | ?✗✓✓ | [246] |
| None | 22 | 10/10 | 5-fold | 81.9 | 91/100 | - | CNN | ResNet-50 | ?✓✗✗ | [247] |
| Manual | 19 | 11/10 | - | 81 | - | B | KNN | - | ✗✓?✓ | [248] |
| None | 16 | 13/4 | 70:15:15 | 80.9 | - | - | FCNN | - | ?✓✓✗ | [249] |
| **Frequency-domain + Time-domain features** | | | | | | | | | | |
| None | 8 | 46/63 | 10-fold | 92.7 | - | B | SVM | Linear kernel | ✓✓✓✓ | [250] |
| None | 16 | 8/18 | 10-fold | 80 | - | B | CNN | Standard | ?✓✗✗ | [251] |
| **Non-linear features** | | | | | | | | | | |
| - | 32 | 8/9 | 5-fold | 85.5 | - | B | FCNN | - | ?✗✗✗ | [252] |
| MI | 2 | 34/27 | 5-fold | 82 | 94.1/66.7 | B | SVM | RBF kernel | ?✗✓✗ | [253] |
| **Raw data** | | | | | | | | | | |
| ICA | 16 | 9/10 | 10-fold | 99.5 | - | B | Ensemble | RNN-GRU + CNN | ?✓✗? | [254] |
| None | 16 | 8/12 | 80:10:10 | 80 | - | B | CNN | Standard | ?✓✓? | [255] |
| **Frequency-domain + Non-linear features** | | | | | | | | | | |
| t test | 19 | 9/8 | 80:20 | 90 | - | B | FCNN | RBF | ✓✓✓✗ | [256] |
| t test | 19 | 17/11 | - | 78.5 | 80/71 | B | KNN | - | ✗✓?✓ | [257] |

#### Eye Tracking

| Feature Selection[2] | ASD/TD | Validation | Accuracy | Sen/Spe | Age | Classifier[3] | AI Spec[4] | Risk of bias | Reference |
|---|---|---|---|---|---|---|---|---|---|
| **Interactions with parents** | | | | | | | | | |
| Manual | 6/26 | LOOCV | 93.8 | 100/92.3 | A | VMM | - | ?-✓? | [258] |
| **Watching videos** | | | | | | | | | |
| LLFS | 61/72 | 10-fold | 87.5 | 87.5/87.5 | B | SVM | Linear kernel | ✗-✓✓ | [259] |
| Manual | 17/15 | 75:25 | 83.4 | 100/66.6 | B | LSTM | - | ?-✓? | [260] |
| PCA | 29/30 | 10-fold | AUC 92 | - | A | FCNN | - | ?-✗✓ | [261] |
| GA | 76/30 | 5-fold | AUC 82 | 69/93 | B | FCNN | - | ?-✓✓ | [262] |
| **Watching websites** | | | | | | | | | |
| t test-RFE | 15/15 | 80:20 | 91.6 | 92.9/90 | C | DT | - | ✓-✓✓ | [263] |
| Manual | 15/15 | 70:30 | 70 | - | C | LR | Simple LR | ✓-✓✓ | [264] |
| Manual | 19/19 | 70:30 | 65 | - | C | LR | Simple LR | ✗-✓✓ | [265] |
| None | 15/15 | 66:33 | 60 | - | C | STA | - | ✓-✓✓ | [266] |
| **Observing images** | | | | | | | | | |

| | | | | | | | | | |
|---|---|---|---|---|---|---|---|---|---|
| F score | 20/19 | LOOCV | 92 | 93/92 | C | Ensemble | CNN + SVM | ✓-✓✓ | [267] |
| LLFS | 14/14 | 75:25 | 59.3 | 68.4/50.6 | B | DT | - | ✓-✓✓ | [268] |

#### Saliency maps

| | | | | | | | | | |
|---|---|---|---|---|---|---|---|---|---|
| None | 14/14 | LOOCV | 99.8 | 100/99.7 | - | Ensemble | XGBoost | ?-✓? | [269] |
| None | 20/19 | LOOCV | 99 | 100/98 | - | Ensemble | CNN + LSTM | ?-✓? | [270] |
| Manual | 37/37 | 5-fold | 85.1 | 86.5/83.8 | B | SVM | Linear kernel | ✓-✓✓ | [271] |
| None | 14/14 | 60:15:25 | 62.1 | 71/54 | B | CNN | ResNet-50 | ?-✓? | [272] |

#### Virtual reality

| | | | | | | | | | |
|---|---|---|---|---|---|---|---|---|---|
| RFE | 35/20 | 5-fold | 86 | 91/82 | B | SVM | Linear kernel | ?-✗✓ | [273] |
| LLFS | 55/52 | 3-fold | 73 | 81/65.8 | C | Ensemble | - | ✓-✓? | [274] |

#### Watching faces

| | | | | | | | | | |
|---|---|---|---|---|---|---|---|---|---|
| LLFS | 29/48 | LOOCV | 88.5 | 93.1/86.2 | B | SVM | RBF kernel | ?-✗✗ | [275] |
| LLFS | 20/41 | LOOCV | 86.9 | - | B | SVM | RBF kernel | ?-✗✗ | [276] |
| MRMR | 77/80 | 10-fold | 84.2 | - | B | SVM | Linear kernel | ✓-✓✓ | [277] |

#### Face-to-face conversations

| | | | | | | | | | |
|---|---|---|---|---|---|---|---|---|---|
| FFS | 20/19 | LOOCV | 92.3 | 84.2/100 | B | SVM | Linear kernel | ✓-✓✓ | [278] |

### Facial Recognition

#### Static facial features

| Feature Selection[2] | ASD/TD* | Validation | Accuracy | Sen/Spe | Age | Classifier[3] | AI Spec[4] | Risk of bias | Reference |
|---|---|---|---|---|---|---|---|---|---|
| None | 556/556 | 80:20 | 95 | 95/95 | B | CNN | VGG-16 + TL | ✗✗✓? | [279] |
| None | 1507/1507** | 90:10 | 94.6 | - | B | CNN | MobileNet-V1 | ✗✗✓✗ | [280] |
| None | 1269/1269** | 92:8 | 91.5 | 93.4/89.7 | B | CNN | DenseNet | ✗✗✓✗ | [281] |
| None | 1468/1468** | 80:20 | 91 | - | B | CNN | VGG-16 + TL | ✗✗✓✗ | [282] |
| None | 1468/1468** | 70:30 | 91 | - | B | CNN | VGG-16 + TL | ✗✗✓✗ | [283] |
| None | 1468/1468** | 87:3:10 | 90.7 | 90.7/90.7 | B | CNN | MobileNet-V1 + TL | ✗✗✓✗ | [284] |
| None | 1468/1468** | 86:4:10 | 90 | 88.5/91.7 | B | CNN | Xception + TL | ✗✗✓✗ | [285] |
| None | 1426/1374** | 86:7:7 | 87 | - | B | CNN | MobileNet-V1 | ✗✓✓✗ | [286] |
| None | 1568/1568** | 81:16:3 | 87 | 87/87 | B | CNN | MobileNet-V1 + TL | ✗✗✓✗ | [287] |

#### Facial attribute recognition

| | | | | | | | | | |
|---|---|---|---|---|---|---|---|---|---|
| Manual | 20/26 | LOOCV | 80.8 | - | B | CNN | - | ✗✓✓✓ | [288] |
| Manual | 49/39 | LOOCV | 72.9 | 76/69 | - | CNN | BottleNeck + TL | ✗✗✓✗ | [289] |

### S-MRI

| Feature Selection[2] | ASD/TD | Validation | Accuracy | Sen/Spe | Age | Classifier[3] | AI Spec[4] | Risk of bias | Reference |
|---|---|---|---|---|---|---|---|---|---|

#### Volume-based morphological features

| | | | | | | | | | |
|---|---|---|---|---|---|---|---|---|---|
| Manual | 24/24* | LOOCV | 98 | - | B+C | Ensemble | RF | ??✓✓ | [290] |
| None | 74/208 | 10-fold | 92 | 87/93 | A | Ensemble | CNN + SFCNN | ✓✓✓✓ | [291] |
| VBM | 24/24 | 10-fold | 92 | - | B | SVM | RBF kernel | ✓✓✗✓ | [292] |
| F score | 78/104* | 10-fold | 90.4 | 84.4/95.9 | B+C | AE + Softmax | Stacked SAE | ✗✓✓✓ | [293] |
| VBM | 60/60 | LOOCV | 86.7 | 86.4/86.8 | C | SVM | Linear kernel | ✓✓✗✓ | [294] |
| VBM | 36/36 | LOOCV | 81 | 81/81 | C | LDA | PLS | ✓✓✗✓ | [295] |
| Manual | 52/40 | 10-fold | 77.2 | - | B | SVM | Linear kernel | ✗?✓✓ | [296] |
| RFE | 22/22 | LOOCV | 77 | 77/77 | C | SVM | Linear kernel | ✓✓✓✓ | [297] |
| LASSO | 54/57* | 10-fold | 75.4 | 74.6/76 | B | SVM | Linear kernel | ✗✓✓✓ | [298] |
| Manual | 120/136* | 80:20 | 73 | 92/68 | C | SVM | Linear kernel | ?✓✓✓ | [299] |
| VBM | 79/105* | 75:25 | 70 | 53/72 | B+C | DNN | PBL-McRBFN | ?✓✓✓ | [300] |
| None | 1555/12623 | 66:17:17 | 66.4 | - | All | CNN | Ensemble | ✗✓✓✓ | [301] |
| None | 1060/1166* | 5-fold | 63.8 | - | B+C | CNN | Fed-3D-Resnet-18 | ✗?✓✓ | [302] |
| VBM | 449/451* | 75:25 | 59.6 | - | B+C | DNN | PBL-McRBFN | ✗✓✓✓ | [303] |
| VBM-RFE | 38/38 | LOOCV | AUC 80 | - | B | SVM | Linear kernel | ✓✓✓✓ | [304] |

#### Surface-based morphological features

| | | | | | | | | | |
|---|---|---|---|---|---|---|---|---|---|
| LASSO-DL | 364/361* | 10-fold | 83 | 80/85 | B+C | Ensemble | - | ✓✓✓✓ | [305] |
| RFE | 100/100* | 60:40 | 80 | 72.5/67.5 | B+C | Ensemble | - | ?✓✓✓ | [306] |
| LLFS-PCA | 50/150* | 5-fold | 75 | 98.6/40 | C | SVM | Linear kernel | ✓✓✓✓ | [307] |

#### Cortical thickness

| | | | | | | | | | |
|---|---|---|---|---|---|---|---|---|---|
| RFE | 20/20 | LOOCV | 90 | 90/90 | C | SVM | Linear kernel | ✗✓✓✓ | [308] |
| None | 22/16 | 10-fold | 87 | 95/75 | B | LMT | - | ✓✓✗✓ | [309] |
| RFE | 40/36 | LOOCV | 84.2 | 80/88.9 | B | SVM | RBF kernel | ✗?✗✓ | [310] |
| MRMR | 325/325* | 5-fold | 62 | - | B+C | FCNN | - | ✗✓✓✓ | [311] |

#### Normalized image

| | | | | | | | | | |
|---|---|---|---|---|---|---|---|---|---|
| None | 112/102* | 5-fold | 99 | 99/99 | C | Ensemble | CNN + SFCNN | ✓✓✓✓ | [312] |
| None | 521/573* | - | 87 | - | - | CNN | ResNet-50 | ✗✗?✓ | [313] |
| DSM | 171/176 | 65:22:13 | 84.4 | 85/84 | A | CNN | CSResNet-18 | ✓✓✓✓ | [314] |
| GA | 500/500* | 5-fold | 73 | - | B | CNN | 3D CNN | ✗✓✓✓ | [315] |
| None | 500* | - | 71 | 66/69 | B+C | CNN | 3D-ResNet | ?✗?✓ | [316] |
| None | 946/1046* | 80:20 | 64 | - | B+C | CNN | 3D CNN | ✗✓✓✓ | [317] |

#### Volume-based morphological features + Cortical thickness

| | | | | | | | | | |
|---|---|---|---|---|---|---|---|---|---|
| t test-MRMR-RFE | 58/59*** | 2-fold | 96.3 | 95/97 | B | SVM | Multi kernel | ✓?✓✓ | [318] |
| None | 245/245* | 90:10 | 60 | - | B+C | LDA | - | ✓✓✓✓ | [319] |

| | | | | | | | | | |
|---|---|---|---|---|---|---|---|---|---|
| None | 83/105 | 70:30 | AUC 64 | - | C | AE + Softmax | Denoising AE | ✓✓✓✓ | [320] |
| Surface-based morphological features + Cortical thickness | | | | | | | | | |
| Elastic net-LASSO | 100/100* | 66:33 | 96.3 | 95.5/97 | B | RR | - | ✓✓✓✓ | [321] |
| None | 361/373* | LOOCV | 60 | 57/64 | B+C | Ensemble | RF | ✗?✗✓ | [322] |
| Surface-based morphological features + Clinical data | | | | | | | | | |
| Manual | 26/24* | 70:30 | 88 | 80/92.9 | - | Ensemble | RF | ?✓✓✓ | **[323]** |
| Surface-based morphological features + Volume-based morphological features | | | | | | | | | |
| RFE | 209/530* | 85:15 | 80 | - | B+C | FCNN | - | ✗✓✓✓ | [324] |
| Manual | 539/573* | LOOCV | 56.3 | - | B+C | FCNN | - | ✗✓✗✓ | **[325]** |
| Surface-based morphological features + Volume-based morphological features + Cortical thickness | | | | | | | | | |
| LLFS | 34/145*** | 10-fold | 93.8 | 88/95 | A | SVM | Linear kernel | ✓✓✓✓ | [326] |
| RFE | 530/573* | 5-fold | 71.6 | - | B+C | FCNN | - | ✗✓✓✓ | [327] |
| None | 325/325* | 5-fold | 59 | - | B+C | SVM | Linear kernel | ✗✓✓✓ | **[328]** |
| Manual | 1060/1166* | 75:25 | AUC 79 | - | B+C | LR | Simple LR | ✗✓✓✓ | [329] |
| VBM | 21/20 | LOOCV | - | 81/65 | B | SVM | Linear kernel | ✓✓✓✓ | [330] |
| Volume-based morphological features + Clinical data | | | | | | | | | |
| Manual | 220/303* | 4-fold | 64 | 63/64 | All | SVM | Not mentioned | ✗??✓ | [331] |
| Histogram of oriented gradients | | | | | | | | | |
| Threshold | 55/209*** | 10-fold | 76.2 | - | B | CNN | Multi-channel | ✓✗✓✓ | [332] |
| LLFS | 119/131* | 10-fold | 65 | 73/58 | B+C | SVM | Linear kernel | ✗?✓✓ | [333] |
| Path signature + Clinical data | | | | | | | | | |
| None | 30/30 | 10-fold | 87 | 83/90 | A | FCNN | SFCNN | ✓✓✓✓ | [334] |
| Curvelet sub-bands | | | | | | | | | |
| WT | 76/75* | 10-fold | AUC 75 | 77/82 | B | SVM | RBF kernel | ✓✓✓✓ | [335] |
| Voxel density | | | | | | | | | |
| t test | 81/50 | LOOCV | 73.3 | 71.6/76 | B | Ensemble | - | ✗✓✗? | [336] |
| Volume-based morphological features + Voxel density | | | | | | | | | |
| None | 30/28* | 10-fold | 82 | 82.4/81.7 | B+C | CNN | 3D Grad-CAM | ?✓✗✓ | [337] |
| Histogram of oriented gradients + Clinical data | | | | | | | | | |
| MRMR | 538/573* | 80:20 | 64.1 | 75.7/51.9 | B+C | SVM | RBF kernel | ✗✓✓✓ | [191] |
| Structural covariance network | | | | | | | | | |
| None | 518/567* | 10-fold | 71.8 | 81.3/68.8 | B+C | CNN | ResNet | ✗✓✓✓ | [338] |
| Morphological brain networks | | | | | | | | | |

| | | | | | | | | | |
|---|---|---|---|---|---|---|---|---|---|
| LLFS | 155/186* | 10-fold | 76.7 | - | B+C | GNN | Hypergraph | ✗✓✓✓ | [339] |
| None | 155/186* | 10-fold | 62.4 | - | B+C | Ensemble | - | ✗✓✓✓ | [340] |
| Graph based | 155/186* | 10-fold | 52 | - | B+C | SVM | Linear kernel | ??✓✓ | [341] |
| **T-fMRI** | | | | | | | | | |
| **Feature Selection[2]** | **ASD/TD** | **Validation** | **Accuracy** | **Sen/Spe** | **Age** | **Classifier[3]** | **AI Spec[4]** | **Risk of bias** | **Reference** |
| Dynamic bodies attention task | | | | | | | | | |
| RFE | 15/14 | LOOCV | 92.3 | 92.3/92.3 | C | SVM | Linear kernel | ✓✓✓✓ | [342] |
| Social and non-social attention task | | | | | | | | | |
| t test | 23/22 | LOOCV | 90 | - | B | SVM | Linear kernel | ✓✓✓✓ | [343] |
| Thinking about social interactions | | | | | | | | | |
| Manual | 17/17*** | LOOCV | 97 | - | C | NB | Gaussian | ✓✓✓✓ | [344] |
| Biopoint task | | | | | | | | | |
| Manual | 82/48 | 85:7:8 | 87.1 | - | - | CNN | 2C 3D CNN | ?✓✓? | [345] |
| None | 72/43 | 60:20:20 | 79.8 | 72.6/85.4 | - | GNN | BrainGNN | ?✓✓✓ | [346] |
| Graph based | 75/43 | 80:20 | 76 | 82/68.8 | - | GNN | - | ?✓✓? | [347] |
| PCA | 82/48 | 85:7:8 | F 89 | - | - | CNN | 2C 3D CNN | ?✓✓? | [348] |
| Biological motion perception task | | | | | | | | | |
| Bootstrap | 21/19 | 10-fold | 64.5 | 70.7/60.9 | B | LSTM | - | ?✓✗? | [349] |
| Language task + Theory of mind task | | | | | | | | | |
| Manual-t test | 13/14 | LOOCV | 96.3 | - | C | LR | Simple LR | ✓✓✓✓ | [350] |
| Cognitive control task | | | | | | | | | |
| Manual | 29/29 | LOOCV | 82.8 | 82.8/82.8 | - | LR | Simple LR | ??✓✗ | [351] |
| Language task | | | | | | | | | |
| LLFS | 30/30*** | - | 75.8 | 74.8/76.7 | B | SNCAE | - | ?✓?✓ | [352] |
| Response to speech | | | | | | | | | |
| LLFS-WT | 50/50*** | 4-fold | 86 | 82/92 | B | CNN | 2D CNN | ✓✓✓✓ | [353] |
| LLFS-WT | 50/50*** | 10-fold | 80 | 84/76 | B | CNN | 2D CNN | ✓✓✓✓ | [354] |
| LLFS-WT-Manual | 33/33*** | 4-fold | 77.2 | 78.1/76.5 | B | CNN | 1D CNN | ✓✓✓✓ | [355] |
| **Other Modalities** | | | | | | | | | |
| **Feature Selection[2]** | **ASD/TD** | **Validation** | **Accuracy** | **Sen/Spe** | **Age** | **Classifier[3]** | **AI Spec[4]** | **Risk of bias** | **Reference** |
| Functional near-infrared spectroscopy | | | | | | | | | |
| None | 22/22 | 70:30 | 95.7 | 97.1/94.3 | B | Ensemble | LSTM + CNN | ✗✓✓✓ | [356] |

| | | | | | | | | | |
|---|---|---|---|---|---|---|---|---|---|
| None | 25/22 | 60:20:20 | 92.2 | 85/99.4 | B | Ensemble | GRU + CNN | ✗✓✓✓ | [357] |
| Manual | 25/22 | 50:50 | 91.9 | 81.6/94.6 | B | SVM | Linear kernel | ✗✓✓✓ | [358] |
| **Kinematic features** | | | | | | | | | |
| Manual | 15/15 | LOOCV | 96.7 | 100/93.8 | B | SVM | Linear kernel | ✓-✓✓ | [359] |
| t test-FFS | 18/20 | LOOCV | 92.1 | 88.9/95 | B | DT | - | ?-✓✓ | [360] |
| t test-FFS | 20/23 | LOOCV | 88.4 | 85/91.3 | B | KNN | - | ✓-✓✓ | [361] |
| Manual | 16/16 | 10-fold | 86.7 | 85.7/87.5 | C | SVM | Linear kernel | ✓-✗✓ | [362] |
| FDR | 37/45 | 10-fold | AUC 93 | 83/85 | B | Ensemble | RGF | ✓-✓✓ | [363] |
| **Response to name** | | | | | | | | | |
| PCA | 22/21 | 75:25 | 93 | 100/92.3 | B | DT | - | ✓-✓✓ | [364] |
| **Video analysis** | | | | | | | | | |
| Manual | 149/79 | 70:30 | 88.9 | 94.5/77.4 | B | LR | Simple LR | ✗-✓? | [365] |
| LLFS | 9/124 | 75:8:17 | 82 | 92/71 | A | FCNN | - | ?-✓✓ | [366] |
| **Taking photos** | | | | | | | | | |
| None | 16/21 | 60:20:20 | 83.7 | - | C | CNN | VGG-16 + TL | ✓-✓✓ | [367] |
| **Vocal analysis** | | | | | | | | | |
| Manual | 1355/1257 | 45:55 | 86.5 | 84/89.2 | B | Ensemble | RF | ?-✓✓ | [368] |
| **PET scan** | | | | | | | | | |
| RFE | 45/13 | LOOCV | 88 | 91/77 | B | LDA | - | ✓✓✓✓ | [369] |

#### Multimodal

| Feature Selection² | ASD/TD | Validation | Accuracy | Sen/Spe | Age | Classifier³ | AI Spec⁴ | Risk of bias | Reference |
|---|---|---|---|---|---|---|---|---|---|
| **rs-fMRI + S-MRI** | | | | | | | | | |
| Manual | 27/24* | 70:30 | 97.8 | 98/97.6 | B | CNN | TPNAS-Net | ✓✓✓✓ | [370] |
| F score-AE | 368/449* | 10-fold | 85.1 | 81/89 | B+C | FCNN | - | ✗✓✓✓ | [371] |
| LLFS | 72/113*** | 4-fold | 80.8 | 84.9/79.2 | B | Ensemble | RF | ✓✓✓✓ | [372] |
| SMF-Graph based | 201/251* | 95:5 | 74.8 | - | B | DT | - | ?✓✓✓ | [373] |
| LLFS | 561/521* | - | 73 | - | B+C | Ensemble | RF | ✗✓?✓ | [374] |
| None | 481/526* | 10-fold | 72.7 | 67.8/76.6 | B+C | GNN | ANE + FCNN | ✗✓✓✓ | [375] |
| PCA-MRMR | 127/153* | 80:20 | 70 | - | B | Ensemble | RF | ✓✓✓✓ | [376] |
| None | 116/69* | 10-fold | 65.6 | 84/33 | B | DBN | - | ?✓✓✓ | [377] |
| AE-LLFS-PCA-ICA | 538/573* | 70:30 | 64.3 | 60/68.3 | B+C | SVM | Multi kernel | ✗✓✓✓ | [378] |
| VAE-LLFS | 539/573* | - | 62.6 | - | B+C | GNN | - | ✗✓?✓ | [379] |
| **rs-fMRI + DWI/DTI** | | | | | | | | | |

| | | | | | | | | | |
|---|---|---|---|---|---|---|---|---|---|
| Graph based | 403/468* | 80:20 | 60.9 | 53.5/69.4 | B+C | SVM | RBF kernel | ✕✓✓✓ | [380] |
| **rs-fMRI + S-MRI + DWI/DTI** | | | | | | | | | |
| LLFS | 46/47 | 66:33 | 92.5 | 97.8/87.2 | B | Ensemble | CRF | ✓✓✓✓ | [381] |
| LLFS | 31/23* | LOOCV | 72.3 | - | B | SVM | Linear kernel | ?✓✓✓ | [382] |
| **T-fMRI + DWI/DTI + Clinical data** | | | | | | | | | |
| RFE-LLFS-t test | 15/15 | 53:47 | 95.9 | 96.9/94.8 | B+C | SVM | Linear kernel | ?✓✓✓ | [383] |
| **S-MRI + DWI/DTI** | | | | | | | | | |
| PCA | 110/83*** | 10-fold | 93.3 | 93.6/96.2 | B | SVM | RBF kernel | ✓✓✓✓ | [384] |
| RFE | 58/48 | 5-fold | 88.8 | 93/83.8 | B | Ensemble | RF | ✓✓✓✓ | [385] |
| t test | 16/16 | LOOCV | 86.7 | 87.3/86.1 | B | SVM | RBF kernel | ✓✓✓✓ | [386] |
| VBM | 14/33 | 70:30 | 75.3 | 24.8/97 | B | Ensemble | RF | ✓✓✓✓ | [387] |
| **EEG + Eye tracking** | | | | | | | | | |
| PCA-FFS | 24/28 | 80:20 | 100 | - | - | NB | Gaussian | ?✓✓? | [388] |
| None | 21/21 | 80:20 | 95 | 95/95 | B | GNN | GFT + FCNN | ✓✓✓? | [389] |
| MRMR | 49/48 | - | 85.4 | - | B | SVM | Linear kernel | ✓✓?✓ | [390] |
| **rs-fMRI + Genetic** | | | | | | | | | |
| RFE-t test | 47/24*** | LOOCV | 86 | 81/88 | B+C | SVM | Multi kernel | ✓✓✓✓ | [391] |
| **Kinematic + Eye tracking** | | | | | | | | | |
| t test-LLFS | 22/22 | 68:32 | 78 | 57/99 | C | SVM | RBF kernel | ✓-✓✕ | [392] |
| **S-MRI + DTI + MRS** | | | | | | | | | |
| RFE | 19/18 | LOOCV | 91.9 | - | C | DT | - | ✓✓✓✓ | [393] |

Acc: Accuracy, ASD: Autism spectrum disorder, LOOCV: Leave-one-out cross-validation, Sen: Sensitivity, Spe: Specificity, TD: Typically developing.

DWI/DTI: Diffusion-weighted imaging/Diffusion-tensor imaging, EEG: Electroencephalography, rs-fMRI: Resting-state functional magnetic resonance imaging, S-MRI: Structural magnetic resonance imaging, T-fMRI: Task-based functional magnetic resonance imaging.

The classifications for the age are A: Toddlers (<2y), B: Children and adolescents (2-18y), C: Adults (>18y).

For the risk of bias, the first dot on the left is indicative of the patient selection domain, next is for the index test (modality) domain, next one is for the index test (AI algorithm) domain, and the last one on the right is indicative of the reference standard domain. ✓ means low risk of bias, ? means unclear risk of bias, ✕ means high risk of bias, and - means that domain is not applicable for the study.

* Data is from the ABIDE dataset

** Data is from the Kaggle dataset.

*** Data is from the NDAR dataset.

<u>1:</u> AAL: Automated anatomical labelling atlas, BASC: Bootstrap analysis of stable clusters, CC200: Craddock-200 atlas, CC400: Craddock-400 atlas, DES: Destrieux atlas, DK: Desikan–Killiany atlas, DLA: Dictionary learning algorithm, DOS: Dosenbach atlas, HBM: Hierarchical Bayesian model, HO: Harvard-Oxford atlas, IP-TFP: Instantaneous phase transfer function perturbation, RBS: Region based segmentation, RSN: Resting-state networks, SBC: Social brain connectome atlas

<u>2:</u> AE: Autoencoder, BOF: Bag of feature, BS: Bootstrapping, DL: Dictionary learning, DR: Dual regression, DSM: Dominant-sequence model, EIIC: Extended invariant information clustering, EW: Element-wise filters, FFS: Forward feature selection, GA: Genetic algorithm, GARCH: Generalized autoregressive conditional heteroscedasticity, HMM: Hidden Markov model, IBP: Indian buffet processes, ICA: Independent component analysis, L1SCCA: L1-norm regularized sparse canonical correlation analysis, LASSO: Least absolute shrinkage and selector operation, LEAN: Layer-wise elimination of accessory nodes, LLFS: Local learning-based feature selection, MCNFD: Multi-site clustering and nested feature extraction, MI: Mutual information, MRMR: Maximal relevance and minimal redundancy, MSDL: Multi-subject dictionary learning, MSTEPS: Multiscale stepwise selection, PCA: Principal component analysis, RFE: Recursive feature elimination, S2n: Signal to nose ratio, SLR: Sparse low-rank representation, SMF: Spatial feature based detection, VBM: Voxel based morphometry, WT: Wavelet transform

**3:** AE: Autoencoder, CNN: Convolutional neural network, DBN: Deep belief network, DFA: Discriminant factor analysis, DNN: Deep neural network, DRBM: Discriminative restricted Boltzmann machine, DT: Decision tree, FCNN: Fully-connected neural network, FLDA: Fisher's linear discriminant analysis, GNN: Graph neural network, KNN: K-nearest neighbors, LDA: Linear discriminant analysis, LMT: Logistic model trees, NB: Naïve Bayes, LR: Logistic regression, LSTM: Long-short term memory, PNN: Probabilistic neural network, PSN-ANFIS: Particle swarm optimization-Adaptive network-based fuzzy inference system, PTN: Prototypical network, QDA: Quadratic discriminant analysis, RNN: Recurrent neural network, RR: Ridge regression, SNCAE: Stacked nonnegativity constraint autoencoder, Sparse-MVTC: Sparse multi-view task-centralized learning, STA: Scanpath trend analysis, SVM: Support vector machine, VMM: Variable-order Markov model

**4:** AE: Autoencoder, ANE: Attention-based node-edge, CNN: Convolutional neural network, CRF: Conditional random forest, DANN: Deep attention neural network, FCNN: Fully-connected neural network, GAT: Graph attention network, GFT: Graph Fourier transform, GRU: Gate recurrent unit, hi-GCN: Hierarchical graph convolutional network, ID: Invertible dynamic, LR: Logistic regression, LR-GCN: Low-rank subspace graph convolutional network, LSTM: Long-short term memory, MT: Multi template, PBL-McRBFN: Projection based learning metacognitive radial basis function network, PLS: Partial least squares, PNN: Probabilistic neural network, PTN: Prototypical network, RBF: Radial basis function, RF: Random forest, RGF: Regularized greedy forest, RNN: Recurrent neural network, s-GCN: Siamese graph convolutional network, SAE: Sparse autoencoder, SFCNN: Siamese fully connected neural network, Sparse-MVTC: Sparse multi-view task-centralized learning, TL: Transfer learning, TPNAS-Net: Topology preserving neural architecture search network
