## Supplementary material for "Automated diagnosis of autism: State of the art": Multimedia Appendix 3: Results of SCS meta-analyses

| Feature set | # Studies | N | Acc | SCS (95% CI) | | Feature set | # Studies | N | Acc | SCS (95% CI) | |
|---|---|---|---|---|---|---|---|---|---|---|---|
| | | | | Sen | Spe | | | | | Sen | Spe |
| **rs-fMRI** | | | | | | | | | | | |
| 3D fMRI | 2 | 222 | 81-94.7 | - | - | Non-oscillatory connectivity | 1 | 72 | 80 | - | - |
| Dynamic functional connectivity | 18 | 7081 | - | 0.74 (0.67-0.79) | 0.74 (0.68-0.79) | Normalized image | 1 | 172 | 57.8 | - | - |
| Effective connectivity | 2 | 340 | 69-87 | - | - | Power spectral density | 2 | 481 | 92-96.2 | - | - |
| Graph metrics | 19 | 11913 | - | 0.74 (0.69-0.78) | 0.69 (0.64-0.74) | ROI-based functional connectivity | 55 | 39697 | - | 0.74 (0.69-0.79) | 0.76 (0.71-0.81) |
| Graph metrics + Clinical data | 1 | 575 | 74.5 | - | - | ROI-based functional connectivity + Amplitude of low-frequency fluctuation | 1 | 184 | 68.5 | - | - |
| High-order functional connectivity | 4 | 1648 | 68.8-81 | - | - | ROI-based functional connectivity + Clinical data | 2 | 1209 | 65-73.2 | - | - |
| Histogram of oriented gradients + Clinical data | 1 | 1111 | 65 | - | - | ROI-based functional connectivity + Clinical data + Information theory | 1 | 871 | 72.5 | - | - |
| Independent components | 3 | 1023 | 77.7-89.5 | - | - | Wavelet-based dynamics features | 1 | 54 | 86.7 | - | - |
| **S-MRI** | | | | | | | | | | | |
| Cortical thickness | 3 | 154 | 84.2-90 | - | - | Surface-based morphological features + Cortical thickness | 2 | 934 | 60-96.3 | - | - |
| Curvelet sub-bands | 1 | 151 | 75 | - | - | Surface-based morphological features + Clinical data | 1 | 50 | 88 | - | - |
| Histogram of oriented gradients | 1 | 250 | 65 | - | - | Surface-based morphological features + Volume-based morphological features + Cortical thickness | 2 | 220 | 93.8 | - | - |
| Histogram of oriented gradients + Clinical data | 1 | 1111 | 64.1 | - | - | Volume-based morphological features | 8 | 1251 | - | 0.81 (0.69-0.89) | 0.83 (0.71-0.90) |
| Normalized image | 3 | 1561 | 71-99 | - | - | Volume-based morphological features + Clinical data | 1 | 523 | 64 | - | - |
| Path signature + Clinical data | 1 | 60 | 87 | - | - | Volume-based morphological features + Cortical thickness | 1 | 117 | 96.3 | - | - |
| Structural covariance network | 1 | 1085 | 71.8 | - | - | Volume-based morphological features + Voxel density | 1 | 58 | 82 | - | - |
| Surface-based morphological features | 3 | 1125 | 75-83 | - | - | Voxel density | 1 | 131 | 73.3 | - | - |
| **EEG** | | | | | | | | | | | |
| Complex networks | 2 | 84 | 81.7-94.7 | - | - | Event-related potentials | 3 | 253 | 70-90.4 | - | - |
| Entropy + Frequency-domain + Time-domain + Non-linear features | 1 | 98 | 97.9 | - | - | Frequency-domain + Non-linear features | 1 | 28 | 78.5 | - | - |
| Entropy + Frequency-domain features | 4 | 488 | 86-99.7 | - | - | Frequency-domain features | 6 | 313 | - | 0.98 (0.93-0.99) | 0.93 (0.84-0.97) |
| Entropy + Non-linear features | 3 | 178 | 90.2-97 | - | - | Non-linear features | 1 | 61 | 82 | - | - |
| **Eye tracking** | | | | | | | | | | | |
| Face-to-face conversations | 1 | 39 | 92.3 | - | - | Virtual reality | 2 | 162 | 73-86 | - | - |
| Interactions with parents | 1 | 32 | 93.8 | - | - | Watching faces | 1 | 77 | 88.5 | - | - |

| | | | | | | | | | | | |
|---|---|---|---|---|---|---|---|---|---|---|---|
| Observing images | 2 | 67 | 59.3-92 | - | - | Watching videos | 3 | 271 | 82-87.5 | - | - |
| Saliency maps | 4 | 169 | 62.1-99.8 | - | - | Watching websites | 1 | 30 | 91.6 | - | - |
| **T-fMRI** | | | | | | | | | | | |
| Biological motion perception task | 1 | 40 | 64.5 | - | - | Dynamic bodies attention task | 1 | 29 | 92.3 | - | - |
| Biopoint task | 2 | 233 | 76-79.8 | - | - | Language task | 1 | 60 | 75.8 | - | - |
| Cognitive control task | 1 | 58 | 82.8 | - | - | Response to speech | 3 | 266 | 77.2-86 | - | - |
| **DWI/DTI** | | | | | | | | | | | |
| Fiber density + Fiber bundle cross-section | 1 | 52 | 73.1 | - | - | Fractional anisotropy + Mean diffusivity + Axial diffusivity + Radial diffusivity + Skewness | 2 | 323 | 73-94.7 | - | - |
| Fractional anisotropy | 1 | 73 | 75.3 | - | - | Graph metrics | 1 | 94 | 68 | - | - |
| Fractional anisotropy + Mean diffusivity | 2 | 224 | 78.3-80 | - | - | | | | | | |
| **Facial recognition** | | | | | | | | | | | |
| Facial attribute recognition | 1 | 88 | 72.9 | - | - | Static facial features | 5 | 12658 | - | 0.90 (0.86-0.93) | 0.90 (0.86-0.93) |
| **Other modalities** | | | | | | | | | | | |
| Functional near-infrared spectroscopy | 3 | 138 | 91.9-95.7 | - | - | Response to name | 1 | 43 | 93 | - | - |
| Kinematic features | 5 | 225 | - | 0.88 (0.78-0.94) | 0.91 (0.82-0.96) | Video analysis | 2 | 361 | 82-88.9 | - | - |
| PET scan | 1 | 58 | 88 | - | - | Vocal analysis | 1 | 2612 | 86.5 | - | - |
| **Multimodal** | | | | | | | | | | | |
| EEG + Eye tracking | 1 | 42 | 95 | - | - | rs-fMRI + S-MRI | 6 | 3356 | - | 0.75 (0.56-0.88) | 0.68 (0.48-0.83) |
| Kinematic features + Eye tracking | 1 | 44 | 78 | - | - | rs-fMRI + S-MRI + DWI/DTI | 1 | 93 | 92.5 | - | - |
| rs-fMRI + DWI/DTI | 1 | 871 | 60.9 | - | - | S-MRI + DWI/DTI | 4 | 378 | 75.3-93.3 | - | - |
| rs-fMRI + Genetic | 1 | 71 | 86 | - | - | T-fMRI + DWI/DTI + Clinical data | 1 | 30 | 95.9 | - | - |
